# Assessing AI and Neurologist Diagnostic Reasoning Against Neuropathological Ground Truth

**DOI:** 10.64898/2026.07.07.26356930

**Authors:** Yu Leng, Ayush Noori, Yingnan He, Anna Favaro, John R. Dickson, Alberto Serrano-Pozo, Marina Avetisyan, Diego Rodriguez, Eric S. Rosenberg, Derek H. Oakley, Vikram Khurana, Julia Maslinski, Benjamin S. Glicksberg, Mahanaz Syed, Paige Ellen Kubon, Jamie Walker, Claudia K. Suemoto, Sudha Seshadri, David A. Clifton, Marinka Zitnik, Bradley T. Hyman, Matthew P. Frosch, Sudeshna Das

**Author notes:** Equal contribution (Yu Leng, Ayush Noori, Yingnan He). <u>Correspondence to</u>: Sudeshna Das, PhD, Full address: 65 Landsdowne St., Cambridge, MA 02139.

## Abstract

**BACKGROUND:** Accurate differential diagnosis of complex neurological disorders remains challenging due to overlapping clinical features and heterogeneous disease presentations. Although large language models (LLMs) show promise in clinical reasoning, prior studies have benchmarked performance against clinician consensus rather than biological ground truth. Evaluation of diagnostic AI in neurology requires benchmarks grounded in neuropathological findings.

**METHODS:** We introduce NeuroBench, a curated benchmark of complex neurological cases with neuropathologically confirmed reference-standard diagnoses, and DIAGNO, a confidence-aware LLM-based system for neurological diagnosis. NeuroBench comprises 791 retrospective case summaries and corresponding autopsy-confirmed diagnoses across three independent health systems: 500 cases from the Massachusetts General Hospital (MGH) Brain Cutting Conference; 200 cases from the Mount Sinai Health System; and 91 cases from the University of Texas Health Science Center at San Antonio (UTHSA). Across all cases, DIAGNO generated top-3 differential diagnoses, employing retrieval-augmented generation for lower-confidence cases. In a subset of 203 complex MGH cases, performance was assessed by three independent blinded adjudicators who evaluated both DIAGNO and neurologists against neuropathological ground truth.

**RESULTS:** NeuroBench encompassed 59 unique neuropathological diagnoses, spanning conditions including cerebrovascular disease, brain tumors, neurological infections, and various neurodegenerative and inflammatory disorders. DIAGNO achieved 83.8% top-3 accuracy across the 500 MGH cases, 76.5% accuracy across the 200 Mount Sinai cases, and 92.3% accuracy across the 91 UTHSA cases. In the subset of 203 MGH cases, DIAGNO achieved higher top-3 accuracy (0.67 versus 0.63) and taxonomy-level accuracy (0.74 versus 0.67) than neurologists. In cases of disagreement, DIAGNO was more often correct than neurologists (29 versus 19 cases). Diagnostic concordance between DIAGNO and neurologists was high (90% agreement in top-3 predictions). In a real-world evaluation on eight cases from Mass General Brigham, neurologists rated DIAGNO’s reasoning favorably (mean 4.03/5) across multiple dimensions.

**CONCLUSIONS:** NeuroBench establishes neuropathological confirmation as a reference standard for evaluating diagnostic AI in neurology, moving beyond clinician-based benchmarking to redefine the ceiling of diagnostic accuracy. Evaluated against this standard, DIAGNO achieved diagnostic performance comparable to neurologists and received favorable clinician ratings in real-world applications, supporting its potential as a clinical decision support tool in neurology.

## Introduction

Accurate differential diagnosis in complex neurological disorders remains inherently challenging, especially in ambiguous clinical presentations^1^. Many neurological diseases share overlapping clinical features, making it difficult to distinguish etiologies based on symptoms and exam findings alone^2^, and even experienced specialists may struggle to diagnose challenging cases. For example, different dementias can present with similar cognitive deficits and behavioral manifestations^3–5^, and idiopathic Parkinson’s disease and atypical parkinsonism share overlapping motor symptoms^6^. Additionally, autopsy studies have shown that premortem clinical diagnoses may be incomplete or incorrect, with postmortem examination sometimes revealing unexpected pathologies even after expert human evaluation^7,8^. For instance, a case of rapidly progressive dementia was ultimately found at autopsy to reflect coexisting paraneoplastic encephalomyelitis and cerebral amyloid angiopathy^9^. These challenges underscore that definitive neurological diagnosis requires postmortem neuropathological examination, which remains the only biological ground truth.

Large language models (LLMs) offer a promising direction for clinical reasoning and diagnostic support^10–15^, with several studies reporting performance matching or exceeding physicians on challenging case vignettes and diagnostic tasks^16,17^. Despite these gains, recent benchmarking studies have shown that differential diagnosis, the cornerstone of early clinical reasoning, remains a major failure point^18^. Early studies are also beginning to evaluate the use of LLMs in neurology. Schubert et al. showed that GPT models can match or exceed expert reasoning on neurology board-style questions^19^. Hewitt et al.^20^ reported low zero-shot GPT-4o accuracy on 30 synthetic glioma cases (13.3%) but near-perfect performance (99.3%) with retrieval-augmented generation (RAG)^21^. Koga et al. reported 52% primary diagnostic accuracy for ChatGPT on 25 neurodegenerative cases ^22^. However, these studies were limited by synthetic data, small samples, or narrow disease scope. Most critically, no prior work has systematically evaluated LLM performance against neuropathological ground truth across diverse neurological conditions. Prior evaluations that benchmark AI against clinical diagnoses treat clinician judgment as a reliable reference standard, obscuring whether observed performance reflects true diagnostic accuracy or shared limitations of premortem information.

To address these gaps, we introduce NeuroBench, a curated benchmark of 791 neuropathologically confirmed neurological cases across three sites: 500 cases from Massachusetts General Hospital (MGH), 200 cases from the Mount Sinai Health System, and 91 cases from the University of Texas Health Science Center at San Antonio (UTHSA). We publicly release a subset of these cases for evaluation of AI for differential diagnosis of complex neurological disorders. Next, we introduce one such algorithm: DIAGNO (<u>D</u>ual-stage Inference with <u>A</u>ugmented <u>G</u>eneration for <u>N</u>eurological <u>O</u>utcomes), a confidence-aware diagnostic system enhanced with RAG^21^. By benchmarking both DIAGNO and board-certified neurologists against autopsy-confirmed diagnoses, we establish a framework that evaluates diagnostic performance against biological ground truth rather than clinician opinion. We further assessed diagnostic concordance and inter-adjudicator reliability, and validated DIAGNO on complex published cases and real-world cases evaluated by neurologists. This study provides the first systematic evaluation of LLM-assisted diagnostic reasoning across diverse, pathologically confirmed neurological disorders and reframes diagnostic benchmarking from AI versus clinicians to performance at the limits of modern medicine.

## Methods

### Study design and data source

For this retrospective study, we curated NeuroBench, a benchmark dataset of 791 autopsy-confirmed neurological case summaries from three independent healthcare institutions. For each donor, NeuroBench comprises two components: (1) a clinical case summary and (2) a corresponding reference-standard diagnosis (Figure 1A). Across all three sites, reference-standard diagnoses were determined through postmortem neuropathological examination by board-certified neuropathologists, providing a biological ground truth rarely available in clinical AI benchmarking studies.

**Figure 1.**
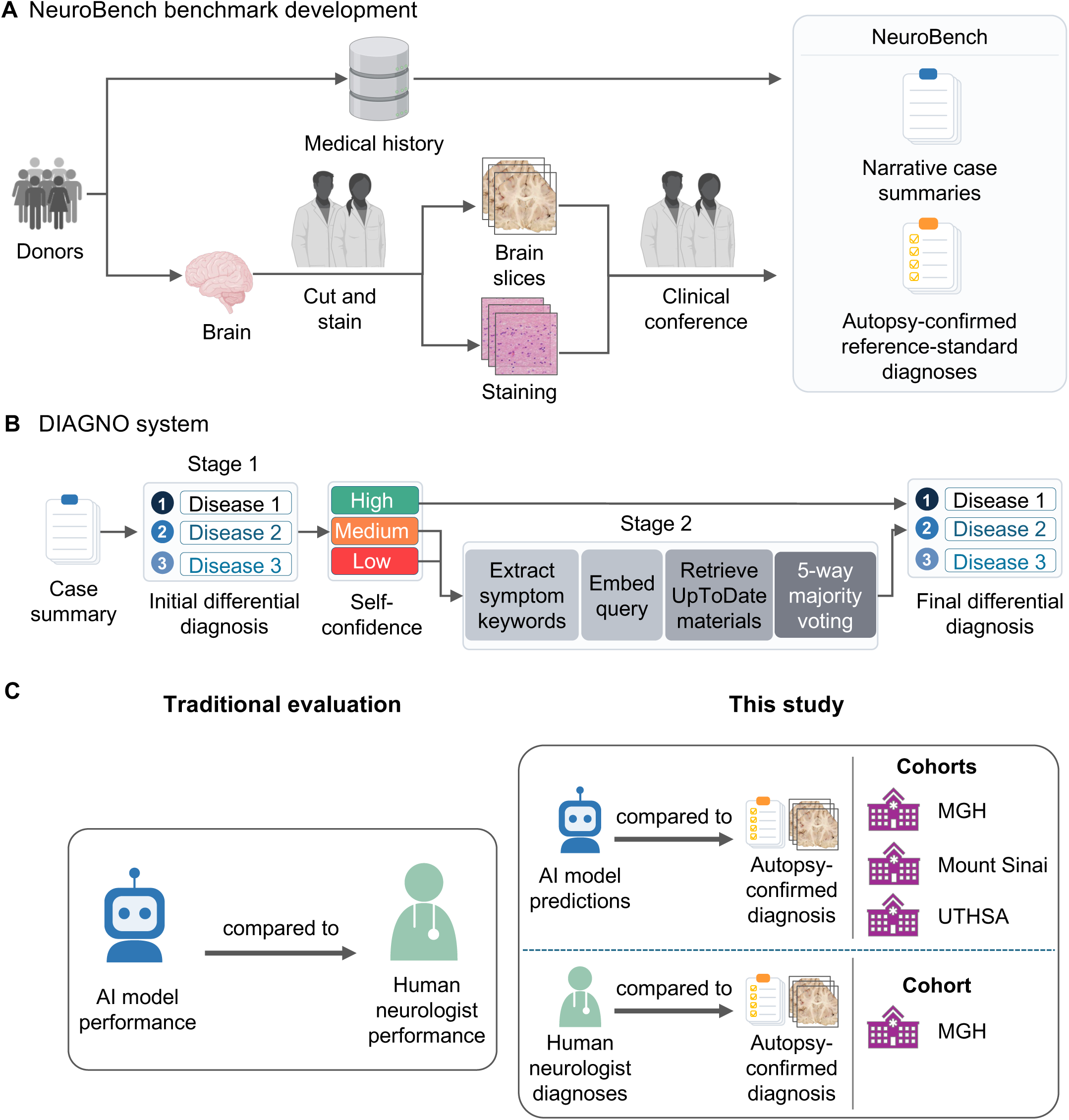
Overview of NeuroBench and the evaluation paradigm. **(A) NeuroBench development.** Each brain donor contributes a clinical case summary and brain tissue, which undergoes neuropathological examination, cutting, staining, and review at a clinical conference to establish a reference-standard neuropathological diagnosis. **(B) DIAGNO architecture.** DIAGNO is a confidence-aware diagnostic system with LLM-based adaptive knowledge retrieval. **(C) Evaluation paradigm.** In this study, DIAGNO and neurologists independently generated differential diagnoses from the same NeuroBench case summaries, which were compared with autopsy-confirmed neuropathological diagnoses at the exact-diagnosis and taxonomy levels.

### Massachusetts General Hospital

We included 500 cases from the Brain Cutting Conference at MGH, a teaching forum for medical trainees predominantly in neurology residency and fellowships but also including medical students, residents, and fellows from related disciplines (pathology, psychiatry, neuroradiology, physical medicine and rehabilitation). Cases spanned December 2007 to December 2019, selected based on availability of sufficient clinical information, autopsy consent, and diagnostic complexity. Both typical and diagnostically challenging cases were included. The original Brain Cutting Conference materials were maintained internally by the MGH Neuropathology Division as clinical case summaries used for conference discussions. Case summaries were written by residents or fellows from and summarized the complete clinical course from symptom onset to autopsy, including medical history, neurological examination findings, cognitive testing, laboratory tests, and outcomes. Key diagnostic tests (*e.g.*, neuroimaging) were deliberately withheld from initial summaries and disclosed only after discussion to preserve diagnostic challenge and reflect real-world clinical reasoning under uncertainty. The internal case identifiers linking the summaries to the corresponding neuropathological diagnoses were handwritten on printed copies of the conference materials. Four members of the study team manually transcribed these identifiers and linked each case summary to its corresponding neuropathological diagnosis in the neuropathology database to create NeuroBench. Neuropathological diagnoses were subsequently reviewed and harmonized into a standardized set of diagnoses to ensure consistent terminology across cases. The harmonized diagnoses were further categorized using a hierarchical diagnostic taxonomy to group related neuropathological diagnoses into broader disease categories. The taxonomy, defined based on expert consensus, consolidated related disease subtypes and synonymous terms under broader diagnostic categories^23,24^ (*e.g.*, ischemic infarcts and hemorrhages were grouped under cerebrovascular disease). These materials were maintained exclusively as internal teaching records and were never publicly accessible or available online, reducing the possibility that they were included in any LLM pre-training corpora and thus minimizing the risk of data leakage. The MGH Institutional Review Board approved this study (protocol number: 2015P001915).

### Mount Sinai Health System

We constructed a neuropathology cohort from longitudinal electronic health records across the Mount Sinai Health System, a major hospital network located in New York City. Cases were required to have at least 10 antemortem clinical notes. All eligible neurodegenerative and hydrocephalus cases were included, while cases from the remaining diagnostic categories were randomly sampled to approximate the diagnostic distribution of the MGH cohort, yielding a cohort of 238 cases. To identify neuropathological diagnoses, candidate autopsy notes were retrieved among notes dated on or after the patient’s recorded date of death. Notes were required to contain (1) an autopsy-related term (“autopsy,” “postmortem,” or “post mortem”); (2) at least one structural autopsy-report anchor (“final anatomic diagnosis,” “cause of death,” “gross description,” or “gross examination”); and (3) at least one neuropathology related term (“neuropathology,” “neuropathological,” “brain cutting,” or “brain weight”). When multiple qualifying reports were identified for the same patient, the most recent report was selected. Any ties on report date were resolved by selecting the report with the greatest text length.

Using a secure, Mount Sinai-approved Azure OpenAI API, longitudinal clinical summaries were generated from the antemortem clinical notes and final diagnoses made by board-certified neuropathologists were extracted from the autopsy reports and mapped to structured diagnoses. Clinical summaries were generated without access to the neuropathology final diagnoses, and diagnostic imaging and pathology reports were withheld. Cases with more than one applicable neuropathologic disease process were permitted to map to multiple categories. All model-generated diagnoses and clinical summaries were manually reviewed for accuracy and diagnostic leakage. Cases lacking an eligible neuropathologic ground-truth diagnosis or an adequate clinical summary after review were excluded, yielding a final evaluation cohort of 200 cases. The Mount Sinai Institutional Review Board approved this study (protocol number: STUDY-20-00338).

### University of Texas Health Science Center at San Antonio

At UTHSA, all brain donors were recruited from the community, various Neurology and Dementia clinics, or from longitudinal South Texas Alzheimer’s Disease Research Center cohorts. For each donor, a physician trainee without specialized training in neurology prepared a clinical summary based on detailed interviews with available next of kin or other knowledgeable informants, supplemented by a review of available medical records and antemortem neuroimaging. The clinical evaluation included cognitive and functional status at the time of death, medical comorbidities, neurological diagnoses, and dementia subtype, where applicable. Clinical summaries and diagnoses were finalized without knowledge of the neuropathological findings. Board-certified neuropathologists subsequently performed postmortem evaluations and assigned the primary neuropathological diagnosis for each case. All 91 cases had both clinical and neuropathological evaluations. Case summaries without the clinical diagnoses were used as input to DIAGNO. Written informed consent for brain donation and use of donor data was obtained from the donors or their legally authorized representatives.

### Development of DIAGNO AI system

To mirror the human reasoning and decision-making process, we developed DIAGNO (<u>D</u>ual-stage Inference with <u>A</u>ugmented <u>G</u>eneration for <u>N</u>eurological <u>O</u>utcomes), a confidence-aware diagnostic system with LLM-based adaptive knowledge retrieval (Figure 1B). For each case, DIAGNO was prompted to simulate an expert neurologist, analyze the clinical summary, and generate a list of three differential diagnoses. DIAGNO generates predictions in two stages. First, the model produces the following structured output: (1) a top-3 ranked differential diagnosis list; (2) a model-generated categorical confidence assessment (“Low,” “Medium,” or “High”); and (3) a brief summary of its reasoning based on the case content. Cases assigned Low or Medium confidence proceed to Stage 2, whereas High-confidence cases bypass retrieval and retain the Stage 1 output. The categorical confidence assessment serves as a confidence signal for adaptive retrieval, consistent with recent studies using LLM self-assessment and uncertainty estimation to guide retrieval augmentation and confidence-aware inference^25,26^. In Stage 2, DIAGNO activates a RAG pipeline to simulate clinical reasoning under uncertainty. Relevant reference materials are retrieved from a curated collection of UpToDate articles^27^, which were preprocessed, embedded using Bio+ClinicalBERT ^28^ and indexed in Pinecone for semantic retrieval. At Mount Sinai, where Pinecone access was not possible, the embeddings were stored in a local vector database.

To tailor retrieval to each patient case, DIAGNO first extracts a set of key symptom-related terms from the clinical summary. Then, embeddings are generated from both the case summary and the extracted keywords, with equal or custom-assigned weights. The resulting embeddings are used to query the Pinecone index and retrieve the top-5 semantically relevant passages. The retrieved content is appended to the prompt, and the model re-generates diagnoses across five independent trials, with the final output determined by majority vote^29^. A detailed example of the prompt and corresponding response is provided in Supplementary Note 1. The prompt also included the standardized set of diagnoses used across the 791 NeuroBench cases.

### Benchmarking DIAGNO on NeuroBench across language models

The DIAGNO framework can flexibly accommodate any underlying frontier language model. To understand how performance varies by model choice, we benchmarked DIAGNO on NeuroBench across several backbone LLMs, holding the prompts, confidence gate, retrieval depth, and trial count constant across models. At MGH, case summaries were fully de-identified before inference. We benchmarked DIAGNO using GPT-4o (2024-05-01-preview) via the Microsoft Azure AI Foundry; GPT-5 (2025-08-07) and GPT-6 Astra (2026-09-03), both via the Harvard Medical School Azure OpenAI gateway; and Claude Opus 5.5. At Mount Sinai, we benchmarked DIAGNO on GPT-5.6 Sol (2026-09-07) through a secure, Mount Sinai-approved Azure OpenAI API. At UTHSA, we benchmarked DIAGNO on GPT-5 (2025-08-07). Across all benchmarking experiments, Stage 1 was run at temperature 0.1 and Stage 2 was run with top-5 retrieval and five independent trials at temperature 0.3 followed by majority vote synthesis. Models for which sampling temperature was not available (GPT-5, GPT-6 Astra, GPT-5.6 Sol, and Claude Opus 5.5) were run using their default reasoning settings.

### Comparing DIAGNO and expert neurologists against neuropathological ground truth

Next, we sought to compare DIAGNO performance in diagnosing complex neurological disorders with that of expert human neurologists, using neuropathologically confirmed diagnoses as the reference standard (Figure 1C). We evaluated performance on a subset of complex NeuroBench cases from MGH. Unlike the frontier LLM benchmarking described above, DIAGNO was prompted to generate a ranked top-three differential diagnosis in free text without being constrained to the standardized NeuroBench diagnosis set, more closely reflecting clinical diagnostic reasoning. We implemented DIAGNO with GPT-4o via a secure on-premises deployment within the Mass General Brigham (MGB) network infrastructure; this allowed us to make age and gender information available to both DIAGNO and the neurologists to reflect realistic diagnostic context, while excluding these variables from both frontier LLM benchmarking and publicly shared examples to protect patient privacy. After de-identification, we released the complex NeuroBench cases as a community benchmarking resource.

For this study, three board-certified neurologists (JRD, AS-P, MA), each with more than 10 years of clinical practice experience, independently reviewed non-overlapping subsets of these cases to produce expert differential diagnoses specifically for this study. Cases were randomly assigned to one of the three neurologists, with each case reviewed by a single neurologist. They collectively spent approximately 40 hours reviewing the cases and generating their differential diagnoses. Neurologists reviewed the same curated case summaries provided to DIAGNO and independently produced a ranked list of the top three differential diagnoses for each case. They were encouraged to consult external resources normally used in clinical practice, such as UpToDate or other medical references, as needed.

For this evaluation, the original neuropathological diagnoses were retained at their full diagnostic granularity rather than harmonized to the standardized NeuroBench diagnostic categories. These reference diagnoses frequently included specific neuropathological subtypes (e.g., fronto-temporal lobar degeneration, TDP43 subtype); DIAGNO and neurologists generated clinical differential diagnoses that could vary in terminology and level of specificity. Therefore, direct string matching between the generated differential diagnoses and neuropathological reference diagnoses would not reliably capture diagnostically concordant responses.

To account for these differences in terminology and diagnostic granularity, three independent adjudicators—two human experts (BTH, a behavioral neurologist, and DHO, a neuropathologist) and one LLM adjudicator^30^ (GPT-4o)—assessed the anonymized, blinded differential diagnoses generated by DIAGNO and the neurologists. DIAGNO and neurologist responses were presented in randomized order. Each top-1, top-2, and top-3 differential diagnosis was assessed against the neuropathological reference diagnosis using binary scoring (correct/incorrect), with the LLM adjudicator additionally prompted to provide brief justifications (Supplementary Note 2). This approach enabled comparison of DIAGNO and neurologist performance against the same neuropathological reference standard while accounting for differences in diagnostic terminology and specificity.

### Performance evaluation metrics

DIAGNO performance across LLMs and datasets was evaluated using top-1, top-2, and top-3 diagnostic accuracy. Top-1 accuracy was defined as the proportion of cases for which the first-ranked diagnosis was correct; top-2 accuracy as the proportion for which the correct diagnosis appeared within the first two ranked outputs; and top-3 accuracy as the proportion for which the correct diagnosis appeared anywhere within the top three outputs^11^.

For LLM benchmarking across the 791 NeuroBench cases, exact diagnosis accuracy was determined by string matching the predicted diagnoses to the corresponding standardized neuropathological diagnoses. Taxonomy-level accuracy was additionally assessed by mapping both the predicted and neuropathological diagnoses to their corresponding diagnostic taxonomy categories and determining category-level matches. This approach better reflects real-world diagnostic reasoning, where identifying the correct disease class often guides disease management even if the precise subtype is unspecified.

For the comparison of DIAGNO with expert human neurologists on the 203 complex MGH cases, performance was evaluated against the original neuropathologically confirmed diagnoses, which retained their full diagnostic granularity. Because these diagnoses could differ from the generated clinical differential diagnoses in terminology and level of specificity, correctness was independently assessed by three adjudicators (BTH, DHO, and LLM) rather than by string matching. Top-1, top-2, and top-3 accuracies were computed separately for DIAGNO and human neurologists. We report adjudicator-specific correctness and “any-adjudicator” correctness, defined as cases rated correct by at least one adjudicator. 95% confidence intervals (CIs) for diagnostic accuracy were calculated using the Wilson score method for binomial proportions. Differences in paired diagnostic accuracy were calculated at the case level, with 95% CIs estimated using a nonparametric paired bootstrap with 10,000 resamples and percentile-based confidence limits.

We measured inter-adjudicator agreement^31^ using Cohen’s kappa coefficient^32^ for all pairwise comparisons among the three adjudicators, computed separately for DIAGNO-generated and neurologist-generated differential diagnoses. Kappa values were interpreted using standard thresholds (κ < 0.61 = poor agreement; κ = 0.61–0.80 = substantial agreement; κ = 0.81–1.00 = almost perfect agreement).

To evaluate the degree of diagnostic alignment between DIAGNO and human neurologists independent of correctness relative to the neuropathological reference diagnosis, we computed concordance at the top-1, top-2, and top-3 levels using a rank-aware matching strategy^33^ (Supplementary Note 3). Top-1 concordance required an exact match between both top-ranked diagnoses; top-2 concordance required any overlap within the top two diagnoses; and top-3 concordance required any overlap across all three diagnoses. This approach captures varying levels of diagnostic convergence and provides insight into alignment between DIAGNO- and neurologist-generated differential diagnoses.

### Statistical analysis

Subgroup analyses were conducted across both the 31 taxonomies and five broader diagnostic classes: cerebrovascular disease (CVD), infectious/inflammatory disorders, neurodegenerative diseases, neoplastic disorders, and other conditions. Accuracy was calculated for DIAGNO and neurologists within each category. Differences in accuracy were examined using chi-square tests, with standardized residuals identifying major contributors. The two-sided McNemar’s test was used for paired comparisons of DIAGNO versus neurologist performance within each subgroup.

### Error analysis

We conducted an error analysis to characterize the types of diagnostic failures observed when neither DIAGNO nor the neurologist included the correct diagnosis within their top-3 differential lists. For each such case, diagnoses were mapped to taxonomy categories to evaluate patterns in misclassification across disease groups.

To further understand sources of diagnostic error, we performed a qualitative typology analysis ^34^ using pre-autopsy clinical diagnoses from medical records and corresponding autopsy reports. Four overarching categories of error were identified: (1) clinically inaccessible information, in which decisive diagnostic evidence was only available postmortem; (2) information gaps, reflecting missing or withheld clinical details such as imaging or laboratory results; (3) diagnostic reasoning errors, where the correct diagnosis was theoretically attainable but missed; and (4) knowledge gaps involving rare or atypical conditions.

For cases with final diagnoses involving tumors or CVD, DIAGNO was additionally prompted to recommend appropriate next diagnostic steps and indicate the urgency of each recommendation using only the information in the case summary. Recommendations were reviewed to assess clinical appropriateness, with particular attention to whether neuroimaging was suggested.

### External and real-world evaluation

To assess diagnostic generalizability, we additionally evaluated DIAGNO on 40 challenging neurological case reports published in *The New England Journal of Medicine* (*NEJM*) from 2020 to 2025 (Table S2). DIAGNO generated top-3 differential diagnoses using the identical pipeline, with performance assessed by the LLM adjudicator against published final diagnoses.

To further evaluate clinical applicability, we conducted an expert assessment using eight real-world cases from two hospitals of Mass General Brigham (MGB) healthcare: MGH and Brigham and Women’s Hospital (BWH). Neurologists at MGH and BWH selected diagnostically complex cases with multiple differential diagnoses. For each case, DIAGNO was provided only with the initial presentation and history of present illness to generate differential diagnoses and accompanying clinical reasoning, without access to the subsequent diagnostic workup or final diagnosis. The neurologists’ differentials were independently extracted from the health record.

Diagnostic performance was evaluated against the final diagnosis established after completion of the diagnostic workup. Two of the eight cases had autopsy-confirmed neuropathological diagnoses. For the remaining six cases, the reference standard was the final clinical diagnosis documented in the medical record, incorporating subsequent diagnostic testing when applicable, and independently reviewed and confirmed by two neurologists (JRD and DR). Thus, these six cases used an adjudicated clinical reference standard rather than a neuropathological diagnosis.

Expert neurologists then evaluated DIAGNO’s output using a 5-point Likert scale across five dimensions: (1) accuracy of content, (2) completeness, (3) appropriateness of follow-up testing suggestions, (4) clinical reasoning, and (5) absence of harm. This evaluation was designed to capture qualitative aspects of diagnostic utility beyond diagnostic concordance, providing an assessment of how the generated differentials and reasoning would be received by practicing neurologists.

## Results

### NeuroBench

NeuroBench encompassed a wide range of conditions, including hydrocephalus, Alzheimer’s disease, CVD, tumors and neoplasms, and amyotrophic lateral sclerosis (ALS), with diagnostic distributions varying across sites (Figure 2A, B, and C). CVD and tumors were the most frequent categories at MGH, whereas hydrocephalus and ALS predominated at Mount Sinai and Alzheimer’s disease predominated at UTHSA. As final diagnoses were recorded as free text, we applied semantic harmonization to consolidate synonymous terms, yielding 59 unique standardized NeuroBench diagnoses, and grouped into 31 taxonomical categories (Table S1).

**Figure 2.**
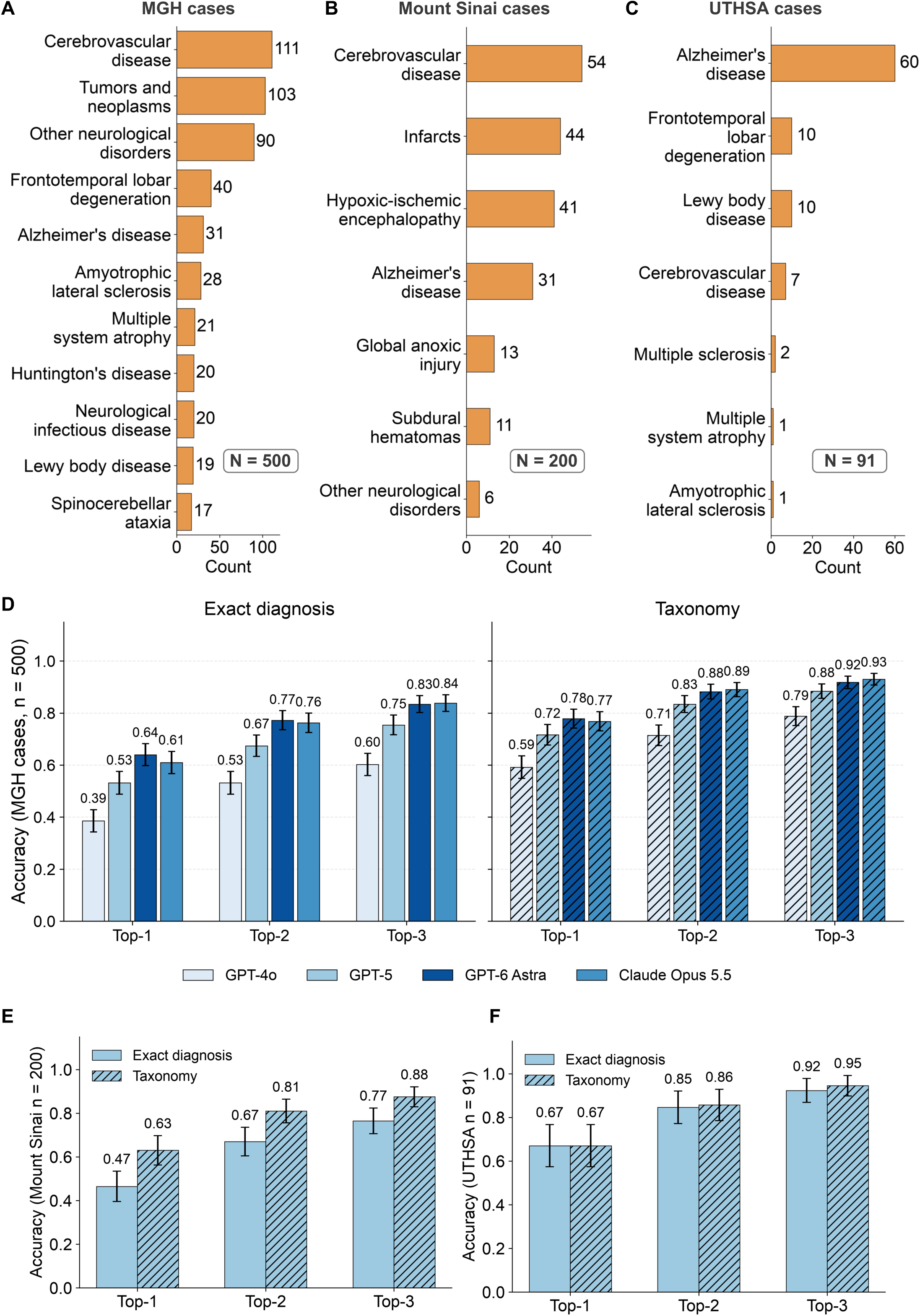
DIAGNO Performance. **(A) Distribution of MGH cases.** Bar plot showing the number of cases in each major neuropathologic diagnostic taxonomy; categories with fewer than 15 cases were grouped as “Other neurological disorders.” **(B) Distribution of Mount Sinai cases. (C) Distribution of UTHSA cases. (D) DIAGNO Performance across language models in the MGH cases.** Top-1, top-2, and top-3 accuracy at the exact-diagnosis and taxonomy levels for GPT-4o, GPT-5, GPT-6 Astra, and Claude Opus 5.5. **(E) External evaluation at Mount Sinai.** Top-1, top-2, and top-3 exact-diagnosis and taxonomy accuracy using GPT-5.6 Sol. **(F) External evaluation at UTHSA.** Top-1, top-2, and top-3 exact-diagnosis and taxonomy accuracy using GPT-5.

### Model benchmarking

DIAGNO’s diagnostic accuracy improved with more capable backbone models. On the 500 MGH cases, exact top-3 accuracy rose from 60.2% with GPT-4o to 75.4% with GPT-5 and 83.4% with GPT-6 Astra, with the same ordering at top-1 (38.6%, 53.2%, and 64.0%) and top-2 (53.2%, 67.4%, and 77.2%) (Figure 2D). DIAGNO implemented with Claude Opus 5.5 achieved 61.0%, 76.2%, and 83.8% top-1, top-2, and top-3 accuracy, respectively. Taxonomy-level accuracy exceeded exact accuracy (top-3: 78.8% with GPT-4o, 88.4% with GPT-5, and 91.8% with GPT-6 Astra); however, the performance difference between the two narrowed as the backbone improved (18.6, 13.0, and 8.4 percentage points, respectively), indicating that stronger models derived less incremental benefit from taxonomy-level scoring. Stronger models also made more confident assessments: the proportion of cases routed to Stage 2 retrieval fell from 84.8% (424/500) with GPT-4o to 81.8% (409/500) with GPT-5 and 66.0% (330/500) with GPT-6 Astra. Nonetheless, for every backbone, cases resolved at Stage 1 without retrieval were diagnosed more accurately than cases routed to Stage 2 (exact top-3: 84.2% vs. 55.9% for GPT-4o, 94.5% vs. 71.2% for GPT-5, and 98.2% vs. 75.8% for GPT-6 Astra), suggesting that confidence self-assessment separates difficult cases across model backbones (Table S3).

Several of these trends persisted at Mount Sinai and UTHSA. At Mount Sinai, DIAGNO implemented with GPT-5.6 Sol achieved 46.5% top-1, 67.0% top-2, and 76.5% top-3 accuracy across 200 cases, and 63.0%, 81.0%, and 87.5% at the taxonomy level (Figure 2E). At UTHSA, DIAGNO implemented with GPT-5 achieved 67.0%, 84.6%, and 92.3% exact accuracy across 91 cases, and 67.0%, 85.7%, and 94.5% at the taxonomy level (Figure 2F). Absolute accuracy differed across sites, consistent with differences in case mix, summary length, and the process by which summaries were generated, but the ordering of top-1 through top-3 accuracy, the advantage of taxonomy-level over exact scoring, and the performance between high confidence and low or medium-confidence cases held across all three sites.

### Diagnostic accuracy

We next compared differential diagnoses generated by DIAGNO on 203 complex NeuroBench cases at MGH versus those produced by expert human neurologists. As expected, absolute diagnostic accuracy was modest across all methods and adjudicators (Figure 3A, Supplementary Figure 1), likely reflecting both case complexity and the conference format where imaging and diagnostic test details were intentionally withheld.

**Figure 3.**
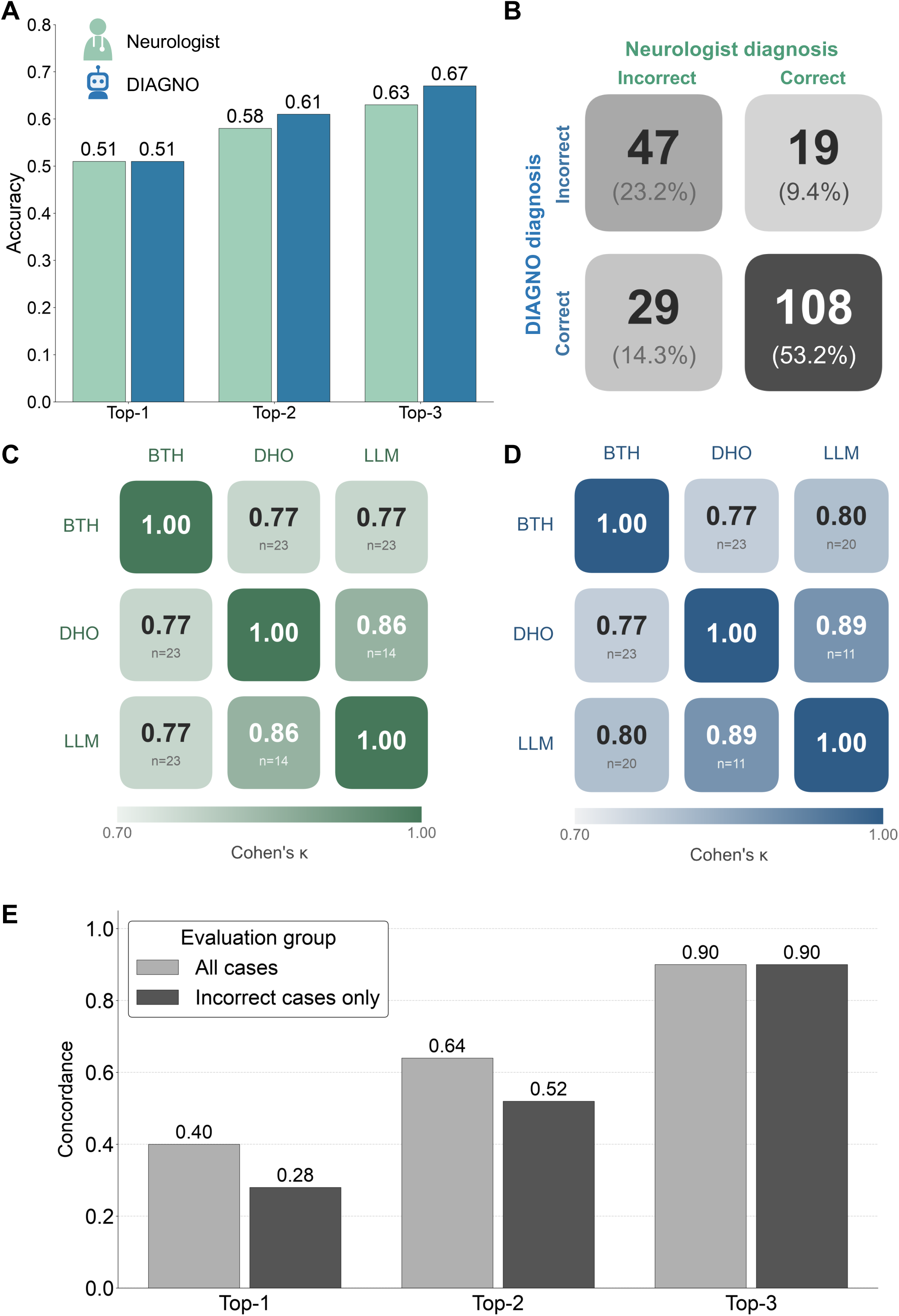
Comparative evaluation of neurological diagnoses by DIAGNO and neurologists using neuropathological reference standards in the 203 complex MGH cases. **(A) Overall diagnostic accuracy of neurologists and DIAGNO.** Top-1, top-2, and top-3 diagnostic accuracy of neurologists and DIAGNO, based on adjudication by two human experts (BTH and DHO) and an LLM (GPT-4o) against neuropathological reference-standard diagnoses. **(B) Comparison of DIAGNO diagnoses and neurologist diagnostic correctness.** Distribution of cases according to the correctness of the top-3 diagnoses from DIAGNO and neurologists. Each cell shows the number and percentage of cases in which both were correct, only the neurologist was correct, only DIAGNO was correct, or both were incorrect. **(C-D) Inter-adjudicator agreement on diagnostic correctness.** Pairwise agreement among BTH, DHO, and the LLM for evaluating the correctness of **(C)** neurologist and **(D)** DIAGNO diagnoses. Cells show Cohen’s κ and the number of discordant cases (*n*) between each pair of adjudicators. (E) Concordance between DIAGNO and neurologists differential diagnoses across top-*k* ranks. Concordance between DIAGNO and neurologist differential diagnoses at top-1, top-2, and top-3, evaluated across all cases (n=203) and separately among cases in which both DIAGNO and neurologists were incorrect (n=29). Top-3 concordance was 0.90 in both groups, indicating substantial overlap between the differential diagnoses generated by DIAGNO and neurologists even when neither included the neuropathological reference diagnosis.

Nevertheless, DIAGNO demonstrated diagnostic accuracy comparable to, and in some analyses higher than, that of expert neurologists. Using an overall accuracy measure where cases were considered correct if any of the three adjudicators marked them as such, DIAGNO achieved equivalent top-1 accuracy (0.51 [95% CI, 0.44–0.58]) and demonstrated higher accuracy for top-2 (0.61 [95% CI, 0.54–0.68] vs. 0.58 [95% CI, 0.51–0.64]) and top-3 (0.67 [95% CI, 0.61–0.74] vs. 0.63 [95% CI, 0.56–0.69]) differentials (Figure 3A). For top-3 accuracy, DIAGNO achieved 0.67 compared with 0.63 for neurologists, corresponding to a paired accuracy difference of 0.05 (95% CI, -0.01 to 0.12). Neurologists and DIAGNO reached the same top-3 diagnostic conclusion in 76.4% of cases, with both correct in 53.2% (*n*=108) and both incorrect in 23.2% (*n*=47) of cases (Figure 3B). In the remaining 48 cases, DIAGNO was correct more often than neurologists (*n*=29 vs. *n*=19), suggesting that DIAGNO may capture diagnostic possibilities that clinicians overlook. Fifteen illustrative case examples each are provided for the following—both correct (Supplementary Note 4), DIAGNO correct (Supplementary Note 5), and neurologist correct (Supplementary Note 6).

Assessments by each individual adjudicator showed a similar pattern, with DIAGNO achieving slightly higher top-1 accuracy across all adjudicators (LLM adjudicator: DIAGNO 0.42 [95% CI, 0.36–0.49] vs. neurologists 0.41 [95% CI, 0.34–0.48]; BTH: DIAGNO 0.48 [95% CI, 0.41–0.55] vs. neurologists 0.46 [95% CI, 0.40–0.53]; DHO: DIAGNO 0.41 [95% CI, 0.34–0.48] vs. neurologists 0.40 [95% CI, 0.33–0.47]). This performance gap increased for top-2 and top-3 differentials, with DIAGNO achieving significantly higher top-3 accuracy than neurologists under adjudicator DHO (McNemar’s test, *p* = 0.034), although this difference did not remain statistically significant after correction for multiple comparisons. A similar trend was observed under the LLM adjudicator, although the difference did not reach statistical significance (*p* = 0.087). This finding aligns with previous research showing that LLMs can help clinicians broaden their differential diagnoses^11^, potentially encompassing rare diseases that clinicians may overlook^35^.

In external validation on 40 challenging neurological case reports from NEJM, DIAGNO achieved slightly lower top-1 accuracy (0.25 [95% CI, 0.14–0.40]) but comparable top-3 accuracy (0.65 [95% CI, 0.50–0.78]; Table S4), demonstrating consistency with the primary results.

### Inter-adjudicator agreement and diagnostic concordance

Neuropathological diagnoses are complex and nuanced, with diagnostic terms varying in specificity and reflecting overlapping entities or hierarchical relationships (*e.g.*, disease subtypes or broader categories), potentially causing interpretive variability across adjudicators. To quantify adjudication reliability, we computed pairwise Cohen’s kappa across two human adjudicators and a LLM adjudicator^36^ for agreement on diagnostic correctness, separately for DIAGNO and neurologist outputs (Figure 3C–D). Agreement was substantial in all comparisons, ranging from 0.77–0.89 for DIAGNO and 0.77–0.86 for neurologists, which may reflect DIAGNO’s more consistent terminology reducing interpretive ambiguity^37^.

Diagnostic concordance between DIAGNO and neurologists was 0.40 for top-1 diagnoses, 0.64 for top-2, and 0.90 for top-3 (Figure 3E). The high top-3 concordance score suggests that although DIAGNO and neurologists may differ in ranking, they frequently identify a similar set of plausible diagnoses, reflecting strong alignment in diagnostic reasoning and complementary decision-support potential. In 47 cases where both DIAGNO and neurologists were incorrect, the top-1, top-2, and top-3 diagnosis concordance between DIAGNO and neurologists were 0.28, 0.52, and a striking 0.90, respectively, closely mirroring the alignment observed in the full dataset (Figure 3E). While top-1 concordance dropped, reflecting increased uncertainty, the near-perfect top-3 concordance indicates DIAGNO and human experts’ convergence on a similar set of plausible differentials in these cases. Despite initial concerns that neurologists’ domain familiarity might bias their errors, DIAGNO showed similar error patterns, suggesting that these cases may reflect intrinsic diagnostic ambiguity rather than solely domain-specific bias.

### Performance across diagnostic categories

Given that exact neuropathological labels can be highly granular (*e.g.*, frontotemporal lobar degeneration (FTLD) pathological subtypes), we complemented exact-match accuracy with taxonomy-based evaluation, grouping diagnoses into 31 clinically meaningful categories (Table S1, Table S5). Taxonomy-level evaluation yielded higher top-1, top-2, and top-3 accuracies for both DIAGNO and neurologists (Supplementary Figure 1D). Notably, DIAGNO achieved 0.74 [95% CI, 0.68–0.80] top-3 taxonomy accuracy, compared with 0.67 [95% CI, 0.59–0.72] for neurologists.

Diagnostic accuracy varied significantly across the five major diagnostic categories for both DIAGNO (Pearson’s χ²=44.14, *p*<0.0001) and neurologists (χ²=33.56, *p*<0.0001). Both performed best in neurodegenerative diseases and less well in infectious or inflammatory disorders and tumors or neoplasms (Supplementary Figure 2). Standardized residuals indicating which categories contributed most to these deviations are provided in Table S6.

DIAGNO demonstrated higher accuracy in neurodegenerative diseases (0.90 [95% CI, 0.78–0.96] vs. 0.78 [95% CI, 0.64–0.87]), infectious/inflammatory disorders (0.43 [95% CI, 0.21–0.67] vs 0.29 [95% CI, 0.12–0.55]) and other neurological conditions (0.56 [95% CI, 0.33–0.77] vs 0.31 [95% CI, 0.14–0.56]), whereas neurologists showed higher accuracy in CVD (0.53 [95% CI, 0.37–0.67] vs 0.48 [95% CI, 0.33–0.63]) and tumors/neoplasms (0.56 [95% CI, 0.41–0.70] vs 0.47 [95% CI, 0.33–0.61]). However, none of the pairwise comparisons between DIAGNO and neurologists were statistically significant (McNemar’s test, all *p*>0.05). Complete 31-taxonomy subgroup results are in Table S7.

### Error analysis

Among the 47 cases misclassified by both DIAGNO and the neurologists, 29 remained incorrect even under the more lenient taxonomy-based evaluation (fifteen illustrative examples in Supplementary Note 7). The majority of these 29 cases were brain tumors (*n*=8) or CVD (*n*=8) (Figure 4A). Other neurological disorders included demyelinating diseases (*e.g.*, multiple sclerosis, leukoencephalopathy), neurodegenerative diseases (*e.g.*, Alzheimer’s disease, Huntington’s disease, Wolfram syndrome), traumatic or structural injury (*e.g.*, white matter and axonal injury, superficial siderosis) (Figure 4A).

**Figure 4.**
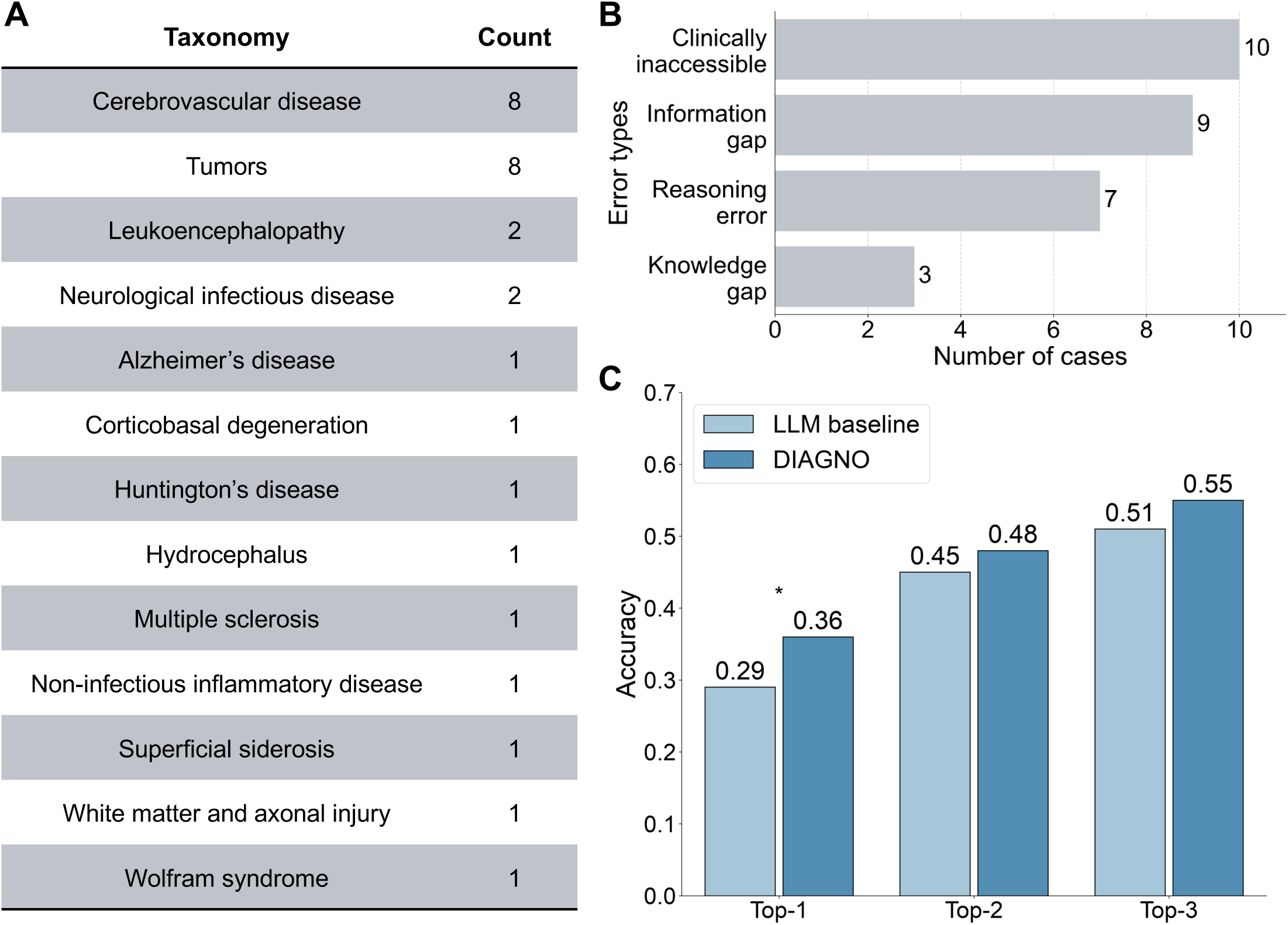
Error analysis of incorrect diagnoses and sensitivity analysis of DIAGNO performance in 203 complex MGH cases. **(A) Distribution of diagnostic taxonomies among incorrect cases.** Among the 29 cases in which both DIAGNO and neurologists were incorrect even at the taxonomy level, the most frequent diagnostic categories were cerebrovascular disease and tumors (n=8 each). **(B) Error typology among consensus error cases.** Distribution of 29 consensus error cases across four categories: clinically inaccessible (*n*=10), where the diagnosis required autopsy findings not inferable from the available clinical data; information gap (*n*=9), where essential clinical information was withheld from the case summary; reasoning error (*n*=7), where available clues were misinterpreted or incorrectly weighted; and knowledge gap (*n*=3), involving rare or atypical presentations that were not recognized. Approximately two-thirds (19/29; 66%) of errors were attributed to limitations in clinically available information rather than reasoning or knowledge errors. **(C) Sensitivity analysis of DIAGNO performance versus the LLM baseline.** Diagnostic accuracy of DIAGNO and the baseline LLM inference model used in Stage 1, restricted to the 150 cases classified as medium or low confidence during initial inference. DIAGNO showed higher accuracy at top-1 (0.36 vs. 0.29), top-2 (0.48 vs. 0.45), and top-3 (0.55 vs. 0.51), with the difference reaching statistical significance only for top-1 (McNemar’s test, *P*=0.029). * indicates *P*<0.05.

To investigate the underlying sources of error, we conducted a qualitative typology analysis by reviewing each case summary, diagnostic output, and autopsy report. We classified errors into four categories (Figure 4B): (1) clinically inaccessible (*n*=10), where decisive diagnostic evidence was only available through postmortem examination; (2) information gaps (*n*=9), where relevant diagnostic information may have been intentionally omitted in the case summaries for educational purposes, limiting the information available for diagnosis; (3) diagnostic reasoning errors (*n*=7), where the correct diagnosis was theoretically attainable but missed due to misinterpretation or suboptimal weighting of available findings; and (4) knowledge gaps (*n*=3), where rare or atypical diseases were not recognized despite adequate diagnostic clues. For the 10 clinically inaccessible cases, the final diagnosis was not evident in the clinical course and could only be confirmed by neuropathological findings identified during autopsy. For example, a patient with no cognitive symptoms on neurologic exam 9 months before death was ultimately diagnosed with Alzheimer’s Disease Neuropathologic Change (ADNC)^23^, with neocortical neuritic plaques consistent with a moderate CERAD^38^ (Consortium to Establish a Registry for Alzheimer’s Disease) score. Such cases demonstrate resilience, defined as ability to maintain their cognitive abilities despite the presence of ADNC^39^, and while blood or CSF biomarkers can estimate amyloid pathology antemortem^40^, the definitive assessment of plaque burden and CERAD scoring requires autopsy-based neuropathological examination^23,41^. Detailed case summaries, DIAGNO and neurologist differential diagnoses, and autopsy-confirmed reference-standards for these 10 cases are provided in Supplementary Note 7 (Examples 1-10).

Additionally, for the 16 cases where DIAGNO incorrectly diagnosed tumors or CVD, we evaluated DIAGNO’s capability to recommend appropriate next-step diagnostic tests, including an assessment of urgency. MRI was suggested as one of the top two tests in all 16 cases and assigned high urgency to 15 of them (Table S8), demonstrating DIAGNO’s ability to recognize situations requiring neuroimaging consistent with standard neurological practice.

Next, we specifically examined performance in a subset of 150 cases from MGH with medium or low initial confidence, where Stage 2 retrieval is activated. In this subset, DIAGNO demonstrated consistent gains over non-RAG-augmented LLM inference in Stage 1 (Figure 4C). Top-1 accuracy improved significantly from 0.29 to 0.36 (McNemar’s test: χ²=4.76, *p*=0.029). Improvements were also observed in top-2 (0.45 to 0.48) and top-3 (0.51 to 0.55) accuracy, although these did not reach statistical significance. Applying Stage 2 retrieval to all cases, including those initially assigned high confidence, did not improve diagnostic performance, supporting the efficiency of selectively applying RAG to medium- and low-confidence cases (Supplementary Figure 3). In addition, majority voting produced modest but consistent improvements in top-k diagnostic accuracy compared with a single RAG inference, whereas taxonomy-level accuracy remained largely unchanged (Supplementary Figure 4).

### Real-world evaluation

Across eight real-world cases from MGH and BWH, DIAGNO was provided with clinical information available at the initial presentation and tasked with generating a differential diagnosis and clinical reasoning. Expert neurologists (JRD and DR) rated DIAGNO favorably overall, with a mean rating of 4.03 out of 5 across the five evaluation dimensions (Figure 5A). Mean ratings were highest for absence of harm (4.75/5), while clinical reasoning received the lowest mean rating (3.38/5); ratings varied across individual cases, consistent with heterogeneity in the clinical presentations (Figure 5B).

**Figure 5.**
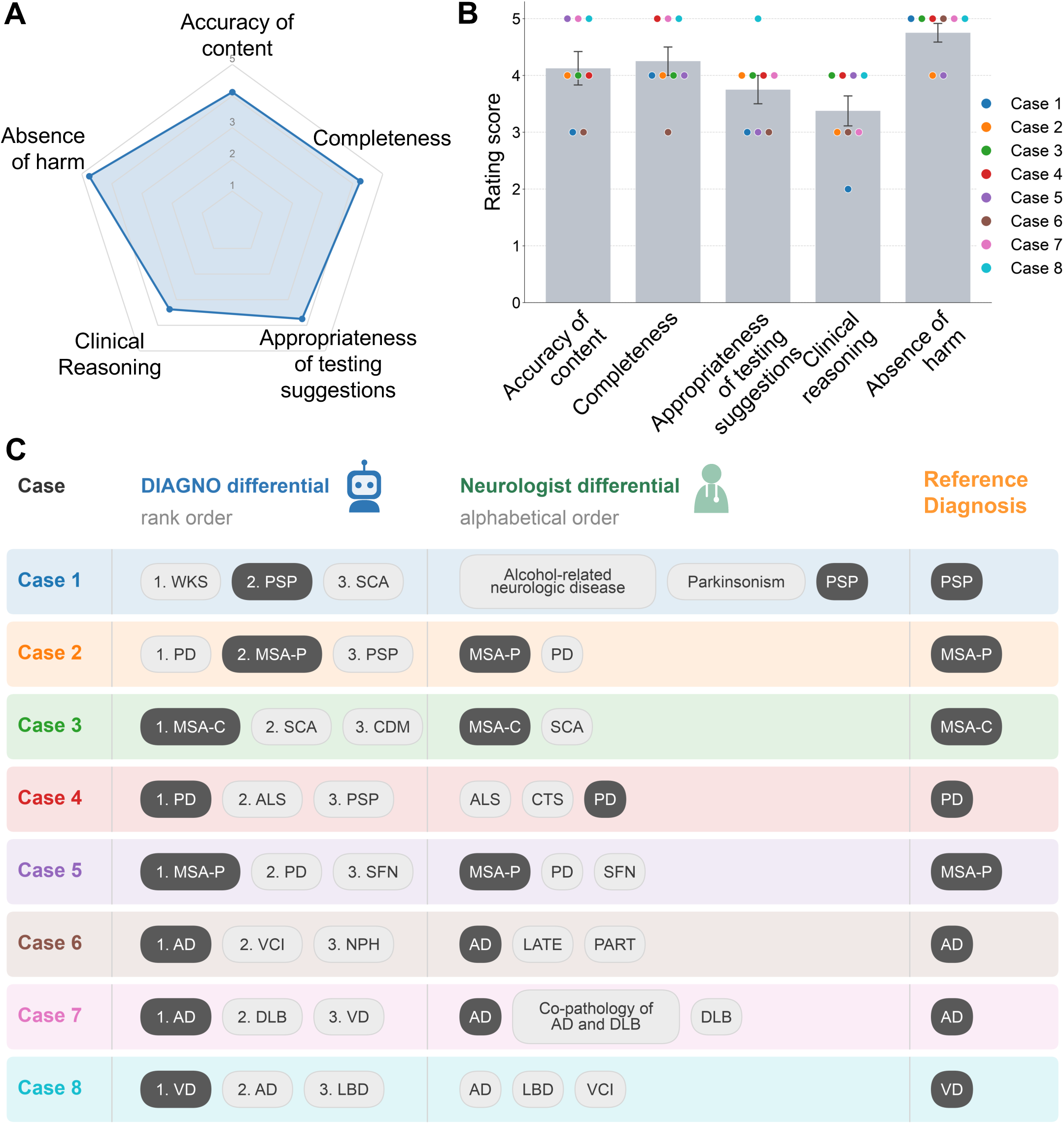
Expert-rated performance of DIAGNO across five clinical dimensions. **(A) Radar plot summarizing mean neurologist ratings of DIAGNO.** Neurologists rated DIAGNO across five dimensions: accuracy of content, completeness, appropriateness of follow-up testing suggestions, clinical reasoning, and absence of harm, using a 5-point Likert scale. **(B) Evaluation across eight real-world cases from Mass General Brigham healthcare.** Bar plots show mean neurologist ratings for each dimension, with points representing individual cases. **(C) DIAGNO and neurologist differentials versus reference diagnosis.** For each case, the top-3 differential diagnoses generated by DIAGNO are shown alongside the neurologists’ differential diagnoses documented at the initial clinical evaluation (unranked and displayed alphabetically) and the reference diagnosis established after full diagnostic workup. Abbreviations: AD, Alzheimer’s disease; ALS, amyotrophic lateral sclerosis; CDM, corticobasal degeneration with motor neuron disease; CTS, carpal tunnel syndrome; DLB, dementia with Lewy bodies; LATE, limbic-predominant age-related TDP-43 encephalopathy; LBD, Lewy body disease; MSA-C, multiple system atrophy-cerebellar type; MSA-P, multiple system atrophy-parkinsonian type; NPH, normal pressure hydrocephalus; PART, primary age-related tauopathy; PD, Parkinson’s disease; PSP, progressive supranuclear palsy; SCA, spinocerebellar ataxia; SFN, small fiber neuropathy; VCI, vascular cognitive impairment; VD, vascular dementia; WKS, Wernicke-Korsakoff syndrome.

Notably, the reference diagnosis established after completion of the diagnostic workup was included among DIAGNO’s top-three differential diagnoses in all eight cases and was ranked first in six of eight cases (Figure 5C). Two of these reference diagnoses were subsequently confirmed by neuropathological examination; the remaining six represented adjudicated final clinical diagnoses. In one case, DIAGNO included the final diagnosis in its initial differential when the neurologist’s documented initial differential did not, indicating that DIAGNO identified a clinically relevant diagnostic possibility not documented by the neurologists at initial presentation. DIAGNO also received consistently high ratings for absence of harm across cases (mean rating of 4.75/5), indicating that experts rarely identified potentially harmful reasoning or recommendations in this eight-case evaluation.

## Discussion

### Main findings and clinical significance

This study introduces NeuroBench, a benchmark of 791 autopsy-confirmed neurological cases spanning 59 unique diagnoses, including 203 complex cases, and uses it to evaluate both DIAGNO and board-certified neurologists against neuropathological ground truth. This design enables direct measurement of diagnostic accuracy anchored to definitive diagnoses rather than clinical consensus. DIAGNO showed comparable top-3 performance to neurologists, with higher exact-match accuracy (0.67 versus 0.63) and taxonomy-level accuracy (0.74 versus 0.67). Importantly, among the 48 cases with discordant top-3 correctness, DIAGNO alone was correct in 29 cases compared with 19 cases in which neurologists alone were correct, suggesting that DIAGNO may identify diagnostic possibilities not captured in clinicians’ initial differentials. In a real-world evaluation, neurologists rated DIAGNO favorably (mean 4.03/5) across multiple dimensions of clinical utility and absence of harm, further supporting its potential applicability as a clinical decision-support tool beyond retrospective benchmarking.

### Strengths

This study offers several strengths enhancing its scientific rigor and clinical relevance. First, evaluating against autopsy-confirmed neuropathological diagnoses, the gold standard in neurological diagnosis, fills a critical methodological gap unaddressed in most prior AI-diagnostic studies. Second, the design incorporates blinded adjudication, direct comparison with board-certified neurologists, and an unusually heterogeneous set of 59 distinct diagnoses, strengthening internal validity and mirroring real-world diagnostic complexity. Third, DIAGNO features confidence-aware output, detailed rationale, and dual-stage retrieval-augmented generation (RAG). Its explicit confidence ratings and structured reasoning summaries allow clinicians to understand how conclusions were reached, essential for interpretability and trust^42^. Stage 2 refinement yielded significant improvement in top-1 accuracy among lower-confidence cases (from 0.29 to 0.36; *p*=0.029), supporting the value of DIAGNO’s confidence-aware, retrieval-augmented architecture in improving performance for uncertain cases. Fourth, external evaluation across MGH, Mount Sinai, and UTHSA demonstrated performance across cohorts with different case populations and diagnostic distributions. DIAGNO achieved a top-3 accuracy of 83.8% in the MGH cases, 76.5% in the Mount Sinai cases, and 92.3% in the UTHSA cases. Variation in performance may refelct differences in dataset composition and diagnostic complexity across sites. The MGH dataset included broader range of neurological and neuropathological diagnoses, including diverse rare and diagnostically challenging cases. The Mount Sinai dataset was enriched for cerebrovascular and hypoxic-ischemic conditions, which can be challenging because of less distinctive clinical presentations of symptoms. The UTHSA dataset was smaller and predominantly composed of neurodegenerative diseases, which often presented with characteristic neurological symptoms that provided more direct diagnostic clues. Nevertheless, the consistently high top-3 performance across cohorts with substantially different case mixes supports the generalizability of DIAGNO’s diagnostic reasoning. Additionally, external validation on 40 challenging *NEJM* cases demonstrates robustness, with top-3 accuracy (0.65) comparable to the complex MGH dataset (0.67). Lower top-1 accuracy likely reflects the exceptional difficulty of published cases and reliance on text-only narratives without multimodal diagnostic data. Despite these constraints, preserved top-3 performance supports the generalizability of DIAGNO’s diagnostic reasoning across datasets. Finally, real-world evaluation at MGH and BWH demonstrated clinical acceptability and safety, with neurologists rating DIAGNO favorably across five dimensions of diagnostic quality and particularly highly on absence of harm (4.75/5).

### Potential applications in medical training and clinical decision support

DIAGNO’s strong alignment with expert reasoning patterns, reflected in high diagnostic concordance even when both human and AI were incorrect, points to substantial educational benefit. This concordance, further supported by error-typology findings that most consensus errors arose from information limitations rather than model-specific failures, implies that DIAGNO can serve as a tool for illustrating expert reasoning pathways. By providing structured outputs with rationale and confidence levels, DIAGNO may aid trainees in recognizing diagnostic uncertainty, broadening differentials, and adopting systematic evidence-guided approaches. This pattern of human-AI convergence has been previously observed: an AI-generated case presentation was published alongside analyses by a master clinician ^43^, demonstrating that such tools can articulate reasoning comparable to experienced diagnosticians.

Beyond training, DIAGNO could potentially support clinical workflows including case-conference preparation by identifying ambiguous language, missed diagnostic cues, or insufficiently developed differentials; second-opinion support for community neurologists; and augmenting diagnostic reasoning in challenging evaluations. Notably, DIAGNO reliably provided appropriate follow-up neuroimaging in cases involving missed diagnoses of tumors or CVD, highlighting potential value even when the primary diagnosis is incorrect. These capabilities point toward hybrid human–AI workflows that broaden differential diagnoses, support uncertainty-aware reasoning, and flag situations requiring urgent diagnostic testing ^33,44^.

### Limitations and future directions

While DIAGNO’s performance highlights its potential as both a diagnostic support and training tool for complex neurological cases, there are some limitations that could be further addressed in future works. First, the retrospective design using case summaries rather than full longitudinal records limits ecological validity; key clinical features may have been omitted to preserve patient privacy. Second, while this study focused on text-based clinical notes with key diagnostic test results withheld, incorporating richer multimodal data (e.g., imaging, biomarkers) could enhance diagnostic accuracy^45^. Third, the real-world evaluation included only eight cases from MGB, was based on retrospective expert assessment, and did not evaluate downstream clinical outcomes (*e.g.*, reduced diagnostic delay, improved patient management). Larger prospective studies are needed to evaluate clinical utility and safety. Finally, when comparing DIAGNO performance with that of expert neurologists, GPT-4o served as both the backbone model for DIAGNO and one of the three adjudicators, which may introduce potential shared model bias. However, results were also evaluated independently by two board-certified human adjudicators, with performance trends consistent across the human and GPT-4o adjudicators.

## Conclusions

In summary, this work introduces a neuropathologically grounded framework for assessing LLM diagnostic reasoning in complex neurological diseases. NeuroBench provides a reusable benchmark anchored to neuropathological ground truth, and DIAGNO demonstrates diagnostic performance comparable to expert neurologists in complex, autopsy-confirmed neurological cases, while identifying additional diagnostic possibilities in some cases, as well as providing diagnostic reasoning and confidence estimates. These findings suggest that AI-augmented diagnostic systems can assist both clinicians and trainees in decision-making in neurology. However, future work is essential to further validate DIAGNO across diverse institutions, settings, and populations—including community and lower-resource settings and varying levels of clinical documentation—integrate multimodal data, and demonstrate clinical impact before deployment in routine care.

## Data Availability

Of the 791 cases in NeuroBench, the 203 complex cases, including deidentified case summaries and corresponding neuropathologically confirmed diagnoses, are publicly available at https://github.com/mindds/DIAGNO.

## Acknowledgements

We acknowledge funding from the following source: NIH R01AG082698, NIH P30AG062421, NIH P30AG066546, the JMR Barker Foundation, and the Bill and Rebecca Reed Endowment for Precision Therapies and Supportive Care. A.N. was supported by the Rhodes Scholarship. D.A.C. was funded by an NIHR Research Professorship and an RAEng Research Chair, and supported by the Oxford NIHR Biomedical Research Centre. The authors would like to thank Mass General Brigham research computing and information technology for supporting this study.

## Data Availability

The complete NeuroBench dataset, comprising 203 deidentified case summaries and corresponding neuropathologically confirmed diagnoses, is available at https://github.com/mindds/NeuroBench upon request.

## Code Availability

The code developed for this study is available at https://github.com/mindds/DIAGNO.

## Notes

### Competing Interest Statement

The authors have declared no competing interest.

### Author Declarations

Institutional Review Board of Mass General Brigham gave ethical approval for this work.

### Summary of Updates

The major revisions include: 1.Expanded NeuroBench to a multi-institutional benchmark. We increased the dataset from 203 cases at a single institution to 791 autopsy-confirmed neurological cases across three independent health systems: Massachusetts General Hospital (n=500), Mount Sinai Health System (n=200), and UTHSA (n=91). 2.Expanded benchmarking across frontier LLMs. We evaluated the DIAGNO framework using multiple frontier language models including GPT-6 Astra and Claude Opus 5.5, enabling assessment of how diagnostic performance and confidence-guided retrieval vary across model backbones. 3.Clarified the comparison with neurologists and reference standards. 4.Added methodological and ablation analyses. 5.Strengthened statistical reporting and presentation.

