## Supplementary Material for "Assessing AI and Neurologist Diagnostic Reasoning Against Neuropathological Ground Truth"

**Supplementary Appendix**

**This appendix has been provided by the authors to give readers additional information about their work.**

### Supplementary Tables

Table S1. Mapping of Neuropathological Diagnoses to Broader Taxonomy-Based Disease Categories

| **Diagnosis** | **Taxonomy** |
| --- | --- |
| Alzheimer's Disease | Alzheimer's Disease |
| Amyotrophic Lateral Sclerosis (ALS) | Amyotrophic Lateral Sclerosis (ALS) |
| Global Anoxic Injury | Anoxic Injury |
| Corticobasal Degeneration (CBD) | Corticobasal Degeneration (CBD) |
| Down Syndrome (DS) | Down Syndrome (DS) |
| Frontotemporal Lobar Degeneration (FTLD) | Frontotemporal Lobar Degeneration (FTLD) |
| Hereditary Sensory & Autonomic Neuropathy (HSAN) Type I | Hereditary Sensory & Autonomic Neuropathy (HSAN) |
| Huntington's Disease | Huntington's Disease |
| Hydranencephaly | Hydranencephaly |
| Hydrocephalus | Hydrocephalus |
| Leukoencephalopathy | Leukoencephalopathy |
| Lewy Body Disease (LBD/DLB) | Lewy Body Disease (LBD/DLB) |
| Mitochondrial Encephalopathy, Lactic Acidosis, and Stroke-Like Symptoms (MELAS) | MELAS |
| Motor Neuron Disease | Motor Neuron Disease |
| Multiple Sclerosis | Multiple Sclerosis |
| Multiple System Atrophy (MSA) | Multiple System Atrophy (MSA) |
| Neuronal Ceroid Lipofuscinosis Type 2 (CLN2) | Neuronal Ceroid Lipofuscinosis (NCL) |
| Neurofibromatosis Type 1 | Neurofibromatosis |
| Neurofibromatosis Type 2 | Neurofibromatosis |
| Granulomatous Ependymitis of Unknown Cause | Non-infectious Inflammatory Disease |
| Demyelinating myelopathy (myelitis) | Non-infectious Inflammatory Disease |
| Unilateral Destructive Inflammatory Process | Non-infectious Inflammatory Disease |
| Paraneoplastic Cerebellar Degeneration (PCD) | Paraneoplastic Cerebellar Degeneration (PCD) |
| Parkinson's Disease (PD) | Parkinson's Disease (PD) |
| Primary Lateral Sclerosis (PLS) | Primary Lateral Sclerosis (PLS) |
| Progressive Supranuclear Palsy (PSP) | Progressive Supranuclear Palsy (PSP) |
| Spinal Muscular Atrophy (SMA) | Spinal Muscular Atrophy (SMA) |
| Spinocerebellar Ataxia (SCA) | Spinocerebellar Ataxia (SCA) |
| Superficial Siderosis | Superficial Siderosis |
| White Matter and Axonal Injury | White Matter and Axonal Injury |
| Wolfram Syndrome | Wolfram Syndrome |
| Hypoxic-ischemic encephalopathy | Cerebrovascular Disease (CVD) |
| Basilar Artery Occlusion (BAO) | Cerebrovascular Disease (CVD) |
| Cerebellar Hemorrhage | Cerebrovascular Disease (CVD) |
| Cerebral Amyloid Angiopathy (CAA) | Cerebrovascular Disease (CVD) |
| Cerebrovascular Disease (CVD) | Cerebrovascular Disease (CVD) |
| Intracranial hemorrhage | Cerebrovascular Disease (CVD) |
| Infarcts (INF) | Cerebrovascular Disease (CVD) |
| Intracerebral Hemorrhage | Cerebrovascular Disease (CVD) |
| Subarachnoid Hemorrhage (SAH) | Cerebrovascular Disease (CVD) |
| Subdural Hematomas | Cerebrovascular Disease (CVD) |
| Adenovirus Meningoencephalomyelitis | Neurological Infectious Disease (NID) |
| Bacterial Endocarditis | Neurological Infectious Disease (NID) |
| Invasive Fungal Cerebritis | Neurological Infectious Disease (NID) |
| Chronic Meningoencephalitis, HHV 4 Related | Neurological Infectious Disease (NID) |
| Cryptococcal Meningoencephalitis | Neurological Infectious Disease (NID) |
| Granulomatous Amebic Encephalitis (GAE) | Neurological Infectious Disease (NID) |
| Anaplastic Astrocytoma | Tumors and Neoplasms |
| Anaplastic Ependymoma | Tumors and Neoplasms |
| Anaplastic Meningioma | Tumors and Neoplasms |
| Primary CNS Lymphoma (PCNSL) | Tumors and Neoplasms |
| Glioblastoma | Tumors and Neoplasms |
| Leptomeningeal Carcinomatosis (LMC) | Tumors and Neoplasms |
| Macroinvasive Pituitary Adenoma | Tumors and Neoplasms |
| Metastatic Carcinoma | Tumors and Neoplasms |
| Midline Glioma | Tumors and Neoplasms |
| Multifocal Atypical Meningioma | Tumors and Neoplasms |
| Oligodendroglioma | Tumors and Neoplasms |
| Pilocytic Astrocytoma | Tumors and Neoplasms |

Table S2. List of NEJM Case Reports Used for External Validation of DIAGNO (N=40)

| **Case ID** | **PMID** | **Year** | **Title** |
| --- | --- | --- | --- |
| 1913477 | 32402166 | 2020 | A Man Aged 76–80 Years with Hyponatremia and Involuntary Movements of the Arm and Face |
| 1916251 | 33382934 | 2020 | A Man Aged 61–65 Years with Memory Loss and Odd Behavior |
| 2002412 | 33176089 | 2020 | A Woman Aged 56–60 Years with Type 1 Diabetes Mellitus and Obtundation |
| 2004976 | 32813954 | 2020 | A Woman Aged 61–65 with Altered Mental Status and Weakness on the Left Side |
| 2004996 | 33053289 | 2020 | A Man Aged 61–65 with Confusion, Fatigue, and Garbled Speech |
| 2027083 | 33369359 | 2020 | A Man Aged 21–25 with Headache and Covid-19 |
| 2004992 | 34233100 | 2021 | A Man Aged 66–70 with Ataxia |
| 2027080 | 34758256 | 2021 | A Man Aged 36–40 with Altered Mental Status and New Onset of Seizures |
| 2027089 | 33626257 | 2021 | A Man Aged 61–65 with Eye Pain and Decreased Vision |
| 2100272 | 34161710 | 2021 | A Man Aged 51–55 with Irritability, Confusion, and Odd Behaviors |
| 2103460 | 34289280 | 2021 | A Woman Aged 61–65 with Cognitive Impairment, Headache, and Memory Loss |
| 2107358 | 34965341 | 2021 | A Boy Aged 6–10 with Transient Weakness, Facial Droop, and Slurred Speech |
| 2115844 | 35020988 | 2022 | A Man Aged 66–70 with Motor Neuron Disease and Odd Behaviors during Sleep |
| 2115852 | 35172059 | 2022 | A Woman Aged 61–65 with Rapidly Progressive Weakness in the Right Arm and Recurrent Falls |
| 2115853 | 35263523 | 2022 | A Woman Aged 61–65 with Depression, Recurrent Falls, and Inability to Care for Herself |
| 2201235 | 36300978 | 2022 | A Girl Aged 11–15 with Redness of the Eyes |
| 2201241 | 36070713 | 2022 | A Woman Aged 21–25 with Headache and Blurred Vision |
| 2201246 | 36069875 | 2022 | A Man Aged 31–35 with Confusion, Headache, and Fever |
| 2211355 | 36416771 | 2022 | A Woman Aged 26–30 with Decreased Vision and Headache |
| 2211365 | 36988597 | 2023 | A Man Aged 26–30 with Convulsions |
| 2211368 | 36812438 | 2023 | A Man Aged 66–70 with Recurrent Strokes |
| 2211512 | 37342926 | 2023 | A Man Aged 76–80 with Left Foot Drop |
| 2300895 | 37195945 | 2023 | A Man Aged 31–35 with Paresthesia of the Arms and Legs |
| 2300973 | 37819957 | 2023 | A Man Aged 76–80 with Involuntary Movements and Unresponsiveness |
| 2309349 | 38091534 | 2023 | A Woman Aged 66–70 with Abnormal Movements and Confusion |
| 2100279 | 39774317 | 2024 | A Woman Aged 21–25 with Headache, Fever, and Respiratory Failure |
| 2309726 | 39141857 | 2024 | A Boy Aged 11–15 with Autism and Decreased Vision |
| 2312725 | 38598798 | 2024 | A Woman Aged 81–85 with Falls and Cognitive Decline |
| 2312732 | 38631006 | 2024 | A Woman Aged 56–60 with Confusion, Aphasia, and Abnormal Head Imaging |
| 2312733 | 38718362 | 2024 | A Woman Aged 26–30 with Back Pain, Leg Stiffness, and Falls |
| 2312734 | 39413380 | 2024 | A Woman Aged 71–75 with Dyspnea, Dysphagia, and Dysarthria |
| 2402484 | 38865664 | 2024 | A Woman Aged 61–65 with the Worst Headache of Her Life |
| 2402488 | 39047245 | 2024 | A Woman Aged 76–80 with Rapidly Progressive Dementia |
| 2402496 | 39442044 | 2024 | A Woman Aged 71–75 with Confusion, Aphasia, and a Brain Mass |
| 2402500 | 39602633 | 2024 | A Man Aged 41–45 with Seizures and Agitation |
| 2412511 | 39813647 | 2025 | A Man Aged 21–25 with Loss of Consciousness and a Fall |
| 2412514 | 39938097 | 2025 | A Woman Aged 26–30 with Headache and Dysesthesia |
| 2412528 | 40632952 | 2025 | A Man Aged 66–70 with Headache and Ataxia |
| 2412531 | 40742263 | 2025 | A Woman Aged 16–20 with Seizure like Activity and Odd Behaviors |
| 2513534 | 41297009 | 2025 | A Woman Aged 56–60 with Visual Disturbances and Right-Arm Shaking |

Table S3. Exact Diagnosis and Taxonomy-Level Top-K Accuracy by Model and DIAGNO Pipeline Stage

| **Model** | **Final stage** | **n** | **Exact Top-1** | **Exact Top-2** | **Exact Top-3** | **Taxonomy Top-1** | **Taxonomy Top-2** | **Taxonomy Top-3** |
| --- | --- | --- | --- | --- | --- | --- | --- | --- |
| GPT-4o | Stage 1 | 76 | 64.5% | 72.4% | 84.2% | 77.6% | 85.5% | 92.1% |
| GPT-4o | Stage 2 RAG | 424 | 34.0% | 49.8% | 55.9% | 55.9% | 68.9% | 76.4% |
| **GPT-4o** | **Total** | **500** | **38.6%** | **53.2%** | **60.2%** | **59.2%** | **71.4%** | **78.8%** |
| GPT-5 | Stage 1 | 91 | 75.8% | 87.9% | 94.5% | 79.1% | 91.2% | 96.7% |
| GPT-5 | Stage 2 RAG | 409 | 48.2% | 62.8% | 71.2% | 69.9% | 81.7% | 86.6% |
| **GPT-5** | **Total** | **500** | **53.2%** | **67.4%** | **75.4%** | **71.6%** | **83.4%** | **88.4%** |
| GPT-6 Astra | Stage 1 | 170 | 88.2% | 95.3% | 98.2% | 90.0% | 97.1% | 98.2% |
| GPT-6 Astra | Stage 2 RAG | 330 | 51.5% | 67.9% | 75.8% | 71.5% | 83.6% | 88.5% |
| **GPT-6 Astra** | **Total** | **500** | **64.0%** | **77.2%** | **83.4%** | **77.8%** | **88.2%** | **91.8%** |
| Claude Opus 5.5 | Stage 1 | 139 | 89.2% | 95.0% | 96.4% | 90.6% | 96.4% | 97.1% |
| Claude Opus 5.5 | Stage 2 RAG | 361 | 50.1% | 69.0% | 78.9% | 71.5% | 86.1% | 91.4% |
| **Claude Opus 5.5** | **Total** | **500** | **61.0%** | **76.2%** | **83.8%** | **76.8%** | **89.0%** | **93.0%** |

Table S4. Performance of DIAGNO on NEJM cases

|  | **Top 1** | **Top 2** | **Top 3** |
| --- | --- | --- | --- |
| **Diagnostic Accuracy Evaluated by GPT** | 0.25  (95% CI 0.14–0.40) | 0.40  (95% CI 0.26–0.55) | 0.65  (95% CI 0.50–0.78) |

Table S5. Detailed Case Counts per Disease Taxonomy of MGH cases (n = 500)

| **Taxonomy** | **Count** |
| --- | --- |
| Cerebrovascular Disease (CVD) | 111 |
| Tumors and Neoplasms | 103 |
| Frontotemporal Lobar Degeneration (FTLD) | 40 |
| Alzheimer's Disease | 31 |
| Amyotrophic Lateral Sclerosis (ALS) | 28 |
| Multiple System Atrophy (MSA) | 21 |
| Huntington's Disease | 20 |
| Neurological Infectious Disease (NID) | 20 |
| Lewy Body Disease (LBD/DLB) | 19 |
| Spinocerebellar Ataxia (SCA) | 17 |
| Leukoencephalopathy | 15 |
| Progressive Supranuclear Palsy (PSP) | 14 |
| Parkinson's Disease (PD) | 12 |
| Multiple Sclerosis | 11 |
| Corticobasal Degeneration (CBD) | 7 |
| MELAS | 4 |
| Neurofibromatosis | 4 |
| White Matter and Axonal Injury | 4 |
| Anoxic Injury | 2 |
| Hydrocephalus | 2 |
| Motor Neuron Disease | 2 |
| Neuronal Ceroid Lipofuscinosis (NCL) | 2 |
| Primary Lateral Sclerosis (PLS) | 2 |
| Spinal Muscular Atrophy (SMA) | 2 |
| Superficial Siderosis | 2 |
| Hereditary Sensory & Autonomic Neuropathy (HSAN) | 1 |
| Hydranencephaly | 1 |
| Non-infectious Inflammatory Disease | 1 |
| Paraneoplastic Cerebellar Degeneration (PCD) | 1 |
| Wolfram Syndrome | 1 |

Table S6. Standardized Residuals from Chi-square Analyses of Diagnostic Accuracy Across Major Disease Categories for DIAGNO and Neurologists

| **Disease Category** | **DIAGNO (Correct=1)** | **DIAGNO (Incorrect=0)** | **Neurologists (Correct=1)** | **Neurologists (Incorrect=0)** |
| --- | --- | --- | --- | --- |
| CVD | -1.18 | 1.71 | -0.82 | 1.07 |
| Infectious/Inflammatory | -1.31 | 1.90 | -1.77 | 2.30 |
| NDD | 2.81 | -4.08 | 2.41 | -3.13 |
| Other | -0.56 | 0.82 | -1.59 | 2.07 |
| Tumors/Neoplasms | -1.70 | 2.47 | -0.58 | 0.75 |

Table S7. Taxonomy-level Overall Diagnostic Accuracy for DIAGNO and Neurologists

| **Taxonomy** | **Neurologist Accuracy** | **DIAGNO Accuracy** |
| --- | --- | --- |
| Tumors and Neoplasms | 0.56 | 0.47 |
| Cerebrovascular Disease (CVD) | 0.53 | 0.53 |
| Alzheimer's Disease | 0.79 | 0.93 |
| Frontotemporal Lobar Degeneration (FTLD) | 1 | 0.92 |
| Lewy Body Disease (LBD/DLB) | 1 | 1 |
| Multiple System Atrophy (MSA) | 0.8 | 1 |
| Huntington's Disease | 0.77 | 0.89 |
| Neurological Infectious Disease (NID) | 0.44 | 0.44 |
| Amyotrophic Lateral Sclerosis (ALS) | 0.86 | 1 |
| Spinocerebellar ataxia (SCA) | 0.5 | 0.83 |
| Progressive Supranuclear Palsy (PSP) | 0.83 | 1 |
| Leukoencephalopathy | 0 | 0.25 |
| Corticobasal Degeneration (CBD) | 0.75 | 0.5 |
| Multiple Sclerosis | 0 | 0.67 |
| Non-infectious Inflammatory Disease | 0 | 0 |
| Anoxic Injury | 0.5 | 1 |
| MELAS | 1 | 1 |
| Motor Neuron Disease | 0.5 | 1 |
| Neurofibromatosis | 0.5 | 1 |
| Spinal Muscular Atrophy (SMA) | 1 | 1 |
| Down Syndrome (DS) | 0 | 1 |
| Hereditary Sensory & Autonomic Neuropathy (HSAN) | 0 | 1 |
| Hydranencephaly | 1 | 1 |
| Hydrocephalus | 0 | 0 |
| Neuronal Ceroid Lipofuscinosis (NCL) | 0 | 1 |
| Parkinson's Disease (PD) | 1 | 1 |
| Paraneoplastic Cerebellar Degeneration (PCD) | 1 | 1 |
| Primary Lateral Sclerosis (PLS) | 1 | 1 |
| Superficial Siderosis | 0 | 0 |
| White Matter and Axonal Injury | 0 | 0 |
| Wolfram Syndrome | 0 | 0 |

Table S8. Summary of GPT-4o’s Diagnostic Test Recommendations for 16 Error Cases involving Tumors/Neoplasms and Cerebrovascular Disease

| **Test Suggested** | **Frequency** | **High Urgency Rate** |
| --- | --- | --- |
| MRI | 16/16 | 0.94 |
| CT | 4/16 | 0.75 |

### Supplementary Figures

Supplementary Figure 1.

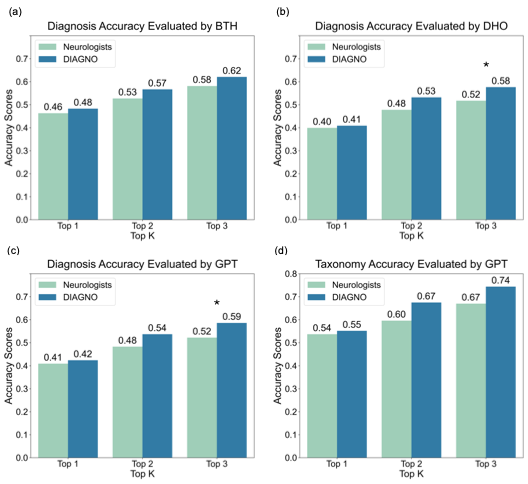

**Comparative Evaluation of Neurologist and DIAGNO Performance in Neurological Diagnosis.** (A-C) Grouped bar charts showing diagnostic accuracy of top-1, top-2, and top-3 differential diagnoses made by neurologists and DIAGNO, evaluated by three adjudicators: (A) neurologist BTH, (B) neuropathologist DHO and (C) GPT-4o, against neuropathological diagnoses as gold standard. (D) Grouped bar plot showing taxonomy-level accuracy of top 1, top 2, and top 3 differential diagnoses made by neurologists and DIAGNO, evaluated by GPT-4o. DIAGNO achieved significantly higher top-3 accuracy than neurologists under adjudicators DHO (McNemar's test p=0.034) and GPT-4o (McNemar's test p=0.087). *Denotes statistical significance at p < 0·05.

Supplementary Figure 2.

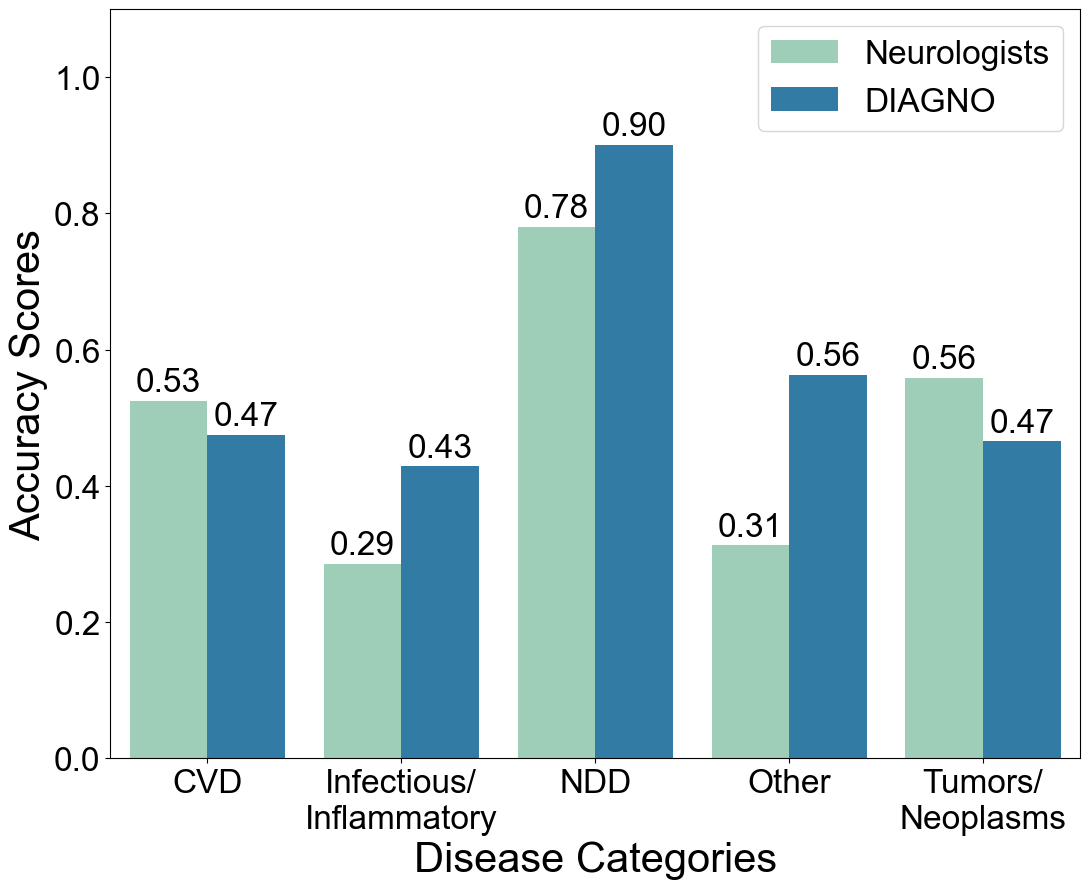

**Diagnostic Accuracy of DIAGNO and Neurologists Across Major Disease Categories**. Comparison of top-3 diagnostic accuracy between DIAGNO and neurologists across five major disease categories: cerebrovascular disease (CVD), infectious or inflammatory disorders, neurodegenerative diseases (NDD), other neurological conditions, and tumors or neoplasms. DIAGNO demonstrated higher accuracy in neurodegenerative, infectious/inflammatory, and other categories, whereas neurologists showed slightly higher accuracy in CVD and neoplastic disorders. None of the pairwise differences between DIAGNO and neurologists reached statistical significance (McNemar’s test, all p>0.05).

Supplementary Figure 3.

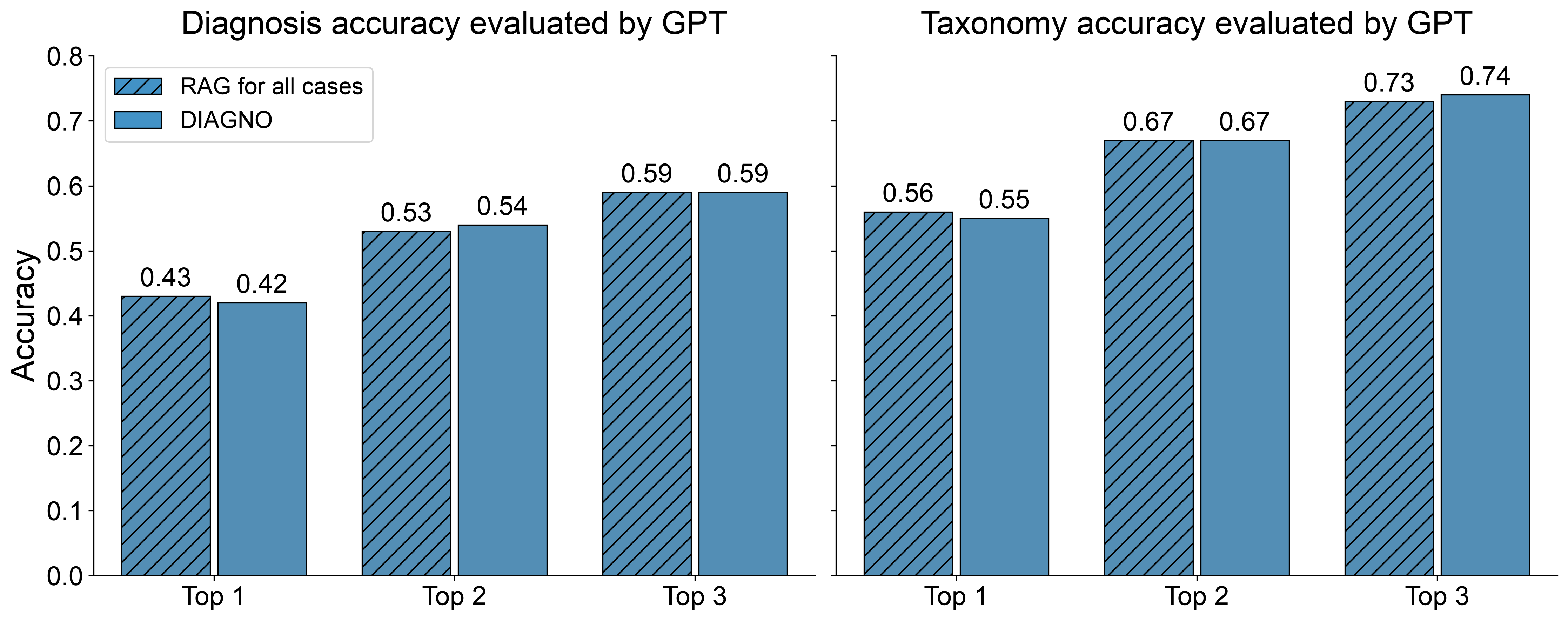

**Sensitivity analysis of the DIAGNO confidence-gating strategy.** Comparison of diagnostic performance between the original confidence-gated DIAGNO pipeline and a modified pipeline in which Stage 2 retrieval was applied to all cases, including those initially assigned high confidence. Applying retrieval to all cases did not improve diagnostic performance, supporting the effectiveness of selectively activating Stage 2 retrieval based on model confidence.

Supplementary Figure 4.

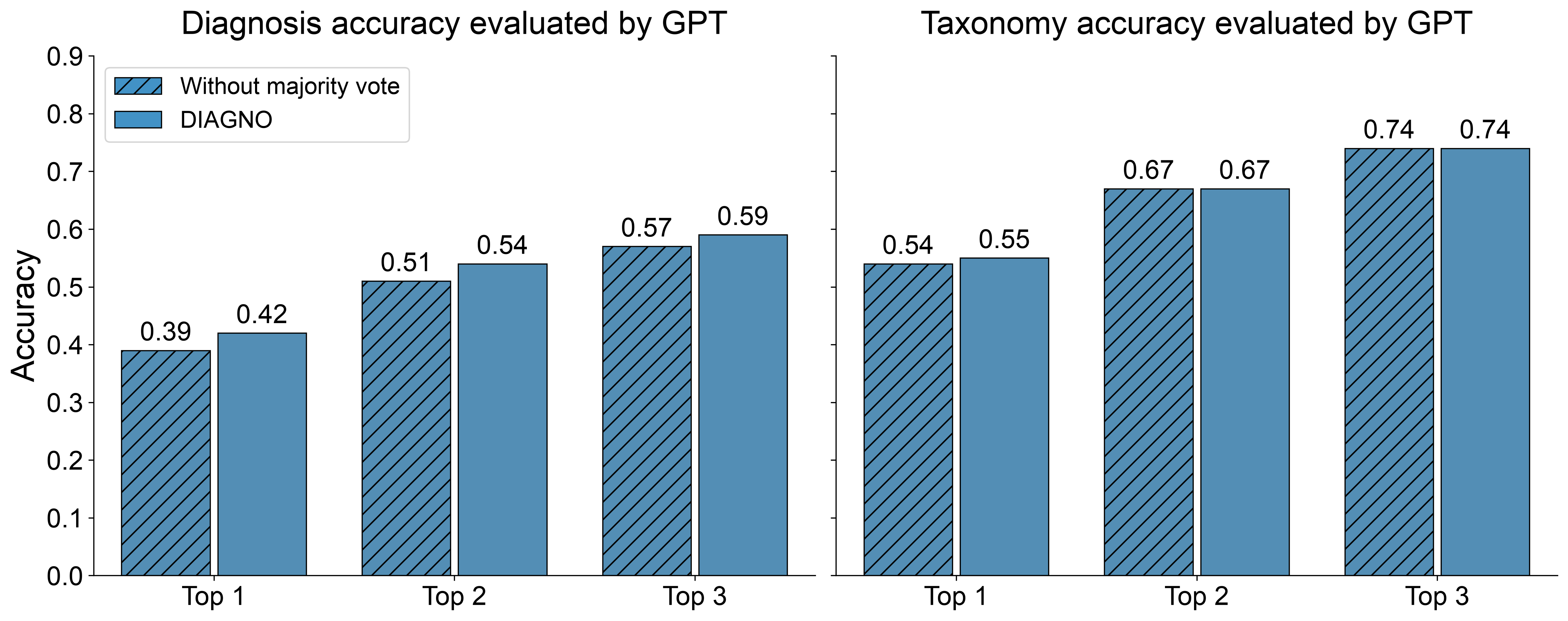

**Sensitivity analysis of the DIAGNO major voting strategy.** Comparison of diagnostic performance between the final DIAGNO pipeline with five-trial majority voting and a single RAG inference. Majority voting modestly improved exact diagnostic accuracy while producing minimal changes in taxonomy-level accuracy, supporting the contribution of the self-consistency component to the DIAGNO pipeline.

### Supplementary Notes

Supplementary Note 1. DIAGNO Prompt and Sample Response for Differential Diagnosis.

You are a neurologist specializing in neuropathological diagnosis. Rank the three most plausible categories for each clinical note while obeying the supplied closed diagnostic taxonomy and exact output format.

Select exactly three distinct diagnoses from the Allowed Neuropathological Categories below. Every returned diagnosis must be copied exactly, character for character, from this list. Do not return diagnoses, synonyms, abbreviations, variants, explanatory text, or combined labels in the numbered diagnosis lines unless they appear exactly in the list.

If the true or suspected diagnosis is not represented, select the three closest categories and set the overall Confidence Level to Low.

*** Not used for complex 203 MGH cases

Allowed Neuropathological Categories:

[the numbered 59-diagnosis list]

***

Follow this format exactly:

**Top 3 Diagnoses:**

1. [exact category label]

2. [exact category label]

3. [exact category label]

**Confidence Level:** [Low, Medium, or High]

**Summarized Reasons:** [Briefly explain why these three allowed categories were ranked highest.]

Analyze the following case summary presented at brain cutting seminar:

[CASE NOTE]

**Sample Response:**
**Top 3 Diagnoses: **

1. Mitochondrial Encephalopathy, Lactic Acidosis, and Stroke-like episodes (MELAS)

2. Frontotemporal Dementia (FTD)

3. Autoimmune Encephalitis

**Confidence Level: ** Medium

**Summarized Reasons: **

1. **Mitochondrial Encephalopathy, Lactic Acidosis, and Stroke-like episodes (MELAS): ** - **Reasons: ** The patient has a history of progressive sensorineural hearing loss, proteinuria, and elevated lactate levels, which are suggestive of mitochondrial disorder. Additionally, fluctuating imaging abnormalities, seizures, progressive cognitive and behavioral changes, and muscle cramps with elevated CK levels support this diagnosis.

2. **Frontotemporal Dementia (FTD): ** - **Reasons: ** The patient's progressive language disturbance, personality changes (short-tempered and nonsensical arguments), and regression with childish behavior and impulsivity are characteristic of FTD. The MoCA score and the tangential speech with inappropriate actions during the exam further support this diagnosis.

3. **Autoimmune Encephalitis: ** - **Reasons: ** The systemic illness with headache, malaise, and fatigue preceding the neurological symptoms, along with fluctuating cognitive impairment, agitation, seizures, and imaging findings showing new abnormalities suggest an autoimmune process. The mildly elevated protein in CSF could also be indicative of an autoimmune etiology. These diagnoses are based on the constellation of symptoms, imaging findings, and laboratory results provided in the patient's history and clinical course.

Supplementary Note 2. AI Adjudicator Prompt and Sample Responses for Neurological Diagnosis Evaluation

You are an expert in neurology and neuropathology. Your task is to determine whether a diagnosis consistent with a neuropathologically verified gold-standard.

Gold Standard Diagnosis (Neuropathology): {dx_1}

Diagnosis to Evaluate: {dx_2}

Evaluate whether the comparison diagnosis is correct with respect to the underlying neuropathology. Provide your evaluation based on the following categories:

1. **Right**: The diagnoses are broadly consistent or sufficiently specific to describe the same condition.

2. **Wrong**: The diagnoses are completely different and describe unrelated conditions.

Keep the response concise and follow this format:

**Answer: ** [Right, or Wrong]

**Confidence Level: ** [High, Medium, or Low]

**Justification: ** [Briefly explain similarities or differences in pathology and context.]

**Sample Response 1:**

**Answer: ** Right

**Confidence Level: ** High

**Justification: ** Both diagnoses are "Multiple System Atrophy," indicating they describe the same neurodegenerative condition affecting the same subtype (predominantly parkinsonian). There is no difference in specificity or context between the two provided diagnoses.

**Sample Response 2:**

**Answer: ** Wrong

**Confidence Level: ** High

**Justification: ** Spinocerebellar Atrophy Type 3 (SCA-3) and Progressive Supranuclear Palsy (PSP) are completely different conditions. SCA-3, also known as Machado-Joseph Disease, is a genetic neurodegenerative disorder affecting the cerebellum and spinal cord, leading to ataxia and other motor impairments. PSP is a distinct neurodegenerative condition characterized by the accumulation of tau protein, primarily affecting the brainstem and basal ganglia, leading to symptoms such as vertical gaze palsy and postural instability. The two conditions have different etiologies, pathophysiologies, and clinical presentations.

Supplementary Note 3. Mathematical Formulation of Top-k Rank-Based Concordance Scores

Let:

- $D^{GPT}=[d_{1}^{GPT}, d_{2}^{GPT},\ldots, d_{k}^{GPT}]$: top-k ranked differential diagnoses made by DIAGNO.
- $D^{Neurologist}=[d_{1}^{Neurologist}, d_{2}^{Neurologist},\ldots, d_{k}^{Neurologist}]$: top-k ranked differential diagnoses made by neurologists.
- $N$: total number of cases evaluated.

We have:

$${Agree}_{k}\left( i \right)=\left\{ \begin{aligned} 1, &if D_{1:k}^{GPT}(i)\cap D_{1:k}^{Neurologist}(i)\neq\emptyset\\ 0, &Otherwise \end{aligned} \right.$$

Where:

- $D_{1:k}^{GPT}(i)$ denotes the set of top-k diagnoses made by DIAGNO for case $i$.
- $D_{1:k}^{Neurologist}(i)$ denotes the set of top-k diagnoses made by neurologists for case $i$.

Therefore, the Top-k Concordance across all cases is:

$$Top k Agreement=\frac{1}{N}\sum_{i=1}^{N} {Agree}_{k}\left( i \right)$$

Supplementary Note 4. Fifteen Examples of Cases where Both DIAGNO and Neurologist were Correct.

**Example 1:**

Neuropathological definitive diagnosis: Alzheimer's Disease

DIAGNO/Neurologist diagnoses:

|  | Top 1 Dx | Top 2 Dx | Top 3 Dx |
| --- | --- | --- | --- |
| DIAGNO | Alzheimer's Disease | Vascular Dementia | Lewy Body Dementia |
| Neurologist | Alzheimer's disease | Vascular dementia | Dementia with Lewy bodies |

Case summary: A XX-year-old right-handed patient was noticed by their family to have subtle but worsening forgetfulness. They also observed the patient to be misplacing items, having decreased initiative and difficulty performing complex tasks. Over the next two years, these problems increased such that the patient had difficulty performing tasks that had previously been straightforward for them. They also described new word-finding difficulty. In addition, the patient became more withdrawn and dysphoric, and was begun on medication.

The patient's past medical history was significant for hypertension, hyperlipidemia, glaucoma, remote surgery for sinus disease and multiple recent pre-syncopal episodes. Their medications included an antidepressant, a beta-blocker, a statin, aspirin, and eyedrops. The patient had stopped any alcohol intake one year prior to presentation. They had smoked a pipe for several years in their youth. The patient lived at home with their partner. They had a graduate level degree in criminal justice and were a retired police officer. The patient's mother died at age XX and father at age XX. Two of their siblings had died in their early XXs from heart disease.

The patient was brought to MGH for evaluation at age XX by their partner and two children because of their concerns. The patient denied any difficulty with their memory and insisted they were presenting to the clinic only because of “the girls” wishes. Their vital signs and general examination were unremarkable. The patient was able to learn four words on the 2nd attempt; at 5 minutes, they recalled none spontaneously, none with a clue, and only 1 of 4 correctly with multiple choice prompts. The patient was incorrect on significant historical details (large gaps about 9/11; reporting that Nixon got sick and retired to Texas), although intact on others (that 2 space shuttles had blown up; details of the JFK assassination). Their knowledge of current events was similarly uneven, with minimal information about recent presidents or a then newly elected governor although they had details on other topics. The patient was entirely unable to perform serial 7’s -- cannot get to 93 and cannot continue from there. Their forward digit span was 7 and reverse digit span was 2 on multiple attempts. Praxis was normal for buccofacial, ideational, and ideomotor testing. The patient had great difficulty with concept formation in the similarities test. Their phonemic fluency was 14 F words in a minute (but with 2 repeated words). They had impairment of alternating sequences in the fist-palm-side test. Their visual fields were full to confrontation, fundal exam was normal (left eye was s/p cataract removal). Their eye movements were full with no nystagmus, facial movements were symmetric and facial sensation was intact. Their hearing was intact to high and low tones. The palate and tongue move in the midline. There was no dysarthria. Sensation is normal to all modalities, including graphesthesia and double simultaneous tactile and visual stimulation. Their strength was full in both upper and lower extremities and symmetric across both sides. There was a hint of cogwheeling in the right upper extremity with rapid tapping of the left hand. There was some minimal tremulousness of the outstretched hands but no tremor at rest. Their right arm swing has slightly decreased swing with their stride, but gait was otherwise normal, and tandem and Romberg are preserved. Their coordination was entirely normal. Reflexes were 2+ in the right upper extremity, 2 in the left upper extremity, 2 in both legs, and plantar responses bilaterally flexor. There is a subtle cross adductor response on the right side. Their MMSE was 27/30. Diagnostic studies were performed and management was begun.

The patient was followed in outpatient psychiatry clinic, and no detailed neurologic examinations are available in the records. A year after their first assessment, their MMSE was reported to be 21/30 and another medication was added.

Two years after their presentation, the patient had a syncopal episode from 3rd degree AV block. A pacemaker was placed. At that time, the patient was still able to prepare their own lunch, do the dishes, shovel the snow, and collect the trash. However, their memory and initiative were worse. They wouldn’t change their clothes if not reminded and they were easily overwhelmed and disoriented in crowds. Their MMSE was 17/30.

Five years after their initial presentation, the patient had minimal spontaneous verbal output but could appropriately respond to Yes-No questions. They were not able to shower by themselves and were incontinent of urine and stool. They were having repeated falls, which prompted cardiology evaluations because of concerns about presyncopy.

The patient died XX years after their initial symptoms (XX years after their evaluation at MGH) and an autopsy was performed.

**Example 2:**

Neuropathological definitive diagnosis: Amyotrophic Lateral Sclerosis (ALS)

DIAGNO/Neurologist diagnoses:

|  | Top 1 Dx | Top 2 Dx | Top 3 Dx |
| --- | --- | --- | --- |
| DIAGNO | Amyotrophic Lateral Sclerosis (ALS) | Hereditary Spastic Paraplegia (HSP) | Multifocal Motor Neuropathy (MMN) |
| Neurologist | Amyotrophic lateral sclerosis (ALS) | Spinal muscular atrophy (SMA) Type 4 | Progressive Muscular Atrophy (PMA) |

Case summary: A XX-year-old patient had been experiencing intermittent fasciculations for XX years in the face and arms, which were relatively stable in severity and frequency. The patient then began to experience intermittent wave-like rushing sensations down both arms like ""muscles were prickling or bunching up"", associated with taking a deep breath. The patient had no significant past medical history but did have a family history of neurologic disease on their father’s side (he died at age XX) including 3 of 5 siblings as well as at least 4 first cousins. Because of the family history, the patient had genetic testing done, which were unrevealing. The patient was seen at outside hospital at that time where studies were performed without abnormality. Ultimately, it was felt that despite the family history, the stability of symptoms and other studies made the diagnosis of the familial neurologic disease unlikely.

Approximately XX months later, the patient started to have trouble cross-country skiing, such that they fell multiple times (highly unusual for such an experienced skier). The patient was falling quite frequently while going downhill and their L hip/knee would give out. Soon, the patient noticed more trouble on other routine ski trails. After several falls, the patient started to have low back pain radiating into leg. The patient was assessed without significant findings and received PT without much benefit. A few months later, the patient had trouble standing on their toes (on the left) and reported that they were limping in compensation for left sided weakness. The patient also thought the left quadriceps was weaker as well. A few months later, the patient had trouble with hip flexion and noticed some fasciculations. Additional laboratory studies were performed, with positive findings. The patient was also determined to have an indeterminate Western blot result for Lyme disease and was started on tetracycline and amantadine.

The patient had worked previously as a nurse, but around this time went out on disability. The patient was working as a nurse but within a year of their first assessment was out on disability. The patient began to use a cane about XX months after they noticed their impaired skiing, shifting to a motorized scooter the next month.

On the time of assessment at MGH the patient was still independent with ADLs. The patient reported that their left leg plegic with some left hand weakness. The patient did not feel that they had any deficits with their right arm or leg. The patient reported that their fasciculations were more prominent in the face and arms. The patient had no dysarthria, dysphagia, diplopia, dyspnea, orthopnea, or AM headache. The patient described being more emotionally labile which they attributed to the stress of their symptoms/diagnosis.

The patient was alert, attentive, fully oriented with coherent and fluent speech. Memory, comprehension, and ability to follow commands were intact. Their extraocular movements and visual fields were full. There was mild weakness of cheek puff, but no other signs of bulbar weakness. Facial sensation was intact to temperature and light touch. Hearing was intact to finger rub. Shoulder shrug and neck flexion were both normal strength. Overall, the patient demonstrated normal muscle tone. Bulk was reduced in the L forearm, L FDI, L thenar eminence, L quadriceps, and L calf. There were scattered fasciculations observed in the L forearm. Manual muscle testing revealed MRC grade strength the following (R/L): Neck flexion 5, Neck extension 5, elbow flexion 5/4, elbow extension 5/4+, wrist flexion 5/5, wrist extension 5/5-, finger flexion 5/5, finger extension 5/4+, APB 5/4, FDI 5/4, ADM 5/5-, hip flexion 5/2, hip extension 5/2, hip abduction 5/2, knee extension 5/1, knee flexion 5/2, ADF 5/1, APF 5/1. There was normal sensation for vibratory perception, pinprick, light touch and proprioception in the bilateral upper and lower extremities. The patient’s reflexes were 2+ in the biceps, triceps, brachioradialis bilaterally, but somewhat more brisk on left. 2+ in the right patella and ankle while absent on the left. The patient’s toes were mute. There was a positive Hoffman's sign bilaterally. Complex motor skills testing revealed normal coordination for finger-nose-finger, rapid-alternating-movements and heel knee-to-shin. There was no tremor, rigidity or cogwheeling. The patient required assistance to stand and essentially dragged the left leg when attempting to move.

The patient was not seen again at MGH. The patient died approximately XX months later (age XX) and an autopsy was performed.

**Example 3:**

Neuropathological definitive diagnosis: Corticobasal Degeneration (CBD)

DIAGNO/Neurologist diagnoses:

|  | Top 1 Dx | Top 2 Dx | Top 3 Dx |
| --- | --- | --- | --- |
| DIAGNO | Corticobasal Degeneration (CBD) | Parkinson's Disease (PD) | Progressive Supranuclear Palsy (PSP) |
| Neurologist | Corticobasal syndrome | Progressive supranuclear palsy | Parkinson's disease |

Case summary: A XX-year-old right-handed patient presented to the clinic with left hand clumsiness. Over the past two years, the patient had found increasing difficulty in using their left hand. The patient played drums as a hobby and noticed that they could not tap out rhythms with the left hand. As this progressed, the patient had trouble using utensils or fastening buttons with the left hand and noticed a left hand tremor that was worse when moving their arm. The incoordination and tremor were exacerbated by stress and in situations where the patient could not see their hand. The patient's partner was concerned because the patient continued to drive, despite what the partner noticed to be a lack of left-sided spatial awareness: the patient often bumped into objects with their left side while walking. The patient also acted out their dreams at times.

The patient's past medical history was significant for an early-stage laryngeal cancer that was treated with laser ablation. The patient also had a XX-year history of depression. The patient's only medication was an antidepressant. The patient characterized their current symptoms as lack of motivation and social withdrawal. The patient's mother died from intracranial hemorrhage, but family history was otherwise noncontributory.

When examined, the patient was friendly and mentally alert without clear impairment in language. The patient's voice was hoarse because of prior surgery. Visual fields were full, with normal saccades and smooth pursuit. The patient's cranial nerves were intact. The patient had rigidity of the neck and left arm. Strength appeared full and symmetric in all four extremities on confrontational testing. Rapid repetitive movements of the left fingers, wrist, and arm were markedly slowed, while the right side and both lower extremities were normal. There was a 3 Hz action tremor of the left arm. Stereognosis was impaired, but other sensory modalities, including graphesthesia, pinprick, vibration, and proprioception were normal, and there was no extinction to double simultaneous stimulation. Deep tendon reflexes were unremarkable, and there was no dysmetria. Romberg's sign was negative; the patient's gait was notable only for a mildly reduced left arm swing. The patient scored 29/30 on the Mini-Mental State Examination (-1 for recall), and 26/30 on the Montreal Cognitive Assessment. The patient's digit span was 6 forward, and 3 backward. The patient named 16 animals; 13 vegetables; 10 fruits; 7 ""F"" words; 6 ""L"" words; and 11 ""S"" words. Following the visit, the patient's medications were adjusted.

The patient returned to the clinic XX months later. The patient experienced side effects of medication, causing them to discontinue treatment and start self-medicating with cannabis gummies. The pace of progress in physical and occupational therapy discouraged the patient from continuing. These frustrations in concert with the patient's depression manifested as suicidality, anger, and screaming at their partner. The patient's coordination worsened and the patient ran into objects more often; the patient's medication regimen was adjusted, and imaging was performed. Soon after, the patient felt increasing hoarseness, and recurrence of their laryngeal cancer was confirmed by laryngoscopy. A total laryngectomy was performed but was complicated by a postoperative hematoma. Afterward, the patient felt easily triggered into a depressed mood and had frequent rage episodes, describing their experiences with colorful profanities in the clinic.

Over the next two years (to age XX) these symptoms progressed to where the patient felt pain in their left shoulder and had little function remaining in their left hand. The patient was unable to grab objects or use utensils. The patient's balance and coordination suffered, and the patient fell occasionally. On examination, the patient had myoclonus and posturing of the left arm with dystonic flexion of the elbow, pronation of the forearm, and intermittent flexion of the second, third and fourth digits. Left leg muscle tone was increased and the patient had difficulty performing heel tapping on the left. Initiation of saccades was slowed. Symptomatic management of the patient's tremors and spasticity was begun, with some benefit.

The patient continued to have worsening disability, spasticity, tremors and pain in their left arm, in one instance falling onto the floor and fracturing their left hip. Standing from a seated position became difficult, requiring the patient to push up with their right arm. The patient's left arm and hand felt contracted, and the patient reported electrical shock-like sensations in their left hand, biceps, and hip, without relief from medications. The patient felt helpless and dependent, reporting problems with balance that precluded them from going up or down stairs in their home, prompting a move to an assisted living facility. A Palliative Care specialist helped to optimize the patient's pain control with oral pain medication. The patient died one year later (at age XX, four years after their initial presentation) and an autopsy was performed.

**Example 4:**

Neuropathological definitive diagnosis: Cerebrovascular disease, acute left pontine hemorrhage

DIAGNO/Neurologist diagnoses:

|  | Top 1 Dx | Top 2 Dx | Top 3 Dx |
| --- | --- | --- | --- |
| DIAGNO | Intracerebral Hemorrhage (ICH) | Subarachnoid Hemorrhage (SAH) | Acute Hepatic Encephalopathy |
| Neurologist | Intracerebral hemorrhage | Hepatic encephalopathy | Osmotic demyelination (myelinolysis) |

Case summary: A XX-year-old individual developed a headache and became less responsive while an MGH inpatient. The patient had a history of cirrhosis secondary to alcohol as well as possible chronic HBV, pulmonary HTN, T2DM, and HTN. The patient had been admitted to the hospital 22 days earlier, following an episode of hematemesis and hematochezia on the day prior to admission, also with RUQ pain. At that time, the patient reported drinking 2-3 ""40s"" a night, but occasionally more on some other days of the week. On arrival to ED, the patient was initially afebrile with BP of 140/100. The patient's labs included WBC 13 (75% N, 2% bands), Hgb 11.9, Cr 3.85 (previous < 1 in 2014), ALT 125/ AST 394 (hemolyzed), Tbili 17.8/ DBili 13, INR 1.5, lactate 3.6. The MELD-Na score was 36. Serum toxicology was negative for salicylates/acetaminophen, with an ethanol level 57. Medical management was begun. The patient was also noted to have acute kidney injury with an elevation of serum creatinine in the 2-3 range (baseline of 0.8-1.1). The patient was not considered to be a liver transplant candidate given multiple prior medical complications related to alcohol use in the setting of multiple discussions about the ongoing risk of continued drinking. Additional labs sent showed: ANA pos 1:160, IgG elevated (1914), free light chains (elevated), LKM-1 neg, anti-mitochondrial positive 1:40/anti-SMA positive 1:160. HBV serologies were discordant with negative HBV sAg and HBV DNA < 20 but HBV eAg positive. The patient also had a coagulopathy for which a trial of IV vitamin K x 3 days (on day 20-22 of the hospitalization) was given.

The patient was improving on the medicine floor when on day 22 of the hospitalization the patient complained of a headache. With the initial complaints, the patient was assessed by the nursing staff, received a PRN dose of acetaminophen and heat packs. The patient's vital signs were stable at that time. About 3 minutes after this, the patient began to shout with pain and staff returned to the room. The patient was initially still oriented and able to communicate, although slightly confused and agitated. The patient's SBP was 160s, the patient was following commands in all extremities, able to move their eyes but not briskly responsive. The patient was also noted to have bilateral sustained lower extremity clonus. The patient quickly became less responsive, opening eyes to sternal rub only, mumbling noises, and with a left gaze preference and left sided weakness. The patient had a lateral tongue bite. There was projectile vomiting of yellow fluid. The patient was intubated, an imaging study was done and the patient was transferred to the neuro ICU.

On arrival to the neuro ICU, the patient's exam was as follows: Dilated and unreactive left pupil, 1mm but reactive pupil on the right, absent corneal responses, absent VOR, absent gag, absent cough, no movement in any extremity. The patient did have some spontaneous breaths on the ventilator. An attempted intervention was undertaken after a discussion with the patient's health care proxy. Shortly after, the patient was noted to have bilateral fixed pupils (L at 3mm, R at 6mm) in association with the persistent other absent brainstem reflexes. The patient's arms now showed no movement to noxious stimuli, while both legs showed triple flexion.

Two days later following discussion with the patient's family, a decision was made to withdraw care and the patient was transitioned to comfort measures only. The patient was terminally extubated, died shortly afterwards and an autopsy was performed.

**Example 5:**

Neuropathological definitive diagnosis: Fronto-temporal lobar degeneration, severe (FTD-TDP43)

DIAGNO/Neurologist diagnoses:

|  | Top 1 Dx | Top 2 Dx | Top 3 Dx |
| --- | --- | --- | --- |
| DIAGNO | Frontotemporal Dementia (Behavioral Variant FTD) | Lewy Body Dementia | Alzheimer’s Disease |
| Neurologist | Behavioral variant frontotemporal dementia | Behavioral variant Alzheimer's disease | Autoimmune/paraneoplastic encephalopathy |

Case summary: A XX-year-old right-handed patient developed aggression and behavioral changes over a few years. The family felt that the patient was “quick-tempered” and would often yell at strangers. The patient also had significant road rage and had been arrested after threatening another driver with a knife. The patient also seemed paranoid, frequently staring suspiciously at cars as they passed by their home. The patient also felt that people in the streets were purposely bumping into them. At the same time, the patient became increasingly concerned about cleanliness: the patient would walk around the house with a flashlight looking for dust, vacuum many times per day, and would yell at strangers for sneezing. The patient frequently repeated themselves and had occasional word finding difficulties. The patient complained of bilateral shoulder tingling, but no other numbness or weakness.

The patient's medical history was notable for right shoulder replacement XX years prior. The patient felt that they were permanently changed by the anesthesia during the procedure: their sense of taste changed, their attention worsened, and they lost interest in their typical daily activities. History was also remarkable for depression (diagnosed in their 40s), personality disorder NOS, lattice degeneration of the eye, hypertension, and pan-hypopituitary syndrome requiring levothyroxine and testosterone injections. The patient also underwent resection of a right thigh melanoma five years earlier (negative sentinel node; no further treatment recommended). The patient did not drink nor smoke. The patient had worked as a graphic designer for over 20 years but had abruptly quit their job four years before presentation because they no longer liked their boss and coworkers. There was no family history of neurologic disease.

The initial examination was remarkable for an abnormal, flattened affect and diminished insight. MMSE was 28/30 and the patient recalled 2/3 words. The exam was otherwise normal. The patient underwent neuropsych testing, notable for FSIQ=90 (expected = 115), MMSE=29/30, trails A = 89 seconds (1st %tile), trails B = 160 seconds (3rd %tile) and decision speed in the 1st %tile. The patient had impaired proverb interpretation, performed at <1st %tile in learning a list of 12 words, and confrontation naming was 23/30 (2nd %tile). Facial recognition and clock recognition were intact. An MRI and an LP revealed abnormalities and a diagnosis was confirmed. The patient's treatment regimen, started prior to this visit, was continued.

Three months later, the patient stopped treatment because they were concerned it was causing hair loss. Over the following few months, the patient's behavior worsened. The patient began having insomnia and tearfulness, often crying about past memories. The patient stopped reading the newspaper and no longer enjoyed viewing artwork—on a visit to the MFA, the patient described that the paintings were “all different than before.” The patient also developed an ‘earworm’ of a specific TV commercial jingle and acquired an intense appetite for sweets, replacing most meals with ice cream. Testing performed a year after the first assessment showed deterioration at confrontation naming and facial recognition but was otherwise stable.

The patient returned to the clinic one year later (now two years since initial assessment). The patient was apathetic and no longer cared if the house was clean. The patient's diet had changed to include only donuts and soda. The patient was more forgetful and frequently had word finding difficulties. On exam, the patient's speech was pressured and they were perseverative. When asked to draw a clock, the patient replied, “What’s a clock?”. The patient was not oriented to date, could not recall recent events, and was unable to recognize any famous faces. The exam was otherwise unchanged. The patient continued to refuse all treatment.

One year later (now three years since initial assessment), the patient's partner called the clinic with concern for the patient's safety. The partner caught the patient playing with razors and the patient had left the gas fireplace on. The patient no longer talked, was incontinent, slept most of the day, and no longer recognized their partner.

The patient was admitted to an OSH for treatment initiation before transfer to a nursing facility. During this admission, the patient died suddenly at the age of XX (four years after initial assessment and approximately 8 years after symptom onset) and an autopsy was performed.

**Example 6:**

Neuropathological definitive diagnosis: Huntington's disease, Vonsattel Grade 1 of 4

DIAGNO/Neurologist diagnoses:

|  | Top 1 Dx | Top 2 Dx | Top 3 Dx |
| --- | --- | --- | --- |
| DIAGNO | Huntington's Disease | Progressive Supranuclear Palsy (PSP) | Major Depressive Disorder with Cognitive Impairment |
| Neurologist | Huntington's disease | Frontotemporal dementia with motor neuron disease (FTD/ALS) | Alexander's disease |

Case summary: A XX-year-old right-handed patient with a past medical history of depression and anxiety presented to MGH neurology with a chief complaint of difficulties at work. The patient reported that for the previous 2 years they had had a progressive decline in work performance. They had worked in a medical billing office for 8 years and were beginning to get poor work reviews. They described difficulties in learning new tasks and adjusting to changes at work. Their organizational skills had worsened: they were inattentive, had difficulty focusing, and were forgetful. They had always been an anxious person but their anxiety had worsened over the same time period and they had begun having panic attacks. Other than these cognitive complaints, they had no other symptoms such as weakness, sensory changes, or gait imbalance.

Their past medical history included depression and anxiety with self-injurious behavior (cutting) since they were a teenager. They were divorced and had two foster children that they were considering adopting. They did not consume alcohol, tobacco or illicit drugs. The patient had several family members with similar symptoms. Their mother became wheelchair bound (died at XX). They had one sibling who was healthy.

General physical exam at this time was notable for obesity (304 lbs). On neurological exam, they were awake, alert and oriented with normal affect. Pupils were equal and reactive. Visual fields were full. Extraocular movements had full range but slightly discontinuous smooth pursuit. Saccades were slightly delayed but had normal velocity and range. Their face was symmetric with normal sensation. Speech was unremarkable. Muscle tone, bulk, and strength were normal. There was no bradykinesia. Luria was learned easily. Sensation was intact. Their gait was slightly wide based and they had 3 deviations during tandem walking. They could recover during retropulsion. Based on their symptoms, medication changes were made.

On follow-up three months later, the patient reported increasing difficulties at work. They were unable to keep up with their work load due to difficulties with memory, organization and multitasking. On average, their daily work load took them 2 hours longer to complete than previously. Their anxiety was also getting worse and they had begun to avoid social situations. Their exam was unchanged. Additional medication changes were made.

Three months later, their anxiety and mood were improved but their cognitive difficulties persisted, leading them to decide to give their foster children up for adoption. They had lost 70 lbs over the previous 6 months. On neurological exam, their mental status was intact (MMSE = 30/30). Visual fields were full. Smooth pursuit was discontinuous and saccades were delayed and slightly slow. Facial strength and sensation were intact. They could protrude their tongue for 10 sec. Muscle bulk, tone and strength were intact. Pronation/supination movements and finger tapping were slightly impaired. Occasional adventitious movements were seen in the hands, feet and shoulders. Their gait continued to be wide with difficulty tandem walking. Neuropsychological testing revealed impairments primarily in sustained attention, executive function, and learning/memory.

Three months later the patient became severely depressed in the setting of their ex-partner committing suicide and was admitted for inpatient psychiatric care. They were treated with medications and ECT. Immediately after discharge, they were admitted to another hospital for continued depression and their medications were changed. Several weeks after their discharge, they were admitted to MGH in the setting of continued depression, worsening anxiety, and complaints of weakness, vertigo (with a sense of being pulled to the right), and gait difficulties requiring a walker. On admission, their affect was blunted and they made minimal eye contact. They were attentive and language was intact. Memory was 1/3 at 5 minutes. Cranial nerve exam was unremarkable. Muscle tone, bulk and strength were normal. Their movements were felt to be bradykinetic. Finger-nose-finger and alternating movements were slow but otherwise unremarkable. Their gait was slow and guarded and they required a walker. Several medications were started.

On follow-up one month later, they continued to have significant symptoms of depression and anxiety with insomnia, nightmares, fatigue, and loss of interest in activities. They continued to have impaired saccades, finger tapping and pronation/supination. They had a new high frequency, low amplitude tremor. Their gait was wide-based and labored with exaggerated slowness and lurching. On the Montreal Cognitive Assessment they scored 23/30. New medications were started but the patient continued to have significant depression and anxiety. Six months later, they committed suicide and an autopsy was performed.

**Example 7:**

Neuropathological definitive diagnosis: Lewy body disease (LBD/DLB)

DIAGNO/Neurologist diagnoses:

|  | Top 1 Dx | Top 2 Dx | Top 3 Dx |
| --- | --- | --- | --- |
| DIAGNO | Dementia with Lewy Bodies (DLB)/Lewy Body Dementia (LBD) | Alzheimer's Disease (AD) | Parkinson's Disease Dementia (PDD) |
| Neurologist | Dementia with Lewy Bodies | Parkinson’s disease dementia | Corticobasal degeneration |

Case summary: A XX-year-old right-handed patient was concerned because of ‘foggy’ thinking. The patient had undergone a partial thyroidectomy for a benign lesion, following which they noted onset of difficulty with memory, trouble finding words, and tremulousness. This was initially thought to be related to anxiety in the post-surgical setting.

The patient's medical history was notable for hypothyroidism, thyroid nodule s/p resection (benign adenoma), hyperlipidemia, and both squamous cell and basal cell carcinomas excised during the prior 6 years. The patient was retired from work as an architect. Family history was notable for a mother diagnosed with Alzheimer’s disease in her 60s, and a father who died of heart disease in his 80s; the patient had two siblings who were alive and well with another having died in an accident from head injury following a bicycling accident.

At a follow-up visit about 7 months after the surgery, both the patient and their family continued to report mild difficulty with word retrieval, and thought processing as well as temperature sensitivity and difficulty falling back asleep after wakening during the night. A test was performed by the primary care physician that was normal. The patient was referred for imaging that showed abnormalities.

Over the next few months, the family noted continued memory issues that would fluctuate considerably. The patient's partner also noted difficulty with spatial processing. For example, the partner noted that while driving the patient would suddenly stop inappropriately, leading to an accident on one occasion (this led to the patient stopping driving). The patient otherwise remained independent with all ADLs and iADLs but had been late on a couple of bill payments. The primary care repeated a test that was abnormal, and a medication was started.

The patient established care with neurology 2 years after symptom onset. During this visit, the patient's partner reported that the patient's forgetfulness had begun before the thyroid surgery and initially improved in the post-op period before worsening again until the medication had been begun. On exam at this visit, the patient was noted to respond slowly to questions with a soft voice. The patient did not know their age, the president, or vice president. The patient had word recall of 1/5 at 5 minutes, though on a second chance could recall 5/5. The patient's speech was fluent, without error, naming, repetition, and comprehension were all intact. The remainder of the exam was normal apart from a positional tremor.

The patient returned to the clinic around 9 months later when they reported increasing difficulty getting dressed, struggling with getting on sleeves, pulling up pants, and buttoning shirts. The patient's cognitive functioning was felt to be stable. The exam at that time was notable for normal speech, recall was now 0/5 at 5 minutes, and the patient had several mistakes when reciting the months of the year backwards. The patient was noted to have restricted upgaze, a resting tremor on the left as well as cogwheel rigidity, more on the left greater than right. The patient's gait was notable for bilaterally reduced arm swing and mildly stooped posture. Medical management was begun with definite improvement.

Roughly 3 years after symptom onset, the patient was no longer able to pay bills or help with most household chores. The patient was still independent with ADLs. Around this time, the family reported that, for about a year, the patient had progressive visual distortions stating that “things do not look right”. Additionally, the patient would intermittently see others in the house when they were not there. The patient's neurologic exam was notable for worsening cognition, persistent cogwheel rigidity, and stooped posture. The patient now made en-bloc turns when walking. Additional medical management was begun.

Over the next few years, the patient had progressive cognitive and motor decline to the point that they were nearly wheelchair-bound and minimally verbal. The patient also had worsening of behavior, with the development of severe paranoia and fixed delusions. The patient was ultimately transitioned to hospice care where they died 7 years after symptom onset and an autopsy was performed.

**Example 8:**

Neuropathological definitive diagnosis: Mitochondrial encephalomyopathy with lactic acidosis and stroke-like episodes (MELAS)

DIAGNO/Neurologist diagnoses:

|  | Top 1 Dx | Top 2 Dx | Top 3 Dx |
| --- | --- | --- | --- |
| DIAGNO | Mitochondrial Encephalomyopathy, Lactic Acidosis, and Stroke-like episodes (MELAS) | Chronic Progressive External Ophthalmoplegia (CPEO) | Mitochondrial Myopathy |
| Neurologist | MELAS (Mitochondrial encephalomyopathy with lactic acidosis and stroke-like episodes) | Mitochondrial myopathy/Kearns-Sayre syndrome | Lysosomal storage disease |

Case summary: A patient in their teens developed migrainous type headaches. They described them as a pounding pain located over the bitemporal and occipital regions associated with nausea, emesis, and photophobia. The patient went on to have severe chronic fatigue as well as episodes of diarrhea and constipation. They were at one point hospitalized for colonic pseudo-obstruction.

The patient was adopted and there was no known information regarding their biological parents. They had a child who developed epilepsy during childhood and later passed away in their early XXs. They had another child who they lived with who had a history of migraines and some minor learning difficulties. Their medical history included the emergence over time of cataracts, COPD, adult-onset type 2 diabetes, and hyperlipidemia.

Neurologic exams at age XX and XX showed mild facial and proximal muscle weakness, some difficulty with finger-to-nose performance but no other clear abnormalities. When the patient was XX, their child (age XX) was complaining of migraine headaches associated with vomiting and well as problematic abdominal cramping and loose stools. Their neurologic exam showed ptosis, facial weakness, 4/5 overall strength in the arms and legs with slightly brisk reflexes in the legs, equivocal plantar responses and awkwardness on finger-to-nose testing.

The patient subsequently developed difficulty climbing up or down stairs and began using a walker for assistance. They endorsed difficulty with balance on uneven surfaces and had fallen several times in the past. There was a report of a stroke sometime in their XXs, but the neurologic symptoms that they experienced were unspecified. They also noted gradual onset of paresthesias and numbness in their hands and feet. They had some incontinence of urine because they were unable to get to the bathroom in time due to weakness. They had chronic hearing loss for which they had been recommended hearing aids.

The patient was initially seen at MGH in their early XXs. At that time, they lived with their child due to some mild cognitive decline as well as limited mobility. Neurologic exam at age XX was notable for bilateral hearing loss, diffuse weakness, hyporeflexia, slowed rapid alternating and point-to-point movements as well as a wide based, hesitant and shuffling gait, requiring support to stand up and turn.

At age XX the patient developed an acute deterioration in their hearing to the point at which their child noted that the patient was unable to understand them even when shouting. The patient was able to respond appropriately to written statements and their speech was initially fluent. Over the next day, they developed “word salad” and garbled speech. They subsequently stopped being able to communicate via written notes. The patient initially presented to an outside hospital with these symptoms where they had minimal verbal output and were unable to follow commands. While at the outside hospital they had two witnessed generalized tonic-clonic seizures. They were subsequently transferred to MGH.

On arrival to MGH, the patient required deep sternal rub to awaken. They had no verbal output and did not follow commands. They would briefly regard after being stimulated, but they did not track. Their pupils were equal and reactive. There was no gaze deviation. Their face was symmetric and they made spontaneous movements of all extremities. Their toes were upgoing bilaterally. Several tests were done and the patient was started on multiple treatments, with no improvement in their neurologic exam.

One week into their hospitalization the patient became bradycardic with profound hypotension. They were emergently intubated and transferred to the ICU. Their neurologic exam remained poor, with eyes closed and no verbal output. There were no spontaneous movements in any extremity, but they did withdraw their arms and legs bilaterally, brisker on the right than the left. They had a prolonged and complicated ICU course. After the patient did not experience any recovery, the family ultimately opted to withdraw care. The patient died shortly thereafter and an autopsy was performed.

**Example 9:**

Neuropathological definitive diagnosis: Multiple System Atrophy (MSA-C)

DIAGNO/Neurologist diagnoses:

|  | Top 1 Dx | Top 2 Dx | Top 3 Dx |
| --- | --- | --- | --- |
| DIAGNO | Multiple System Atrophy, Cerebellar variant (MSA-C) | Progressive Supranuclear Palsy (PSP) | Lewy Body Dementia (LBD) |
| Neurologist | Multiple systems atrophy, cerebellar subtype (MSA-C) | Corticobasal degeneration (CBD) | Multiple systems atrophy, Parkinsonian subtype (MSA-P) |

Case summary: A right handed individual developed changes in their sleep in their early 40s. Their partner first noticed prominent snoring and then, after about 3 years, the patient developed thrashing and jerking movements along with vocalizations during sleep. During this time, the patient also developed balance problems and “lack of clarity” in their speech. The patient had some difficulty with tasks (using a hammer and nail) and their writing was less neat than previously and required more concentration. The patient reported depression and problems with mood, with abnormally frequent episodes of crying. The patient reported dry mouth when anxious and occasional light headedness with standing up from lying down or with quick head turns.

The patient had a stutter as a child, but otherwise their medical history was unremarkable. Home medications included an antidepressant. Family history is notable for their mother dying of renal cancer at age 67; their father, siblings and children had no neurologic disorders. The patient worked as a sheet metal mechanic, with exposure to hexane and toluene. The patient drank ~10 beers per week until about two years into their symptoms; stopping alcohol intake did not produce improvement in symptoms.

The patient was first seen at MGH at age XX. Additional review of their history provided no evidence of bladder or erectile dysfunction. Physical exam revealed a well-appearing individual with blood pressure 134/84 left arm lying and 128/82 standing. The patient was oriented to the date, time and place. Encoding of four words required 3 attempts, but all four were recalled at five minutes. Forward digit span was seven, and reverse digit span was three. The semantic fluency was 13 and phonemic fluency 7. There are instances of repeating words both in the phonemic and semantic tests. Cranial nerve examination was notable for prominent hippus with pupillary light reflex, slight saccadic intrusion with eye movements and subtle nystagmus with lateral gaze but preserved saccades. There was a mild dysarthria. Tone, bulk and power are normal throughout. The finger-to-nose testing showed only minimal unsteadiness at the endpoint. Heel-to-shin testing showed the movement decomposed in several phases. Gait was mildly wide-based and unsteady, and the patient did not adequately stand with feet together in the tandem position. Their Romberg test was negative. The reflexes were 3+ throughout with crossed supraclavicular reflexes. Jaw jerk and palmar grasps were negative. Babinski’s sign was absent bilaterally. Sensation was intact to pin, touch, vibration and position; testing double simultaneous tactile and visual stimulation was normal. A range of testing and imaging was performed and management was begun.

The patient was seen in follow up 6 months later and reported that their balance, articulation and handwriting had worsened. On exam, their blood pressure was 126/80 lying, 110/80 1 minute after standing and 114/80 after 5 minutes. The patient had subtle saccadic intrusions into pursuit eye movements and hypometric horizontal saccades. The patient had some difficulty articulating rapid alternating syllables.

Eighteen months after their first assessment the patient was walking with a cane and holding on to objects as they moved. The patient also reported erectile dysfunction and monthly episodes of urinary incontinence. On exam, their affect was flattened. Their eye movements showed prominent saccadic intrusions into pursuit. The patient had square wave jerks at rest, hypometric saccades, nystagmus, which is gaze-evoked direction-beating in the horizontal plane and with upgaze. Eye movements were full in all directions and convergence was intact. Heel-to-shin testing showed side-to-side dysmetria on the right with in-plane jerking on the left.

A year later, the patient was falling 2-3 times per week, requiring a walker. There was coughing with solids and liquid PO intake, occasional incontinence of stool, and complete urinary incontinence. Their exam was relatively stable apart from increased difficulty understanding their speech.

On their final exam at MGH, five and a half years after first presentation, their sleep disorder was still present although there was less vocalization during sleep. The patient was fully incontinent and reported some light headedness when they first sat up in the mornings. Their exam was relatively unchanged from previously, and their mental status was well preserved – aware of major news events, able to discuss their mood, etc.

Eight months later, the patient died at an outside hospital at the age of XX (6 years after presentation to MGH and 8-9 years after onset of their symptoms) and an autopsy was performed.

**Example 10:**

Neuropathological definitive diagnosis: Neurofibromatosis Type 1

DIAGNO/Neurologist diagnoses:

|  | Top 1 Dx | Top 2 Dx | Top 3 Dx |
| --- | --- | --- | --- |
| DIAGNO | Neurofibromatosis Type 1 (NF1) | Posterior Reversible Encephalopathy Syndrome (PRES) | Brainstem Glioma |
| Neurologist | Neurofibromatosis type I | / | / |

Case summary: A XX-year-old patient developed headaches over two months. These occurred several times per day, lasted several minutes and were predominantly frontal. There was no history of light-headedness, seizures, nausea, vomiting or loss of consciousness. After two months of these headaches, the patient was found to have papilledema and was admitted to an outside hospital.

The patient's past medical history included: mild valvar pulmonic stenosis (diagnosed in early infancy); patches of skin “discoloration” (noted by age XX); and the emergence of pubic hair at age XX. There was no family history of neurologic disorders.

At the outside hospital, imaging and additional procedures were performed which established a diagnosis and provided some improvement in the patient's headaches. The patient underwent further treatment which had to be shortened due to complications. Going forward, the patient was seen by their physicians at regular intervals and, by report, was overall well. The patient had several skin lesions removed surgically. The patient completed their education, graduating from college. At age XX, the patient was referred to MGH because of an imaging abnormality observed on a routine scan.

On the patient's exam on first MGH visit, they had normal vital signs. Heart and lung exam was unremarkable to examiner. There were a dozen flat pigmented cutaneous lesions >1.5 cm in size as well as freckling in the axilla and groin. The patient also had 9 discrete cutaneous masses, either pedunculated or ‘lump-like’. The patient's mental status and language functions were intact. Pupils were reactive, visual fields were full to confrontation, and extraocular movements were intact. There was no face asymmetry. Palate and tongue were midline. No sensory or motor deficit was detected and there was no dysmetria. Reflexes were 3+ in legs, 2+ in arms throughout, with downgoing toes. The patient's gait was narrow based and they could tandem walk. No additional management was begun.

Six months later, the patient had nausea, vomiting, and headache which were progressive over about 2 weeks. These were associated with a decrease in appetite and some difficulty swallowing. The patient also noted increased headache with Valsalva. The patient was admitted and an intervention undertaken. During the hospitalization, the patient was documented to have a functional oro-pharyngeal swallow. When seen a few weeks after the hospitalization, the patient reported improvements in their symptoms. Additional management was undertaken, beginning about two months after discharge.

Two weeks into the new regimen, the patient presented to the ED after two episodes of speech arrest/garbled speech lasting about 30 seconds with a possible right mouth droop. The patient's exam was unremarkable apart from memory impairment (only 1 of 3 objects at 5 minutes) and horizontal nystagmus on left lateral gaze. The patient was diffusely hyperreflexic (3+). The new regimen was maintained although the patient continued to have episodic neurologic deficits including brief periods of diplopia lasting a few minutes in which there was relief with closing of one eye but not through any alterations of head position or gaze direction.

Four months into this treatment, the patient presented because they felt very weak upon standing. The patient had had falls, particularly when sneezing or coughing. The patient also reported intermittent bifrontal and occasional occipital headache, intermittent blurry vision and horizontal diplopia (improved somewhat after the patient received new glasses with prisms but not entirely). The patient now had some difficulty with swallowing both liquids and solids. The patient described intermittent tingling sensations of forearms, upper arms, and legs bilaterally as well as palpitations.

On admission, the patient's blood pressure was 170/95, heart rate 126, respiratory rate 18 (98% sat. on room air). The patient's only cognitive deficit was in memory testing (0 of 3 at 5 minutes, 1 of 3 with prompting). Cranial nerve exam was unremarkable. The patient's strength was 4/5 in upper extremities and 4+/5 in bilateral lower extremities but symmetric. Sensation was diffusely intact to light touch without evidence of extinction. The deep tendon reflexes were diffusely increased, with cross-adduction and sustained clonus of lower extremities. The patient's toes continued to go down. The patient had bilateral dysmetria in finger-to-nose test without truncal ataxia. The patient required a 2-person assist to stand, and had a positive Romberg’s sign. Gait was not assessed.

One day into the patient's admission, the patient was found unresponsive in bed. There was no response to sternal rub and the patient did not localize pain in any extremity. The patient was tachycardic to 140s, hypertensive to 220s, and hypoxemic on 6L with air movement audible bilaterally. The patient was transferred to the NICU, intubated and their hemodynamics and oxygenation were managed. An exam performed after sedation was weaned for 20 minutes showed the patient to be opening their eyes and following simple commands, moving all four extremities. There was a slight rightward gaze preference. The patient remained in the ICU for 6 days, with a series of ongoing discussions with the family. It was gradually decided that the patient would undergo extubation with pain/distress management guided by the palliative care service. The patient died shortly after extubation and an autopsy was performed.

**Example 11:**

Neuropathological definitive diagnosis: Emboli and vascular wall injury secondary to bacterial endocarditis

DIAGNO/Neurologist diagnoses:

|  | Top 1 Dx | Top 2 Dx | Top 3 Dx |
| --- | --- | --- | --- |
| DIAGNO | Embolic Stroke | Cerebral Venous Thrombosis (CVT) | Infective Endocarditis with Septic Emboli |
| Neurologist | Multiple ischemic infarcts (last one in L MCA) due to infective (likely bacterial) endocarditis | Mycotic aneurysms complicated by intracranial hemorrhage due to infective (likely bacterial) endocarditis | Bacterial meningitis complicated by multiple ischemic infarcts |

Case summary: A XX-year-old patient presented with back pain. The patient was diagnosed with spinal stenosis for which they received steroid injections.

Their past medical history was significant only for a known PFO and VSD noted at childhood and followed by echo until the age of XX without incident.

Subsequently, the patient presented to a community health center where they were treated for pneumonia and anemia, including drainage of the right lower lobe bronchus with biopsy (pulmonary nodules had been noted on imaging). The patient had been recently diagnosed with uveitis. The patient recovered, but continued to receive ocular steroid injections for uveitis.

Two months following their treatment for pneumonia, the patient was admitted to an outside hospital with diplopia. Imaging studies were abnormal and additional work up revealed other abnormalities. The patient recovered well and was discharged home on aspirin.

Two months after discharge, the patient presented to their PCP with complaints of weakness, fatigue, and intermittent chills over the past month. Studies were performed and the patient was sent home. Four days later, the patient then presented to a community medical center with night sweats and fever. At this time, results from previous studies were now available and the patient was admitted.

One day later, the patient reported new onset of headache with right hemianopsia. Studies showed new findings. The patient remained stable over the next week and was transferred to MGH for further management.

On arrival at MGH, the patient's exam was significant for a dense right inferior quadrantanopsia and superior quadrant with vision intact to finger movement in each eye individually. The patient remained neurologically stable for the next couple weeks while receiving treatment. Despite stable symptoms, new imaging was concerning, prompting an intervention to be performed. Over the next week, the patient began to feel more nauseated and fatigued due to recurrent episodes of atrial fibrillation.

The patient then had an episode of transient word finding difficulty which developed into clear aphasia prompting additional studies. While these additional studies were being performed, the patient became acutely unresponsive.

Examination at this point found the patient to be able to open their eyes but not follow any commands. There was a left gaze deviation, right sided plegia, and trace movement only to noxious stimuli on the left. Another intervention was undertaken. The patient's exam did not improve over the next week and the decision was made to focus on provision of only comfort measures. The patient died XX months after their first symptoms (1 month after transfer to MGH) and an autopsy was performed.

**Example 12:**

Neuropathological definitive diagnosis: Progressive supranuclear palsy (tauopathy)

DIAGNO/Neurologist diagnoses:

|  | Top 1 Dx | Top 2 Dx | Top 3 Dx |
| --- | --- | --- | --- |
| DIAGNO | Progressive Supranuclear Palsy (PSP) | Parkinson's Disease (PD) | Multiple System Atrophy - Parkinsonian variant (MSA-P) |
| Neurologist | Progressive supranuclear palsy | Corticobasal degeneration | Frontotemporal lobar degeneration-tau |

Case summary: A XX-year-old right handed individual with a prior history of heavy alcohol use and marijuana abuse presented with slowing of their gait and difficulties with balance over a year. Their symptoms began with stiffness in their legs leading to balance issues and multiple falls that were limiting their ability to jog and perform other outdoor activities. Their partner reported gradual slowing of their movements, difficulty with posture and balance and loss of automatic movements including blinking and smiling.

They had no significant past medical history. Their medications included an anti-anxiety medication and multivitamins. There was no family history of similar symptoms.

Over the next year, they began to have a reduction in their verbal output without any word finding difficulties (they were previously known to be a “big talker”). Their partner also expressed that they appeared severely depressed and anxious; they also felt that the patient had withdrawn from their normal activities of daily living. They were having difficulty with organization and multitasking and required assistance with their ADL’s. They reported increased difficulty with dressing and showering due to increased stiffness in their extremities.

On initial neurological exam, they were alert and cooperative, were able to speak in fluent sentences with mild to moderate dysarthric speech. Their general cognitive functioning was notable for some slowing, but in general they had good cognitive performance with 30/30 on mini-mental state examination and 0 errors on the Blessed Dementia Scale. They had a masked face with a paucity of movements. Cranial nerve exam revealed restriction of upgaze on extraocular movements, but otherwise good voluntary saccadic eye movements and smooth pursuit eye movements and good elevation of the palate without evidence of tongue weakness or fasciculations. They had notable axial rigidity along with moderate leg rigidity and mild arm rigidity, predominantly of the left side, with a mild cogwheeling component. Stance was stiff and gait was mildly wide based with a cautious and short stride with reduced arm swing. Sensory exam and deep tendon reflexes were unremarkable. Diagnostics were performed.

Over the next XX years, they continued to have increased falls, some of which were severe enough to prompt emergency room visits. They had increasingly prominent anxiety with rumination on death by choking at night, agoraphobia, increasing difficulty with swallowing and cough, and increasing slowness in their gait and mild cognitive difficulties. They reported often losing their train of thoughts and their continuing symptoms were very distressing to them. Repeat MMSE revealed 28/30 and 2 errors on the Blessed Dementia Scale. Their exam was now notable for a weak cough and gag reflex, restriction of both upgaze and downgaze but still had good voluntary saccadic eye movements. Over the next year, their swallowing difficulties continued to worsen and they eventually required a PEG tube placement for nutritional support. They were hospitalized several times for aspiration pneumonia.

At age XX, they presented with jaundice and elevated liver enzymes, secondary to biliary obstruction from a possible pancreatic mass. No tissue diagnosis was obtained but they had a biliary dilation procedure for symptom management. Their neurologic condition had continued to decline. They were now barely able to stand with assistance, and moved their right leg, but required assistance with their left leg. Their exam was notable for markedly reduced blink rate and paucity of movements. Cranial nerve exam revealed some elevation of the palate but with poor tongue coordination; weak gag and cough. Their mouth would remain open. Extraocular movements were completely gone. They had severe leg rigidity (L>R) with severe L arm rigidity with dystonia, but still very mild R arm rigidity. Their family opted to pursue palliative care. They died at age XX and an autopsy was performed.

**Example 13:**

Neuropathological definitive diagnosis: Spinal Muscular Atrophy (SMA)

DIAGNO/Neurologist diagnoses:

|  | Top 1 Dx | Top 2 Dx | Top 3 Dx |
| --- | --- | --- | --- |
| DIAGNO | Spinal Muscular Atrophy (SMA Type 1) | Congenital Myopathy | Mitochondrial Myopathy |
| Neurologist | Spinal muscle atrophy (SMA) type 1 (Werdnig-Hoffman's disease) | / | / |

Case summary: A XX-year-old patient presented to the MGH Emergency Department from Logan Airport, where the patient had just arrived with their family from Puerto Rico. The patient was admitted to the Pediatric Intensive Care Unit for management of desaturations during the flight and for coordination of care for their chronic medical issues.

The patient had been born at full term following an uncomplicated pregnancy (7.4 lbs and 27.5 inches). Prenatal ultrasounds and screening tests were reportedly normal, although access to medical records was limited given the then recent significant damage in Puerto Rico. According to the patient's mother, there were no perinatal complications and the patient went to the regular nursery. The patient's mother had first become concerned about the patient's development when the patient was XX months old and seemed to be making fewer movements than expected and was also not lifting or turning their head. The patient began babbling at XX-XX months. At XX months, the pediatrician felt that the patient was not meeting developmental milestones and the patient was referred to a physiatrist. A combination of clinical findings, imaging and laboratory studies established a diagnosis.

The patient continued to have delayed development and never achieved head control, nor was the patient able to roll, sit, or crawl. At XX months, the patient developed pneumonia, requiring hospitalization and eventually leading to tracheostomy and G-tube placement. The patient had been ventilator-dependent since that time. The patient's respiratory function slowly declined over time, resulting in multiple hospitalizations in Puerto Rico for respiratory issues, most recently two weeks prior to the patient's arrival in Boston. The family had planned to move to Boston to seek further medical care for the patient, but the move was expedited in the setting of the hurricanes and lack of reliable availability of electricity for the patient's ventilator.

On exam at the time of the patient's arrival in the Pediatric Intensive Care Unit, the patient was noted to be non-dysmorphic. The patient's pupils were equal, round and reactive to light and accommodation, with no ptosis and the patient's extra-ocular movements were full. The patient had minimal voluntary jaw movement, along with significant contractures. The patient's face activated symmetrically and sensation was intact in V1-V3. The patient's hearing was grossly intact to tuning fork. The patient could not turn their head or shrug their shoulders. The patient had severe, diffuse contractures of their upper and lower extremities and exhibited no head or truncal control, requiring a chair for support with sitting. The patient displayed minimal voluntary movement of the limbs or trunk along with jaw contractures and significant facial weakness. The patient's muscle tone and bulk were profoundly diminished, including severe tongue atrophy and fasciculations. Sensation was grossly intact light touch and vibration throughout. The patient's cognitive status was difficult to assess given their motor limitations. The patient communicated only via facial expression and by fixing their gaze on objects of interest, such as the television.

Of note, the patient's younger sibling was 1-5 years old when the family moved to Boston. The sibling had initially exhibited normal development before experiencing loss of skills, and the sibling was no longer able to walk or support themselves although they did not require mechanical ventilation. There was no other known significant family history.

The patient was hospitalized multiple times over the following year, primarily for worsening respiratory issues that persisted despite multiple changes to the patient's trach and ventilator settings. The patient experienced multiple respiratory tract infections as well as other complications of their illness, including lower extremity fractures. Given the patient's declining respiratory status despite multiple interventions, the decision was made to redirect care to focus on patient comfort. Mechanical ventilation was discontinued. The patient died shortly afterwards and an autopsy was performed.

**Example 14:**

Neuropathological definitive diagnosis: Spinocerebellar Degeneration (Spinocerebellar Ataxia, Type 2)

DIAGNO/Neurologist diagnoses:

|  | Top 1 Dx | Top 2 Dx | Top 3 Dx |
| --- | --- | --- | --- |
| DIAGNO | Spinocerebellar Ataxia (SCA) | Multiple System Atrophy (MSA-C for cerebellar variant) | Friedreich's Ataxia |
| Neurologist | Spinocerebellar ataxia | ALS with motor neuron disease | Huntington's disease |

Case summary: A XX-year-old patient started to develop some intermittent slurring of their speech and difficulty with their balance. They were otherwise healthy and active. They were an avid hiker, were part of the National Park Service Search and Rescue and were a skilled mountain biker, competing at the national level. They did have a history of intermittent muscle cramps. At age XX, they had a concussion with no recollection of events for at least XX to XX hours.

At age XX, they developed difficulty with ascending and descending stairs, and difficulty keeping pace with their friends. They described some trouble carrying objects. Around that same time, their handwriting started to fail and speech became more slurred. They were also having some choking events with liquids and solids - small pieces of foods such as lettuce would get stuck and cause them to have significant coughing spells.

Their family history included an illness in their mother with onset in her late-XXs that affected her gait and balance. She died at age XX. Their maternal grandfather had an illness that resulted in impaired gait and speech considered to be MS. He died in his early XXs. The patient had completed college and had worked as a stone mason. They did not smoke and typically drank 2-3 beers on the weekends.

When they were XX years old, they started to have further difficulties with their gait and started using a cane. One year later, they reported some difficulty with reading and seeing small print. They also reported some troubles with their memory. They described a new aversion to heights and feelings of nervousness around crowds of people. They denied presyncopal episodes, pain, paresthesias or vertigo.

Their exam at that time was notable for a digit span of 5 numbers forwards with repeated attempts, and 3 numbers backwards. They named 24 animals in one minute and five words starting with the letter F. Speech was dysarthric and mildly slowed, but understandable. Their visual acuity was normal. Saccades were slow. Face was symmetric and facial sensation was intact. Their palate and tongue moved in the midline. Their tone was decreased in the arms and legs. Their strength was full throughout. Reflexes were absent in the lower extremities and at the brachioradialis, 2 at the triceps bilaterally, and trace at the biceps. The left plantar response was extensor and the right was flexor. There were no release phenomena. There was absent vibration sense in the toes and ankles bilaterally. Position sense was decreased in the toes bilaterally. Sensation to pin-prick and light touch were normal. They were able to walk with a wide stance but showed considerable staggering and slightly reduced speed. They took several steps to turn. They had swaying with Romberg that was exacerbated with eye closure. Heel-to-shin testing produced jerky movements in the axis; there was decomposition of movement with drawing a circle. There was impairment of rapid tapping and alternating movements. Finger-to-nose was mildly dysmetric. Their drawing of a spiral showed prominent hypermetric swerves. A range of laboratory studies were performed; imaging and other studies were abnormal. Medical management was begun.

At age XX, they developed progressive trouble with their memory, forgetting the names of birds (very unusual as they were an avid bird watcher). They would forget appointments and began to have difficulty recalling how to perform tasks. On exam, they were able to spell ""world"" forwards, but not backwards (""d-l-u-o-w""). Their eye movements showed severe slowing during both attempted saccade and pursuit. The eyes were quiet at rest without nystagmus. Their up gaze was mildly limited; down gaze was preserved. Finger-to-nose testing showed significant endpoint dysmetria with oscillation at the elbow. They were able to walk with assistance but were quite unstable. Truncal titubation was present with both standing and walking.

At age XX, they began to develop significant anxiety, manifesting as frustration and intermittent rage. Their memory continued to fail and their family reported that they were no longer able to hold a conversation. They lost bladder control and required assistance with the majority of their ADLs. By age XX, choking became more of an issue, with some foods eliciting significant coughing. Their exam showed near complete ophthalmoplegia except for minimal vertical gaze; in the horizontal plane, they turned their head to the target with eyes fixed at a mid-position. There is no optokinetic nystagmus. With maximal assistance, they were able to stand and walk about 20 feet. There was no notable tremor.

At age XX, they were admitted to an outside hospital for aspiration pneumonia -- nonverbal with minimal spontaneous movement. They died shortly after initiation of hospice care and an autopsy was performed.

**Example 15:**

Neuropathological definitive diagnosis: Metastatic breast cancer

DIAGNO/Neurologist diagnoses:

|  | Top 1 Dx | Top 2 Dx | Top 3 Dx |
| --- | --- | --- | --- |
| DIAGNO | Metastatic Brain Tumor (Breast Cancer) | Paraneoplastic Syndrome | Primary Central Nervous System (CNS) Lymphoma |
| Neurologist | Metastatic breast cancer with metastasis to the brain | Intraparenchymal hemorrhage | Reversible cerebral vasoconstriction syndrome with subarachnoid hemorrhage |

Case summary: A XX-year-old left-handed patient was in their normal state of health when they began to develop forgetfulness and fatigue over a few months. Additionally, their partner and co-workers had noticed subtle personality changes – they felt the patient was less engaged and energetic. Within the month leading up to their presentation, the patient had begun to experience severe headaches characterized by sharp pressure in their head that would shoot down their back/leg followed by vomiting with relief of symptoms. The patient also reported occasional episodes of right leg weakness/sensory changes and occasionally felt that their right peripheral vision was not intact.

Their history was notable only for BRCA2+ ER/PR+, HER2 negative breast cancer diagnosed three years previously. The patient had been treated with mastectomy, followed by radiation therapy and was maintained on tamoxifen.

Due to their persistent symptoms, the patient presented to an OSH. Physical exam was unremarkable at this time and an imaging study was performed. Given the results of this study, the patient was transferred to MGH where a therapeutic intervention was undertaken. Following this, the patient had improved symptoms, but was experiencing persistent fatigue requiring afternoon napping, and complained of right facial numbness in the V1-V3 distribution, as well as decreased hearing in their right ear. Their exam at this time was notable for recall of 3 of 3 items after 1 minute, but only 1 of 3 after 5 minutes, even when given multiple choice options. Hearing was slightly decreased on the right, and there was decreased sensation to light touch in the V1-V3 distribution. Finger-to-nose dysmetria was present, right greater than left. Additional therapeutic interventions were initiated.

One month following their presentation to MGH, additional therapeutic interventions were undertaken. Nine months following presentation the patient continued to do well, only experiencing occasional ‘painless’ migraines characterized by slightly blurred peripheral vision which lasted a few minutes and resolved with acetaminophen.

Nine months later, although the patient remained symptom-free, imaging studies were concerning and additional therapeutic intervention was begun. Their clinical status remained stable.

One year later the patient experienced an episode of right facial weakness and aphasia lasting 20 minutes prompting admission to MGH. On admission the patient was alert, and oriented, but was inattentive and unable to state the days of the week backwards. The rest of their exam was notable for intention tremor with the left hand on finger-to-nose testing. An additional therapeutic intervention was undertaken and following this the patient was experiencing waxing and waning mental status associated with visual hallucinations, delusions, and inattentiveness. Later that evening their exam worsened and the patient was poorly interactive, non-verbal, and not following commands. Their eyes opened to voice only, pupils were equal and reactive, but with intermittent right gaze deviation. There was bilateral upper extremity localization to noxious stimuli. Imaging was obtained and therapeutic interventions were sought with little effect. The patient was transferred to the ICU and made CMO five days later given their grim prognosis. The patient died three days later (age XX, roughly three years after presentation to MGH) and an autopsy was performed.

Supplementary Note 5. Fifteen Examples of Cases where DIAGNO was Correct and Neurologist was Incorrect.

**Example 1:**

Neuropathological definitive diagnosis: Neuronal Ceroid Lipofuscinosis Type 2 (CLN2)

DIAGNO/Neurologist diagnoses:

|  | Top 1 Dx | Top 2 Dx | Top 3 Dx |
| --- | --- | --- | --- |
| DIAGNO | Neuronal Ceroid Lipofuscinosis (NCL) | Leigh Syndrome | Alpers-Huttenlocher Syndrome |
| Neurologist | Batten Disease | Lafora disease | Juvenile metachromatic leukodystrophy |

Case summary: A XX-year-old patient started having difficulty interacting with their peers. They had been delivered at full term, weighing 9 lbs, to a healthy G1P1 mother after an uncomplicated pregnancy. They met all milestones until age XX when they were thought to have speech delay by their parents although formal evaluation placed them within the normal spectrum. In the months after this, they developed increasing separation issues as well as hyperactivity, impulsivity and inflexibility both at home and at school. EEG and imaging performed at age XX were normal. Starting at age XX, they became less social. They had difficulty playing with other kids as they did not have conversations with their peers but instead resorted to crying and throwing toys. Their parents often had to feed them because otherwise they would not eat. Seizures began at age XX, with four patterns: (1) tonic clonic movements R>L, lasting 2-5 minutes on average, associated with incontinence, preceded by aura of staring, giddiness, dizziness, abdominal pain and headache; (2) tonic clonic movements without aura, lasting 2-5 minutes on average; (3) ""spacing out"" or ""zoning out""; and (4) drop attacks without warning and loss of consciousness. Multiple medications were tried before moderate seizure control was obtained. There was no regression of milestones. Shortly after the onset of their seizures, they started having visual problems. Their parents noticed that they were watching TV and reading books in very close distance. Ophthalmology evaluation revealed visual acuity of OU 20/125 and ""symmetric retinal changes in the fovea of both eyes which were suggestive of a chronic degenerative process"" and bull's eye maculopathy. No other studies were performed.

Past medical history included cutaneous mastocytosis, recurrent ear infections status post bilateral ear tube placement, and a history of head trauma without loss of consciousness. Family history was notable for migraine (mother), febrile seizures (paternal aunt and cousin), bipolar disease (paternal great uncle) and depression (paternal great grandfather). The family heritage included East European (Russian, Poland), Italian, English and Scottish. They attended kindergarten where they received PT/OT/ST.

They were first seen at MGH when XX years old. On exam, their head circumference was 52cm. They were well groomed, very pleasant with no apparent distress. On mental status exam, they appeared very active and were running around the exam room but eventually they settled down in the corner of the room and watched a video for 10 minutes. They placed the portable video player fairly close to their eyes and were moving their head from left to right during the viewing. They spoke 4-5 word sentences and were able to follow commands. On cranial nerve exam, pupils were equal, round, and reactive to light; there was no ptosis, and extraocular movements were intact without nystagmus or saccadic breakdown; face, palate elevation and shoulder shrug were symmetric; hearing appeared intact; there was no dysarthria. Motor exam revealed full strength with normal tone. Deep tendon reflexes were 2+ and symmetric bilaterally at the triceps, biceps, brachioradialis, quadriceps and gastrocnemius/soleus. Toes were down bilaterally. Sensation was intact for light touch throughout. They did not have truncal ataxia. Their gait appeared normal, running a fair distance without difficulty. Repeated imaging 3 months prior to this assessment was abnormal, including abnormal spectroscopy. LTM at the same time was abnormal. Serum lactate was abnormal; a wide range of other biochemical studies were performed.

They continued to have decline in their vision and motor skills, becoming legally blind at XX years. They participated in an experimental trial at age XX, without evidence benefit. By the age of XX they were no longer walking. They had a G-tube placed. Their speech output was confined to ""knock knock"" as in the joke and ""hungry"". Multiple medications were tried in order to control their seizures and myoclonus, but without success.

XX years later, on physical exam, their weight was 58 kg and they sat in a wheelchair. They had frequent brief facial and upper extremity myoclonic jerking every 30 secs to a min. They did not open their eyes to stimulation and their eyes were dysconjugate. Their hearing remained intact. Face looked symmetric. They had atrophy and increased tone in upper and lower extremities. There was bilateral Hoffman's. They had brisk reflexes 4/4 throughout, with sustained ankle clonus. Toes were mute. During the course of the hour long visit, occasional brief myoclonic jerks with tonic posturing, especially of lower extremities, were noted. These were not stimulus-sensitive and there was no evidence of abnormal startle.

Several months later, they were referred to palliative care, which was provided at home. A few months later, they had breakthrough seizures and episode of status epilepticus. They died a few days later, age XX, and an autopsy was performed.

**Example 2:**

Neuropathological definitive diagnosis: Hereditary Sensory & Autonomic Neuropathy (HSAN) Type I

DIAGNO/Neurologist diagnoses:

|  | Top 1 Dx | Top 2 Dx | Top 3 Dx |
| --- | --- | --- | --- |
| DIAGNO | Charcot-Marie-Tooth Disease (CMT) | Hereditary Sensory and Autonomic Neuropathy (HSAN) | Amyloid Neuropathy |
| Neurologist | Charcot-Marie-Tooth (CMT) hereditary neuropathy | Friedreich's ataxia | Adrenomyeloneuropathy |

Case summary: A patient began to notice sensory loss in both feet when the patient was in their late teens, which progressed over a few years to involve their hands. In their mid-XX (approximately XX years later), the patient began to experience motor weakness of the distal muscles of their hands and feet, which progressed over the next few years to the distal muscles of their arms and legs. The patient had a history of multiple hand ulcers partly due to their sensory loss with subsequent amputation of multiple fingers. The patient also had significant atrophy of their distal limb muscles. The patient had been walking with two ankle foot orthotics (AFO) since their mid-XX and at age XX began to require more functional support with a knee-ankle foot orthotic (KAFO) and a crutch to support their left arm. At that time, the patient was otherwise independent with most activities of daily living and had learned adaptive techniques e.g. holding utensils in between the digits of their hand when eating and using their computer keyboard primarily with their thumbs. The patient denied any significant pain in their limbs. The patient was not aware of any abnormalities in blood pressure, bowel or bladder dysfunction. The patient endorsed dyspnea on exertion, particularly when climbing stairs.

The patient's family history was notable for multiple family members with similar symptoms including their sibling, parent and multiple cousins. The patient's parent, who died at age XX, had similar symptoms and eventually required bilateral below knee amputations due to infected leg ulcers. A cousin who was also affected with similar sensory and motor changes also had some degree of pulmonary dysfunction.

At age XX, the patient's exam was notable for an intact cranial nerve exam. Their motor exam was notable for bilateral shoulder abduction is 5/5, biceps and triceps 4+/5, wrist extension and flexion 3+/5. Finger extension, finger flexion and hand intrinsic muscle strengths 1/5. In the lower extremities, hip flexion was 5/5, knee extension 4/5, knee flexion 3+/5 and ankle dorsiflexion, plantar flexion, and toe extension/flexion were all 0/5. Sensory exam revealed decreased vibratory perception below the knees and in the fingers. There was no pin prick or light touch perception distal to the mid forearm and below the knees, and was present in the proximal portions of their limbs but decreased. Diagnostic studies were performed.

Four years after the above evaluation (at age XX), the patient noted a palpable right neck lump for which eventually discovered to be 2.5 cm right neck mass from a moderate-to-well-differentiated invasive squamous cell carcinoma possibly arising from the tonsil. The patient was treated with chemotherapy with a targeted therapy and radiation therapy after which their oropharyngeal cancer remained stable. Six years later the patient underwent a routine EGD for their Barrett Esophagus and was noted to have high grade dysplasia and invasive adenocarcinoma of the distal esophagus. The patient was started on chemotherapy with low-dose carboplatin, taxol and XRT however post-treatment imaging unfortunately revealed extensive spread of their disease to lymph nodes in the chest, abdomen, retroperitoneum, and to their liver, bilateral lungs and left adrenal gland. Palliative Care was provided. The patient died at age XX and an autopsy was performed.

**Example 3:**

Neuropathological definitive diagnosis: Spinocerebellar Atrophy Type 6 (SCA-6)

DIAGNO/Neurologist diagnoses:

|  | Top 1 Dx | Top 2 Dx | Top 3 Dx |
| --- | --- | --- | --- |
| DIAGNO | Spinocerebellar Ataxia (SCA) | Progressive Supranuclear Palsy (PSP) | Multiple System Atrophy (MSA-C for cerebellar variant) |
| Neurologist | Multisystem atrophy - cerebellar type | Progressive supranuclear palsy | Parkinson's disease |

Case summary: A XX-year-old right-handed patient started to veer to the right when they walked. Over the next several years, their imbalance progressed since then to the point of requiring a walker. By their mid-XX’s they were falling (frequency not specified in the notes). Despite not noticing subjective weakness in their extremities, they developed trouble with fine dexterity (picking up small objects, taking the top off a water bottle, using food utensils) and their writing became untidy and progressively illegible. Their speech became affected a few years after symptoms onset, but swallowing function had remained relatively preserved. Over the decade after onset of their difficulties, they developed diplopia for which they had two corrective surgeries with suboptimal results. They reported occasional urinary accidents for which they wore a pad for safety. They had no major cognitive concerns, but they became easily distractible and could not pay full attention to the task at hand. They also had noted pins and needles in the left leg (these were present prior to their treatment for breast cancer).

Their medical history was significant for a carcinoma of the left breast at age XX s/p lumpectomy, chemotherapy, and radiation therapy; and a traumatic brain bleed secondary to a motor vehicle accident at age XX managed without surgery. Their mother had begun falling when in her mid-XXs and was subsequently wheelchair dependent. Their mother had 5 sisters -- one was in a wheelchair, and their children had similar complaints. The patient was 1 of 8 siblings; 4 of them (3 individuals and 1 individual) also had similar difficulties. Their 4 children were all healthy at the time.

They were seen at MGH Neurology at age XX. They were alert and engaged throughout the visit, although their story was not always as logically presented as possible. They were aware of current events in the news. Their visual fields were full to confrontation testing. The pupils were 3 mm and reacted to 2 mm directly and consensually to light, and with accommodation. Early cataracts obscured adequate fundoscopy. When considering their eye movements, the eyes were quiet at rest, with mild saccadic intrusion into pursuit movements both horizontally and vertically. Saccades were almost normal, and they had gaze-evoked direction beating nystagmus in the horizontal plane. Their ability to suppress the vestibulo-ocular reflex (VORC) was impaired. Facial sensation and movements were full and normal. Hearing was intact to high and low tones. The palate and tongue moved in the midline. There was mild cerebellar dysarthria, more so with rapid consonants. Finger dexterity was slowed and dysrhythmic, L>R. Finger-to-nose testing showed end-point tremor with occasional oscillation at the elbows as well. Rapid finger following had overshoot, L>R. Rapid alternating movements were dysrhythmic. There was proximal overshoot when placing the heel on the opposite knee. Heel-to-shin testing was slowed in phases with an occasional superimposed jerking bilaterally. Gait was slowed and unsteady, with them taking only a few steps on their own. Romberg and tandem were not safe to test. The deep tendon reflexes were 2+ throughout, and no release phenomena were observed. There was normal appreciation of touch, pin-prick, position, and vibration. Double simultaneous visual stimulation was intact.

About a year after this visit, they had two episodes of confusion and unresponsiveness. They were seen at an OSH, and medical management was begun following a series of studies (Information regarding exam and studies is not available). Following intervention no subsequent episodes were reported.

They were seen again about 6 months afterwards (now age XX). They had fallen from bed the previous month, fracturing two vertebrae. At this visit, their XX-year-old child who accompanied them reported that they were having balance problems when they stood quickly and had become cautious as they walked. At this visit, the patient felt their symptoms were progressing. Their exam now showed more prominent saccadic intrusions in both planes. Their dysarthria was more prominent. Heel-to-shin testing showed side to side movements on the right and in-plane jerking with occasional superimposed lateral movements on the left. They were unable to walk at all.

They were not seen again at MGH. By report from their family, they had recurring aspiration pneumonias and decline in overall health. They died at hospice at age XX and an autopsy was performed.

**Example 4:**

Neuropathological definitive diagnosis: Spinocerebellar Atrophy Type ATP2B3 (X-Linked)

DIAGNO/Neurologist diagnoses:

|  | Top 1 Dx | Top 2 Dx | Top 3 Dx |
| --- | --- | --- | --- |
| DIAGNO | Spinocerebellar Ataxia (SCA) | Multiple System Atrophy (MSA-C for cerebellar variant) | Amyotrophic Lateral Sclerosis (ALS) |
| Neurologist | Progressive supranuclear palsy | Corticobasal degeneration | Multisystem atrophy |

Case summary: A XX-year-old right-handed patient noticed that they were walking “differently”, with a feeling that they were leaning somewhat forward when standing. They noted tinnitus with a high-pitched ringing, but no loss of hearing. About XX months later, they noted, while riding their bicycle, that they had difficulty judging distances and were afraid that they would crash into things. Subsequently, they had difficulty moving off and on the bicycle. They reported increased sensitivity to alcohol: a few beers resulted in their appearing intoxicated (very different from their baseline). Around age XX, their speech became slower and more “gruff” than previously. By the time that they sought an opinion at MGH, they needed to concentrate while walking, otherwise, they would lose their balance. They needed to hold onto the walls or furniture when walking, as it made them steadier to touch items. They were unable to turn their head to the side while walking, otherwise they would fall. They reported having recently seen an amateur video, where the image was jumping around the screen, and they remarked to their partner that that is how the world looked to them. There was also a decrement in the legibility of their handwriting, and they noticed that they were frequently dropping objects. In the few months immediately preceding their first appointment, they had a ""meltdown”, as they could not stop thinking about their deteriorating function.

Their medical history included BPH, erectile dysfunction (onset in their mid-XXs) and migraines. They were a nonsmoker and denied excessive alcohol use. They worked as a vice president of a publishing company. There was no history of neurologic disease in their immediate family.

On initial exam at MGH (age XX), their mental status was intact. There was no change in HR or BP when they shifted from sitting to standing. They had square wave jerks at rest and persistent downbeat nystagmus evoked mostly on bilateral gaze. They had saccadic intrusions with pursuits, as well as saccadic overshoots. Their speech was slow, but not obviously dysarthric. The remaining cranial nerves were normal. Their strength was full throughout. There were a few fasciculations in the forearms, a spastic catch in the right arm and increased tone in the right leg. The deep tendon reflexes were 2+ in the arms, 3+ at the knees and 2+ at the ankles bilaterally. The plantar responses were equivocal. They had positive crossed adductors, as well as supraclavicular and pectoral reflexes. The jaw jerk and Hoffmann reflexes were negative. Pinprick, vibration, and position were normal throughout. There was mild dysmetria and over-shoot in the upper extremities bilaterally. The heel-to-shin test revealed slowing of movement down the shin without loss of accuracy. There was some breakdown of circular movements with the right leg. Their gait was notable for reduced right arm swing, but otherwise normal stance and stride but they were unable to tandem. There was mild sway on Romberg maneuver.

Over the following years, they developed progressive gait instability and recurrent falls. When seen at age XX, there was a mild stare at rest, with microsaccadic oscillations in the horizontal and vertical directions. Laterally directed gaze produced direction-beating nystagmus with a downbeat component; their vertical gaze was better down than upwards. The pursuit movements showed saccadic intrusions. The saccades showed hypometria on some attempts and hypermetria on others, more in the horizontal plane than vertical, with refixation saccades. Their speech was moderately dysarthric but their words are otherwise understandable. There was mild high tone hearing loss L>R. The heel-to-shin testing shows in-plane dysmetria on the right with lateral movements on the left. Their finger-to-nose testing was slowed, with decomposition of movement and endpoint tremor bilaterally, L>R, and instability at the nose. Rapid alternating movements were slowed and awkward, left hand movements producing entrained movements on the right hand. Rapid finger following showed mild overshoot bilaterally. Their gait was wide-based, unstable, stiff-legged, high stepping, and slow.

They demonstrated dysphagia with both solids and liquids, and ultimately required a PEG. They had multiple hospitalizations for pneumonias. Their family noticed the emergence of problems with multi-tasking and short term memory, as well as increased obsessional thinking and impulsivity.

At age XX, their eye movements were slow in all directions. Their speech was moderately dysarthric with impairments in lingual, buccal and guttural sounds. They had fasciculations of the arms and calves. Their finger-to-nose testing was slowed with endpoint tremor bilaterally. Position and vibration sense were impaired in both great toes. They could not appreciate pinprick or cold temperature on the limbs, trunk or face. They could no longer walk and were wheelchair dependent. Given their overall decline, they transitioned to hospice. They died approximately XX years after their initial presentation to clinic (XX years after symptom onset) and an autopsy was performed.

**Example 5:**

Neuropathological definitive diagnosis: Alzheimer's Disease

DIAGNO/Neurologist diagnoses:

|  | Top 1 Dx | Top 2 Dx | Top 3 Dx |
| --- | --- | --- | --- |
| DIAGNO | Frontotemporal Dementia (with prominent language variant: Primary Progressive Aphasia - Semantic Variant) | Alzheimer's Disease Neuropathological Changes (ADNC) | Lewy Body Dementia (LBD) |
| Neurologist | Semantic variant of primary progressive aphasia (svPPA) | Corticobasal degeneration | Frontotemporal dementia with motor neuron disease (FTD-MND) |

Case summary: A XX-year-old right handed individual was seen because of word finding difficulty. The patient estimated that this had been going on for four years, but their partner and daughter had noticed a problem for only the past two years when the patient often stopped in mid-sentence, apparently blocked at finding the right word. The patient indicated that they knew what they wanted to say but just couldn’t get it out. The patient would employ circumlocution and paraphasias. For example, when the patient meant to say “tree”, they might say “leaf”. This was especially true with time and numbers where “30” might be stated as “300”. The patient indicated that they were slightly more forgetful than they used to be but their family failed to corroborate this point. They did point out that the patient was less quick in sequencing and following things in order, a deficit the patient admitted existed in what the patient and the family called “multi-tasking”. The patient remained with a pleasant, unchanged personality and still drove a car and handled money. The patient was independent in ADLs and traveled independently abroad and helped advise their daughter in her business.

The patient's past medical history included only asthma and high cholesterol. On initial exam, there was mild diminished hearing, but no focal neurologic signs. The patient performed 4/4 gestures to command and mimicry easily. The patient named only 2/4 common objects to confrontation, calling a bowtie a ‘square tie’ and being unable to name ‘watch strap’.

XX months later, naming impairment was evident both during discourse as well as on formal testing. Memory was impaired at the stages of acquisition and retrieval on verbal memory measures, but was generally within normal limits on a test of nonverbal memory. Memory retention appeared to be well preserved across all types of tasks. Phonemic fluency was normal/average, although semantic fluency was significantly impaired. Other aspects of language appeared normal, including repetition, comprehension, prosody, and written expression (although a few minor errors were noted). Performances on tests of attention and executive functioning were variable, as the patient performed normally on several measures, but demonstrated greater impairment on others. One self-reporting scale indicated mild depressive symptoms, although the patient performed normally on two other self-report measures of emotional functioning.

Subsequently, the patient's family described changes in behavior: erratic purchasing of items; the patient had always had had obsessive interest, but this was increased and the patient was newly focused on clocks. The patient also had developed a ""mean streak"" -- yelling at their daughter, loud screaming and swearing, which was completely out of character for the patient. The patient was also taking naps on a regular basis, which seemed to be increasing, but the patient did seem to function better after the nap. Late in the day after working all day, the patient often did not function very well at all. There had been no changes in the patient's eating behavior; the patient had always loved sweets and continued to eat them.

XX years later, the patient needed to use cues from a letter/word/picture book that their daughter made for them to 'practice' and would sometimes switch words (snow for rain, hot for cold). The patient was now having major fluctuations from day to day, sometimes surprising their partner with what they remembered or could do. The patient had become obsessed with certain foods and couldn’t make decisions about what to eat. The patient was increasingly funny, sometimes inappropriate, sometimes hurtful. The patient showed no loving emotion but would reciprocate when hugged. The patient was having trouble with shape orientation, ate only with a spoon and could not be alone.

On exam the patient had frequent word-finding difficulties and had a very difficult time repeating anything except bisyllabic single words. The patient could retrieve words a bit better with phonemic cues and now had problems with imitational limb movements (suggestive of transposition into the body space of the examiner). There were subtle problems with ideomotor praxis, particularly in the patient's left hand. The patient was unable to do a simple calculation problem and was unable to estimate the length of a person's spine.

XX years later the patient showed significant progression in symptoms with motor movements including moving feet, twitching hands, making throwing movements when watching basketball. The patient's mood was generally very happy (happier than they used to be), but could also get frustrated/angry easily, especially in conversations. The patient needed help with dressing, bathing, and hygiene, although the patient would say they could do it. The patient ate using their fingers. The patient was having difficulty reaching/grasping, and unintentionally throwing glasses and cups. The patient would often be incontinent of urine, especially at night. The patient only recognized people they saw on a daily basis and was extra affectionate. The patient's language impairment was now generally consistent with global aphasia, and the patient was unable to follow commands on neurologic exam. A year and a half later, XX years after symptom onset, the patient died and an autopsy was performed.

**Example 6:**

Neuropathological definitive diagnosis: Multiple Sclerosis

DIAGNO/Neurologist diagnoses:

|  | Top 1 Dx | Top 2 Dx | Top 3 Dx |
| --- | --- | --- | --- |
| DIAGNO | Amyotrophic Lateral Sclerosis (ALS) | Hereditary Spastic Paraplegia (HSP) | Multiple Sclerosis (MS) |
| Neurologist | Spinocerebellar ataxia (SCA, suspect Type 3) | Spinocerebellar ataxia with axonal neuropathy (SCAN1) | Cerebellar variant of multiple systems atrophy (MSA-C) |

Case summary: A XX-year-old left-handed patient developed numbness down the front of their left leg. The patient initially attributed these symptoms to sequelae of a superficial wound they suffered while crawling down a rusted fire escape, but the numbness continued after the wound resolved. The patient sought medical attention and, due to comorbid low back pain, a study was performed, resulting in treatment with physical therapy. However, the leg numbness continued, and a year and a half later, their left arm became numb. The patient had no neck pain and no history of trauma. Their employees noted that the patient looked like they were limping. The patient noted their handwriting had deteriorated. The patient also felt they were not as quick-witted as they had been in the past and felt their speech was off. Three years after initial symptom onset, the patient was referred for neurologic evaluation.

The patient had no significant past medical history and took only a herbal supplement for mood. The patient's mother had been diagnosed with a neurologic condition that left her with significant gait impairment. The patient also had a cousin with the same condition. The patient smoked one pack of cigarettes and drank one glass of wine per day.

At the initial neurologic exam, the patient had fluent speech without dysarthria and no apparent cognitive problems. Their visual acuity was 20/25, and their extraocular movements were intact. The patient had left-sided pronator drift with subtle weakness in triceps, wrist extension, and hip flexion. The patient had decreased vibratory sensation in both feet. The patient had hyperreflexia on the left with a positive Babinski. On coordination testing, the patient's left side was dysmetric on finger to nose testing with dysdiadochokinesia in their arm and leg. The patient could only take one or two steps on tandem gait. Diagnostic studies were performed and management was initiated.

One year after initiating treatment, the patient had also started a specialized diet, and they felt their dexterity and strength on the left were improved. The patient felt their prior speech difficulty had resolved completely. However, the patient had difficulty concentrating, felt fatigued, had decreased sexual drive, and co-workers had commented the patient was more irritable. An additional therapy was started. The patient's irritability improved but the fatigue continued, so yet another therapy was added and adjustments were made to the initially instituted treatment.

One year later, the patient's gait deteriorated to the point that they required a cane. The patient was experiencing aching muscle and joints. Because these symptoms were a possible side-effect of their ongoing treatment, alterations to their management were undertaken with initiation of a new multi-drug regimen. The patient had a mild skin reaction to this regimen, but their overall exam remained stable with only minor worsening over the next five years. At that time, the patient developed osteoporosis and one part of their multi-drug regimen was discontinued.

Unfortunately, a year later, the patient's balance, fatigue, and dysarthria worsened, and they began to have dysphagia and a hoarse voice, so their treatment regimen was changed to a single agent. After three years of dysphagia, the patient underwent further workup of their dysphagia and voice changes. Studies revealed a laryngeal malignancy; treatment for their neurologic disease was stopped. The patient received a range of treatments for the malignancy, and four years after establishment of this diagnosis, the patient transitioned to hospice. The patient died shortly thereafter (18 years after neurologic symptom onset) and an autopsy was performed.

**Example 7:**

Neuropathological definitive diagnosis: Midline glioma, H3K27M mutation

DIAGNO/Neurologist diagnoses:

|  | Top 1 Dx | Top 2 Dx | Top 3 Dx |
| --- | --- | --- | --- |
| DIAGNO | Progressive Multifocal Leukoencephalopathy (PML) | Paraneoplastic Syndrome | Brainstem Glioma |
| Neurologist | Metastatic cancer with brain metastasis | CNS lymphoma | Acute demyelinating encephalomyelitis |

Case summary: A XX-year-old individual presented because of headaches, diplopia and difficulty walking emerging over 6-7 weeks. The patient's headaches were accompanied by photophobia and nausea without vomiting, similar to previously reported migraines, but were now increasing in severity and duration. During a headache, standing exacerbated the head pain and the patient reported feeling lightheaded and off balance. Around the same time period, the patient began to experience double vision that was becoming more frequent. The patient described images on two images on top of each other that resolved with closing either eye, but was unable to tell if it was worse in any one particular direction of gaze. The patient's difficulty with balance and ambulation had eventually resulted in a fall down a flight of 3 steps with direct impact of the occiput onto stone steps, which brought the patient to medical attention. The patient felt that their left leg may have felt weaker over the last few weeks and contributed to the fall. Frequent hiccups were also reported. The patient had no history of recent illness, trauma, alterations in appetite or travel outside the country; the patient noted an unintentional weight loss that could not be quantified.

The patient's past medical history was significant for atrial fibrillation on anticoagulant medication, HLD, migraine headaches, gastric bypass, depression, PTSD and a sensorimotor axonal neuropathy. Work up of the patient's cardiac dysfunction included a TTE that had shown an EF 57% and symmetric LVH w/ PWT 15mm. The patient's idiopathic sensorimotor neuropathy had developed as stabbing pain in the feet XX years prior. NCS/EMG revealed primary axonal involvement that was stable on repeat testing across the next 2 years. Work up had included a normal SPEP, B12, ESR, CRP and Lyme titer. The patient was not diabetic. An ANA (1:640) and Sm antibody were positive and RNP negative. There were no additional findings of rheumatologic disease. The patient had found some relief on pregabalin. The patient had also lost their sense of smell XX years prior to presentation; there were some imaging abnormalities observed but no diagnosis was established and the deficit persisted.

On presentation, the patient was cognitively intact. Evaluation of the cranial nerves was remarkable for inability to abduct the left eye on leftward gaze and an abducting nystagmus in the right eye with an inability to adduct the left eye on rightward gaze. Additionally, limitation in upward gaze was noted with the left eye. Facial sensation and strength were preserved. On motor testing, strength was mildly reduced in the left upper and lower extremity and with an accompanying asymmetric increase in tone on the left. Reflexes were depressed diffusely and absent at the ankles. Toes were mute bilaterally. The patient's gait was described as lurching with veering to the left. Imaging and laboratory studies were obtained. CSF studies demonstrated glucose 68, TP 73, TNC 3 (tube1) and 1 (tube 4), 27-29% neutrophils, 22-57% lymphocytes, 40-14% monos; ACE, Listeria, and Syphilis antibody were negative. CSF IgG was high at 6.6 but no bands were seen on electrophoresis. Cytology did not reveal abnormal cells. Additional studies included serum ANA, RPR, Ro/La, NMO, Lyme, HIV, and ACE which were unremarkable. Additional imaging was performed. Medical management was begun and the patient was discharged home after 6 days in the hospital.

Over the next 2 weeks, symptoms showed no improvement with treatment. The patient was readmitted for further workup at which time their right-sided hand weakness had worsened, diplopia and hiccups had become more frequent and the patient complained of trouble swallowing, solids more than liquids. A diagnostic procedure was performed. During the hospital course, the patient had trouble managing secretions and was treated for a URI and PNA. Frequent apnea developed and the patient opted to not undergo any additional treatment, asking that their care be shifted to focus on comfort measures. The patient died about a week after this transition and an autopsy was performed.

**Example 8:**

Neuropathological definitive diagnosis: Huntington's Disease

DIAGNO/Neurologist diagnoses:

|  | Top 1 Dx | Top 2 Dx | Top 3 Dx |
| --- | --- | --- | --- |
| DIAGNO | Huntington's Disease | Progressive Supranuclear Palsy (PSP) | Frontotemporal Dementia (FTD) |
| Neurologist | Lewy body dementia | Frontotemporal dementia with motor neuron disease | Chronic traumatic encephalopathy |

Case summary: A XX-year-old individual was brought to an ED by their son because of altered behavior. The patient first drove from their house in Virginia to their son’s house in Massachusetts and upon arrival called their son and demanded they leave work and sign a power-of-attorney document. The son returned home to find the patient visibly shaken. The patient told their son that they were fearful for their life and for their family’s safety. The patient told their son that if they returned to their job, they would be arrested without trial and placed in prison for life. At this point because their behavior was so far from their baseline, they were brought to an ED. When asked about the circumstances, the patient reported that they had a “top secret” job with the government and could not discuss any further specifics. The patient also reported feeling “hyper” and endorsed decreased sleep and appetite, weight loss, and occasional headaches. The patient was admitted to a hospital. A diagnosis was confirmed and a medication was started.

The patient's medical history was remarkable for two TBIs: at age XX, the patient was hit by a windmill, leading to coma for 3 days, and at age XX, the patient was hit by a car, resulting in retrograde memory loss. The patient smoked more than 2 ppd for most of their life. The patient neither drank nor used illicit drugs. The patient worked as an Operations Research Analyst for the Air Force. The patient had a family history of neurologic disease. The patient's partner committed suicide 5 years prior.

Six months later, the patient presented to a specialist. The patient felt that the medication was helping, but believed that they were not as “quick” as they used to be and that their attention span had decreased. The patient reported some mild gait changes and sporadic calf fasciculations that improve with walking as well as occasional nocturia. On exam, saccades were slow to initiate and were slow in velocity; saccadic intrusions were present. Speech was slow but clear. The patient required verbal auditory self-cues to perform the Luria test, but was able to do 4 in 10 seconds. The patient had mild to moderate bradykinesia and abnormal movements of the eyes and mouth. Gait was slightly wide-based with a slightly diminished arm swing. The patient had difficulty performing the tandem maneuver, with 3 deviations from a straight line. The patient underwent formal testing, which demonstrated borderline performance in visual and spatial learning and slow psychomotor speed.

Over the following XX years or so, the patient complained of unrelenting fatigue and progressive weight loss despite eating up to 4000 kcal/day. The patient also experienced worsening arthritis of their hands and developed chronic back and shoulder pain. Serial exams showed slowly worsening dystonia, but otherwise remained stable. The patient's back pain had worsened, prompting spine imaging that showed an abnormality. On exam, the patient had weakness of hip flexion at the LLE, significant kyphoscoliosis and had new abnormal movements of the bilateral lower extremities with stable abnormal movements of the eyes and mouth. The patient's exam was otherwise unchanged.

The following year (now XX years since initial diagnosis), the patient developed a feeling of heaviness in the legs as well as cramping of the calves and thighs and occasional electrical sensations down the back of their legs. The patient continued to endorse ongoing fatigue and weight loss. The patient's exam was notable for abnormal movements of the eyes, mouth, and bilateral arms and legs but was otherwise unchanged.

One year later (now XX years since diagnosis), the patient developed cramping and tingling of the hands, as well as difficulty using their hands: trouble buttoning their shirt, shaving, and picking up pills. The patient also reported an occasional sense of choking. Fatigue and weight loss worsened and the patient developed occasional reflux and chest pains. A few months later, the patient presented to an outside provider with new symptoms. An extensive work-up was done and a new non-neurologic abnormality was found. The patient was admitted to hospice for further care. The patient died at the age of XX, eleven years after presentation to the ED, and an autopsy was performed.

**Example 9:**

Neuropathological definitive diagnosis: Multiple System Atrophy (MSA-P)

DIAGNO/Neurologist diagnoses:

|  | Top 1 Dx | Top 2 Dx | Top 3 Dx |
| --- | --- | --- | --- |
| DIAGNO | Multiple System Atrophy (MSA-P) | Parkinson’s Disease (PD) | Progressive Supranuclear Palsy (PSP) |
| Neurologist | Progressive supranuclear palsy | Corticobasal syndrome/corticobasal degeneration | Parkinson's disease |

Case summary: A XX-year-old individual was seen because of difficulty walking. The patient reported that two years earlier they developed right arm and neck pain with numbness in their right thumb. While this was being managed conservatively, they developed decreased dexterity with their right hand. Their symptoms were stable until a few months before the clinic visit, when they additionally developed a sensation of burning/""prickliness"" in the right upper leg which gradually spread to the right foot over ensuing weeks. The patient also became more unsteady described as clumsiness in their right foot while walking and when trying to get up from sitting. Their partner had noted their voice becoming softer. There had been no changes in mood or personality. The patient had seen an outside physician and been prescribed a medication with limited benefit although it had been discontinued because of a rash. Their past medical history was minimal, with GERD (on a generic medication). The patient had smoked for 8 years but quit >XX years earlier. There was no history of excessive alcohol intake or other drugs.

At initial examination, cognition was intact. Their speech was fluent but mildly hypophonic. Extraocular movements were full except for mild upgaze limitation. Saccades and smooth pursuit were intact. Decreased blink rate was noted in both eyes. Axial tone was mildly increased, and there was increased tone (1-2+) right greater than left and upper extremity greater than lower extremity. Bradykinesia was present in the right upper and lower extremity. There was no rest or intention tremor; finger-nose-finger and sensory exams were normal. Their strength was preserved throughout apart from slight right nasolabial fold flattening. Their gait showed stooped posture with shortened stride length, decreased arm swings (right > left). The patient passed the 'pull test' but used extra steps for their turns. Medical management was adjusted.

The patient did not subjectively notice improvement although on follow-up visit their bradykinesia was mildly improved in the right upper extremity. Six months after initial evaluation, they began to complain of more difficulty walking with increased limping and dragging of the right leg. The patient also complained of constipation. An additional medication was added to their regimen with some improvement in walking.

The patient underwent a sleep study that showed snoring and severe periodic limb movements of sleep. Although not captured during the study, the patient complained of waking up at night acting out their dreams and talking in sleep. They also developed increasing anxiety and fatigue requiring additional medications.

Two years after starting therapy, the patient complained of progressive difficulty walking as well as increasing urinary frequency and urgency (getting up as many as 4-5 times per night). Examination did not show tremor at rest or during action. Tone was mildly increased axially and on the right side. Extraocular movements did not have any restriction. Mild bradykinesia was noted bilaterally right greater than left. The patient had marked impairment of speech with both dysarthria and moderate hypophonia. They had difficulty rising from a chair, requiring use of their arms. Their gait was mildly stooped, slow and shuffling with en bloc turns. The patient had moderate retropulsion on 'pull testing'.

One month later, the patient was hospitalized for near syncope while walking. Systolic blood pressure was in the 80s to 90s responsive to IV fluid. The patient was discharged after addition of a medication to their regimen. On follow-up, their systolic blood pressure continued to be in the 90s though their systolic blood pressure dropped by 6 points when standing from sitting.

The patient started experiencing multiple falls with deteriorating gait. They also had frequent urge for urination and were found to have neurogenic detrusor overactivity on urodynamic studies requiring intervention. Over the next two years, their gait and speech particularly continued to deteriorate. A trial of an adjunctive medication provoked visual hallucinations. Their handwriting became illegible. Six years after first evaluation, the patient moved into an assisted living facility requiring help with all daily activities.

The final visit, seven years after their first assessment, revealed the patient to be alert but very frail appearing. Marked axial rigidity was present. The patient was alert and oriented but their speech was barely intelligible because of dysarthria. Marked blunting of facial expression was present. Their eye movements were described as intact with saccadic breakdown. The patient was markedly rigid and bradykinetic for repetitive movements. They were able to stand with assistance and could take 1-2 small steps with support. The patient was transitioned to hospice care. The patient died at age XX (slightly more than 7 years after their first visit) and an autopsy was performed.

**Example 10:**

Neuropathological definitive diagnosis: Glioblastoma, IDH-wild type

DIAGNO/Neurologist diagnoses:

|  | Top 1 Dx | Top 2 Dx | Top 3 Dx |
| --- | --- | --- | --- |
| DIAGNO | Glioblastoma Multiforme (GBM) | Primary Central Nervous System Lymphoma (PCNSL) | Progressive Multifocal Leukoencephalopathy (PML) |
| Neurologist | RCVS | Ischemic strokes | Cerebral amyloid angiopathy |

Case summary: A XX-year-old patient presented to an outside emergency department after an episode of speech impairment. Earlier that day while at work the patient had a headache that began with pain behind their right eye. Later when the patient came home from work they were talking with their partner and the partner noticed that the patient was ""speaking with garbled words and was making no sense"". The partner tried to have the patient spell their words and the patient was able to write their son's name but then dotted a 'w' as if it were an 'i'. These symptoms lasted about 20-30 minutes and then mostly resolved. The patient had experienced a headache off and on for the last several days and had been feeling more stressed with taking care of their XX-year-old father. The patient denied nausea, vomiting or visual changes. The patient did not have any fever, chills, numbness or weakness.

The patient's past medical history was significant for obesity, hypertension, fibromyalgia, depression, arthritis, migraine, obstructive sleep apnea and GERD. The patient had a remote history of seizure in infancy and then again in the 1980's. The patient had a concussion in their childhood. The patient also had a history of herniated disc of the lumbar spine and multilevel neural foraminal stenosis. The patient was a nonsmoker and rarely drank alcohol.

Physical examination from this initial encounter was not available for review. The patient was admitted to the hospital as imaging demonstrated an abnormality and a therapeutic intervention was initiated. Three days later the patient was discharged from the hospital, their presenting symptoms having resolved.

Two weeks later at a follow-up visit for the recent hospital admission, the patient was doing well apart from a mild headache and some increased reflux esophagitis. Physical examination demonstrated an obese, short individual with normal mood and affect. The patient's judgement and thought content were normal. Neurologic exam demonstrated that the patient was oriented to person, place, and time. The patient had normal muscle tone, and normal reflexes. There was no cranial nerve deficit. Coordination and gait were normal.

Three months later imaging demonstrated a worsening abnormality requiring ongoing treatment. Two months after this at a follow-up visit, the patient's presenting concern was ongoing headache. The patient's headache at this time was described as typically dull, nonspecific, with a sense of pressure, tightness, and fullness in their neck and around their head, and occurring daily. The patient did not confirm any migrainous components currently, although that had been an issue in the past, with episodic sound- and light-sensitivity occurring once or twice every other month. On exam, the patient was alert, oriented, and attentive. Speech was hesitant with word finding and some comprehension issues but no dysarthria. Visual fields were full. Cranial nerve exam was normal. The patient had normal bulk and tone. There was no pronator drift. Upper and lower extremity strength was 5/5. Sensation was intact. Reflexes were normal in the extremities. Both toes were down-going. Appendicular coordination was intact. Gait was slightly wide based but stable. Imaging showed worsening findings and therapeutic interventions were continued.

Eight months after this the patient presented to the ED for an episode of severe reflux but left without being seen. Two months later the patient was admitted for a diverticulitis flare. Two months after this at a follow-up visit the patient was feeling depressed, tired, irritated, feeling weak and having some speech difficulty. On exam, the patient was alert and oriented but demonstrated some expressive and receptive aphasias. Cranial nerve was normal except for loss of vision of the right lower quadrant bilaterally. On reflex exam the patient had 1+ brachioradialis reflex bilaterally and 2+ patellar reflexes bilaterally. Sensory and motor exams were normal. Coordination and gait were normal. Imaging showed worsening findings and an alternative approach was discussed.

The patient died one month later, 1.5 years after their initial presentation, and an autopsy was performed.

**Example 11:**

Neuropathological definitive diagnosis: Progressive Supranuclear Palsy (PSP)

DIAGNO/Neurologist diagnoses:

|  | Top 1 Dx | Top 2 Dx | Top 3 Dx |
| --- | --- | --- | --- |
| DIAGNO | Progressive Supranuclear Palsy (PSP) | Multiple System Atrophy (MSA-P, parkinsonian variant) | Corticobasal Degeneration (CBD) |
| Neurologist | Huntington's disease | / | / |

Case summary: A XX-year-old individual began having frequent falls after being treated for breast and colon cancer. Two years earlier, the patient noted that their right nipple was inverted and painful. The patient was diagnosed with breast cancer requiring a radical mastectomy and resection of lymph nodes with radiation and chemotherapy (adriamycin, Cytoxan and tamoxifen). The patient did well, but the following year was found to have colon cancer on routine colonoscopy which was successfully resected. Beginning about a year after this, the patient began to have difficulty with their gait, falling frequently and hitting their head ""100 times."" The patient would not fall in a particular direction, but had difficulty catching themselves when they did fall. The patient reported that they would lose their balance easily, with falling provoked by things as simple as bumping into a door. The patient began to use a cane for stability.

Apart from the two malignancies, the patient's only prior medical problem was hypertension. When the patient first presented because of the falls, their only medications were amlodipine, naprosyn, and baby aspirin. The patient previously worked as a mechanic for photocopiers, had 50 pack-year smoking history and occasional alcohol use. The patient's father died at XX with multiple neurologic problems including TIAs, strokes, and carried a diagnosis of Parkinson disease, with difficulty walking and shaking resulting in becoming wheelchair-dependent. A paternal uncle had a similar course and diagnosis. The patient's two children were in good health. Additionally, there was a strong family history of multiple cancers.

The patient began their care with a local neurologist, and a diagnostic study suggested an abnormality prompting a therapeutic intervention. As the patient developed a rash and showed no clear improvement this was discontinued. The patient was seen at MGH a year later for a second opinion. At that time, the patient's exam was notable for possibly slurring of their speech, mild difficulty with praxis, saccadic pursuit, left end-gaze nystagmus, and mild increase in tone in upper and lower extremities with contralateral reinforcement. Finger-nose-finger and heel-to-shin were dysmetric at the endpoints (more so on the left than the right). The patient's gait was wide based and fairly unstable requiring assistance. Romberg sign was absent. Sensory exam and reflexes were normal. Numerous diagnostic studies were performed, with only a single abnormal result.

The patient was re-evaluated two years later when they were no longer able to stand or walk without assistance and required use of an electric scooter. The patient had tremors almost constantly and had worsening of their speech. The patient would choke with any kind of food and developed some urinary urgency. The patient's exam revealed BP of 200/104 while sitting and 140/90 (asymptomatic) after one minute of standing. The patient had moderate dysarthria and occasional variable large amplitude movement of their upper and lower extremities. The patient had mild overshoot in the upper extremities and impairment of rapid alternating movements (still greater on the left than the right). The patient was unable to walk without assistance. The patient was started on a medication unfortunately without benefit and additional studies were performed.

The next year, in follow-up, the patient was found to have square wave jerks at rest with severe dysarthria, slowing of speech and difficulty maneuvering their tongue. The adventitious movements were more frequent and had ""milkmaid"" movements of the hands when holding onto the examiner's fingers. The patient was admitted to the neurology service for expedited work-up and treatment where they had minor improvement, but diagnostic evaluation only yielded results of unclear significance.

The patient continued to show functional decline. The patient began to use assistive devices for communication because their speech was so severely dysarthric that it was difficult to understand single words. The patient had persistent dysphagia and required near-continuous home care to assist with their activities of daily living. At best, the patient was able to transfer out of their wheelchair. On exam (three years after the prior exam), the patient's speech had developed a nasal quality with slowing and impairment of lingual, palatal and buccal consonants. The patient's tongue had near constant movements without fasciculations. There was some decreased bulk in the intrinsic hand muscles bilaterally. The patient also had mild truncal titubation at rest with intermittent lower extremity movements.

The patient suffered a C7 fracture at home after slipping off of their sofa. The patient was admitted but their hospital course was complicated by pneumonia followed by a large pulmonary saddle embolus in the setting of bilateral DVTs, requiring anticoagulation. Following worsening respiratory function, the patient was transitioned to DNR/DNI. The patient died two days later (at age XX, eight years after their initial presentation) and an autopsy was performed.

**Example 12:**

Neuropathological definitive diagnosis: Glioblastoma Multiforme

DIAGNO/Neurologist diagnoses:

|  | Top 1 Dx | Top 2 Dx | Top 3 Dx |
| --- | --- | --- | --- |
| DIAGNO | Glioblastoma Multiforme (GBM) | Primary Central Nervous System (CNS) Lymphoma | Metastatic Brain Tumor |
| Neurologist | Primary CNS vasculitis | CADASIL | Cerebral venous sinus thrombosis |

Case summary: A XX-year-old right-handed patient presented to an emergency department with a headache lasting several hours. This was felt to be an exacerbation of their ocular migraine headaches (which were consistently 20 minute episodes of a zig-zag pattern in their left visual field with flashing lights without pain). Their exam was reportedly “reassuring”, so they received symptomatic treatment and were sent home. Over the next several months they had intermittent headaches (reported as 4-5/10, right frontal/retro-orbital) without any visual phenomena, or oto- or photophobia. About 2 months after the ED visit, they noticed poor balance, falling to the left, and bumping to object on their left side. They had intermittent right incisor tooth pain and a stiff neck over the same time period. They also reported episodes of mild motor complaints: at times having difficulty hitting the correct key while typing and momentary difficulty in coordinating a movement with their left foot. Three months after their first ED visit, they returned to the outside ED because of continued and worsening headache. Imaging studies were performed and they were transferred to MGH.

Their past medical history included ocular migraine, 20 pack-year smoking (quit at the age of XX), heavy drinking until they were XX, and depression. Their medications included an antidepressant and prn anxiolytic. Their most recent mammography was 3 years earlier and colonoscopy was 15 years earlier; both were unremarkable. Their family history is significant for migraines, GI cancer (grandmother) and prostate cancer (grandfather).

At MGH, their vital signs and general exam were unremarkable. On neurological exam, they were oriented to place and time; they had intact registration and recall but had difficulty listing the months backwards. Their pupils were equal and reactive. Visual field testing revealed left homonymous hemianopia. Extraocular movements and facial sensation were intact. They had a mild left lower facial asymmetry but their tongue was midline and the palate elevated symmetrically. They had mild weakness of their left sternocleidomastoid, mild left upward drift, and mild left deltoid, biceps, and triceps weakness. They also had mild weakness of left hip flexion. They had an up-going toe on the left but their reflexes were normal and symmetric otherwise. They had mild dysmetria on finger-nose testing in the left upper extremity. Their gait was unsteady. Further diagnostic testing was performed and treatment was started. They were discharged after an 11 day hospitalization; at that point, they still had the field cut, but their headaches were milder, and their weakness improved.

They were seen one week later in clinic for follow up and their clinical condition was stable. However, they presented to the emergency department a week after their clinic visit with left lower extremity weakness and odd behavior. On exam, they had difficulty with calculation and were unable to list the months of the year backwards. They also had a left visual field cut, mild left facial droop, mild extensors weakness in left upper extremity, and mild left hip flexion weakness. Imaging studies showed new findings, prompting a different therapeutic regimen. They were discharged to a rehab facility. Over the next two months, their deficits resolved moderately including both motorically and cognitively.

A month later, they had a ten day course of worsening left sided weakness involving both arm and leg, along with worsening balance. With multiple alterations in their medical management were made, there was some improvement. They also reported problems with their short term memory and attention to tasks. There was also increasing lower extremity edema.

A month later, they were re-admitted with confusion and failure to thrive. They were found to be disheveled, confused, with worsening left sided weakness. Their ambulation was limited to few steps. While an inpatient, they were noted to become acutely short of breath. Diagnostic testing was performed and new medications were started. Two days later, they had acute onset distal left lower extremity plegia. Their therapeutic plan was changed and a therapeutic procedure was performed. Although their condition remained stable for several days, they were discharged to hospice. They died 3 weeks later (7 months after their presentation to MGH) and an autopsy was performed.

**Example 13:**

Neuropathological definitive diagnosis: Neurofibromatosis type 2

DIAGNO/Neurologist diagnoses:

|  | Top 1 Dx | Top 2 Dx | Top 3 Dx |
| --- | --- | --- | --- |
| DIAGNO | Neurofibromatosis Type 2 (NF2) | Chiari Malformation Type 1 | Intracranial Hypertension (Secondary to Chiari Malformation) |
| Neurologist | Idiopathic intracranial hypertension, refractory to medical treatment | Ventriculo-peritoneal shunt complicated with bilateral subdural hematomas due to overshunting | Cerebral venous sinus thrombosis |

Case summary: A XX-year-old individual presented to an outside clinic because they noticed that their left biceps was smaller than their right biceps. Bilateral papilledema was noted on exam. Workup was unremarkable except for a Chiari type 1 malformation was noted on MRI, as well as acute on chronic denervation of the left biceps and deltoid noted on EMG/NCS. A diagnosis was not established, and the patient was followed clinically.

XX years later, the patient started college with a major in music and transferred their care to MGH. During their first clinic visit, they denied any headaches, visual problems, facial weakness, swallowing difficulty, weakness (except in their left arm), numbness, hearing problems, tinnitus, bowel or bladder dysfunction. Their exam was notable for bilateral optic disc elevation and 4/5 strength at the left biceps (27 cm in the mid-biceps region as compared to 30 cm the right). A test was performed, which established the diagnosis. However, no causative lesion was identified for their arm atrophy.

Following the diagnosis, the patient initially did very well except for occasional headaches, for which they were started on a medication with good response. They graduated from college XX years later and went on to become a flute player and maker. The patient was followed clinically and with regular imaging -- which prompted an intervention XX years later. Following the intervention, they had transient right leg weakness, which later resolved completely.

The patient remained clinically stable until XX years later when they started to notice a slight decline in their hearing. Formal hearing test revealed deficits in their pure-tone averages rather than their word recognition scores (WRS). They also developed occasional episodes of right leg ascending numbness, for which another medication was started. The following year, the patient experienced a sudden decrease in their hearing in the setting of a viral illness. The decrease in hearing had persisted since and they had difficulty following conversations when there were background noises. Formal hearing test showed a measurable decline in their WRS. The patient also noticed a mass under their left chin. They experienced discomfort and changes in their hearing when they pressed the mass.

XX years later, the patient developed recurrent episodes of headaches, visual obscurations and imbalance. A test was performed, which confirmed the diagnosis and prompted an intervention. Their symptoms improved following the intervention. However, their hearing continued to worsen and the WRS declined to 38% (from 82%) over the course of a year. Surveillance studies showed progression of their disease and an experimental drug was trialed. The patient developed severe nausea with the drug and had to discontinue it several months later.

XX years later, their headaches recurred, for which they underwent an intervention. Following the intervention, their headache improved, however, they developed difficulties projecting their voice and swallowing liquids.

XX years later, the patient reported new sensation of numbness and tingling in their legs associated with difficulty walking, which resolved after another intervention.

XX years later, the patient developed severe headaches which they described as “feeling I’m going to die”. The headaches were positional (worse lying down and better standing up) and associated with darkening of their central vision and tinnitus. The headaches were refractory to maximal medical management and they had to quit their job as a flute maker. On exam, they had normal mental status and language. Their pupils were 6 mm OD/5 mm OS and reactive. Facial sensation and strength were normal. Tongue deviated to the left with atrophy. There was no pronator drift and their strengths were full except for 4/5 at the left biceps with atrophy. Sensation, coordination and gait were all normal. The patient then underwent an intervention for their severe headaches. Immediately following the intervention, they were initially awake and able to follow commands. However, several hours later, they were noted to have decreased mental status, which rapidly progressed to somnolence with a new anisocoria, prompting multiple emergent studies and interventions.

Their exam continued to be poor in the following days. A family meeting was held, and the decision was made to focus on comfort. The patient died 2 days later at age XX (XX years after the initial presentation) and an autopsy was performed.

**Example 14:**

Neuropathological definitive diagnosis: Cerebrovascular disease

DIAGNO/Neurologist diagnoses:

|  | Top 1 Dx | Top 2 Dx | Top 3 Dx |
| --- | --- | --- | --- |
| DIAGNO | Alzheimer's Disease | Vascular Dementia | Lewy Body Dementia |
| Neurologist | Alzheimer's disease | Mixed Alzheimer's and cerebrovascular small vessel ischemic diseases | Mixed Alzheimer's, Lewy body, and cerebrovascular small vessel ischemic diseases |

Case summary: A XX-year-old individual was referred to the neurology clinic for cognitive changes. The patient described issues with short term memory noting that they frequently repeat themselves, couldn’t recall day to day events, and had word finding difficulties. The issues had progressed over the prior 3 years, causing increasing difficulties paying bills and taking their complex regimen of medications. The patient had lost interest in social interactions. They continued to read, watch favorite television shows, and do crossword puzzles. The patient was now living in an independent unit of a retirement community. On review of systems, the patient also noted visual loss of unclear duration, hyposmia for many years, urinary incontinence, and peripheral neuropathy.

Past medical history was significant for obesity, OSA, CHF, long-standing poorly controlled DM, HLD, HTN, and pulmonary embolism in the setting of DVT XX years before presentation (on warfarin). The patient had congenital polydactyly treated with multiple surgeries in childhood. The patient was a retired professor and had also worked as a therapist. The patient had no smoking history or other toxin exposure. Family history included an aunt with Parkinson disease, a cousin with Alzheimer disease, and a great aunt with dementia not otherwise specified, in addition to breast cancer affecting the patient's mother and daughters.

On examination, the patient was oriented to person, place, month and year, but not date. The patient completed the modified trails B task accurately but had difficulty drawing a cube. Language (simple naming, repetition, and comprehension) was intact. The patient's speech had normal rate, rhythm and prosody. The patient generated 18 words beginning with the letter L in 60 seconds. The patient took four trials to register 5 words, and after a delay, recalled 2/5 spontaneously and the remaining 3 with either a category or multiple choice cue. Digit span was intact. The patient made one error on serial 7’s. Abstraction was normal. The patient's mood appeared mildly depressed and they showed signs of psychomotor slowing. The patient's thought processes were logical, judgment was intact, and they showed no evidence of psychosis. On the remaining neurological examination, the patient had visual field defects in inferotemporal quadrants bilaterally, remaining cranial nerve III-XII intact. Motor tone, strength and bulk were normal throughout. There was no tremor or cogwheel rigidity. Sensation was intact to light touch. Vibration was diminished in the distal lower extremities. Deep tendon reflexes were zero to trace in all extremities. Cerebellar testing showed normal finger-nose maneuvers. Gait was wide based and slowed. There was no retropulsion instability. A range of studies were performed and follow up was arranged.

A few months later, the patient had acute onset of left leg weakness, gait instability (“felt tilted”) and had difficulty seeing out of their left eye, which apparently resolved after a few minutes. The patient was evaluated at an outside hospital where their neurological exam was unchanged from baseline and other studies were unrevealing.

Almost one year later, the patient again presented to a local hospital with headache, slurred speech and aphasia (not described further). The patient's blood pressure was elevated to systolics of 200s and their blood glucose had been 380 the previous day. The patient's symptoms resolved over about 3 hours along with optimization of their blood pressure; by report multiple imaging studies were unremarkable. In the following 6 months, the patient had 2 similar episodes of transient confusion, aphasia, and slurred speech – each time with admission to an outside hospital. No additional information from these episodes is available.

The patient's cognitive status declined over the following years, they more frequently missed medications, had more difficulty reading, and had confusional episodes. Formal memory testing was not performed, but the report from the patient's daughter was that their deficits were inconsistent. At the patient's final visit with their PCP, they are described as “cheerful, interacting with short sentences”. The patient died at age XX, approximately 3 years after their first assessment, and an autopsy was performed.

**Example 15:**

Neuropathological definitive diagnosis: Multifocal demyelinating lesions in the periventricular white matter and spinal cord, consistent with history of multiple sclerosis

DIAGNO/Neurologist diagnoses:

|  | Top 1 Dx | Top 2 Dx | Top 3 Dx |
| --- | --- | --- | --- |
| DIAGNO | Multiple Sclerosis (MS) | Chronic Inflammatory Demyelinating Polyneuropathy (CIDP) | Primary Progressive Multiple Sclerosis (PPMS) |
| Neurologist | Spinocerebellar ataxia | Adrenoleukodystrophy | Multisystem atrophy |

Case summary: A XX-year-old individual developed intermittent headache and double vision. The patient presented to an outside hospital, where they performed a battery of tests including CNS imaging and a lumbar puncture (results not available). The patient's symptoms resolved, and although a possible diagnosis was made there was no treatment.

The patient did not present to the hospital again until XX years later when they had an isolated “grand mal” seizure. The patient worked as a carpenter, and the seizure was thought to be related to “inhalation of vapors” at work. The patient was started on an anti-seizure medicine, though this medicine was eventually discontinued without further seizures.

One year later, at age XX, the patient was hospitalized at an outside facility after developing progressive difficulty with walking and balance. A careful review of the patient's history revealed brief transient episodes of neurologic impairment in the past including left facial droop (diagnosed as Bell’s palsy), left leg fatigue, and a band-like sensation around the abdomen. The patient had additional workup (only limited information available) and a diagnosis was established. The patient was started on treatment, and while their walking and balance improved, they were left with mild residual deficits.

The patient remained stable until age XX when they had another episode of difficulty walking. The patient was treated acutely, their long-term regimen was changed, and they were referred to MGH Neurology for further care. When seen in clinic, a full neurologic examination documented the patient's new baseline. The patient's cranial nerve exam was notable for visual acuity with correction 20/20, normal funduscopic examination, PERRL, mild horizontal nystagmus, mild left facial weakness (LMN pattern) with intact sensation, mild dysarthria, and midline plate/tongue. The patient's motor exam revealed no deficits. In the patient's lower extremities, they had decreased vibratory sensation L>R. The patient had bilateral lower leg ataxia L>R. The patient's knee and ankle jerks were 3+ bilaterally with upgoing toes. The patient ambulated with a mildly spastic ataxic gait and was unable to tandem.

Less than a year later, the patient developed worsening ataxia, a persistent moderate tremor on the left with action and rest components. The patient's therapeutic regimen was adjusted.

Over the next several years, the patient was overall fairly stable on the new treatment regimen, but they developed several new symptoms that proved difficult to treat including urinary urgency/frequency and fatigue. The patient developed osteopenia of the left hip and suffered from frequent urinary tract infections. The patient's neurologic exam was stable except for new subtle right internuclear ophthalmoplegia and mild weakness in left foot dorsiflexion. The patient required a cane intermittently for ambulation. The patient's therapeutic regimen was adjusted again.

One year later, at the age of XX, the patient developed progressive and relentless worsening of strength on the left side. Various new approaches to treatment were attempted but without significant overall benefit. By the age of XX, the patient was wheelchair bound due to complete plegia on the left side. Several medicines had been started to treat symptoms of the patient's disease including baclofen for spasticity and botox injections for incontinence. The patient required frequent catheterization, and they had several hospitalizations for sepsis as a complication of urinary tract infections.

Five years later, at the age of XX, almost thirty years after the patient's first symptoms, the patient suddenly developed difficulty breathing. The patient otherwise appeared to be in their usual state of health. EMS was activated, but respirations had stopped by the time they arrived. The patient was pronounced dead and an autopsy was performed.

Supplementary Note 6. Fifteen Examples of Cases where DIAGNO was Incorrect and Neurologist was Correct.

**Example 1:**

Neuropathological definitive diagnosis: Cerebrovascular disease: small vessel vasculopathy with arteriolar sclerosis, severe, diffuse

DIAGNO/Neurologist Dx:

|  | Top 1 Dx | Top 2 Dx | Top 3 Dx |
| --- | --- | --- | --- |
| DIAGNO | Multiple Sclerosis (MS) | Neurosarcoidosis | Recurrent Ischemic Stroke |
| Neurologist | Vascular dementia due to severe small vessel ischemic disease | Multi-infarct vascular dementia | Neurosarcoidosis |

Case summary: A XX-year-old right handed individual presented to an ED because their left side did not “feel right”. The day prior the patient felt somewhat “off” and was dizzy in the afternoon when finishing work. The patient thought that was from their inner ear and went to bed. Overnight the patient noticed that they were unstable when they got up to go, tilting to the left but not falling. In the morning, this unsteadiness was worse and the patient fell as they tried to get to the bathroom. The patient's partner also noticed thickening of their speech. The partner got the patient in the car and took them to ED. The partner noted a facial droop and that the patient was unable to bear weight on their left leg. At the ED, the patient denied sensory deficits or paresthesias in their extremities but did feel that the left side of their face was numb (“like novocaine”). The patient felt they had trouble bringing words out, but denied trouble thinking or word-finding. The patient had felt “electrical shots” down their left leg over the past day. The patient had experienced similar episodes of dysequilibrium in the past, which they attributed to inner ear disturbances, but never before accompanied by weakness. The patient had no pain, had made no abrupt maneuvers or movements prior to onset, and there had been no recent trauma.

The patient's past medical history included hyperlipidemia and biopsy-proven pulmonary sarcoidosis, diagnosed XX years earlier via mediastinoscopy for hilar adenopathy discovered in the work-up for a painful peripheral neuropathy. The patient had been treated with steroids for several months before being tapered off and was on no medications at presentation. The patient's mother had diabetes and a paternal aunt had multiple sclerosis.

On admission, the patient had decreased sensation to light touch on the lower left face, left facial weakness with slow activation of their forehead, a flaccid left arm, 4/5 weakness of the proximal left leg muscles but 0-2/5 strength distally. The patient's left toes went up. Imaging and other studies including an LP (clear CSF, 1 WBC, 50 RBC, glucose 65, protein 35) were performed, following which management was begun.

The patient was discharged to a rehab facility where they made extremely rapid progression in returning to normal, such that when seen a month later their facial droop remained but sensation was normal. The patient had “fairly good” strength in the left leg proximally (not further quantified), although distally there was foot-drop. With intense effort, the patient could elevate their left arm approximately 60 degrees and extend it from the body 8 to 12 inches. The patient had no movements with their hand. There was increased tone in the left arm greater than leg with spread of reflexes and a profound Hoffman's sign. Subsequently, the patient still had the facial droop but now had full strength in deltoid, biceps and triceps muscles on the left, with 4/5 weakness in the left finger extensors and 4-/5 weakness in left finger intrinsic muscles, and 5-/5 weakness in left grip strength. The patient's tone remained elevated on the left and those toes continued to go up. The patient's treatments continued.

XX years later (age XX) the patient was assessed again. The patient had had no neurologic events since the prior hospitalization although their management continued. The patient's face now moved symmetrically. The patient's tone was still increased on the left which influenced their gait (described as “spastic”), while their strength was 4/5 in their left deltoid, triceps, finger extensor, hamstring. The patient's left toe remained upgoing, with 2+ reflexes on the left (1+ on the right).

A few months later the patient had two episodes of language deficit. The first was sudden onset of difficulty typing correct words. The patient tried to go home from work but was unable to find their keys. In the ED the patient didn't know their partner's name but knew they were their partner. The patient couldn't say how many children they had. The patient was frustrated and was aware that they were not at their baseline. The patient's partner and the nurse noted a subtle right facial droop. There was no slurring of the speech. The patient was admitted overnight, studies were performed and management initiated. A month later the patient had a second similar episode when driving. The patient used incorrect words and was unable to read. The patient had no weakness, was able to pull the car over and complete procedural tasks. Each episode lasted ~1 hour, and the patient had memory of both episodes.

When seen several months later, the patient's partner felt that the patient's thinking processes, multitasking problem-solving executive functions were not quite normal. The patient was vaguely aware of these changes, having needed to shift out of a demanding administrative job. Several months later the patient was admitted with new loss of dexterity in their right hand and severe left-sided weakness and gait instability that closely resembled the deficits from the earliest episode. On exam there was marked spasticity, 4/5 flexor and 3/5 extensor strength in left arm with 2/5 digital abductor weakness of the left hand, while the left leg showed spasticity, 3/5 flexor and 4/5 extensor strength. There was normal tone and 5/5 strength in both upper and lower right extremities. There was no bradykinesia, pronator drift, fasciculations, myoclonus or tremor on either side. The notes don’t clearly describe deficits in the right hand. The patient was discharged after a few days.

At age XX, the patient was again admitted after experiencing sudden fatigue while at home following which they were found by their family to be confused, responding to all questions with ""no"", disoriented, unable to name their family members. The patient had a severe headache, and had nausea, dry heaving and small amount emesis. The patient's mental status continued to worsen, stopping following commands and intermittently being difficult to arouse. The patient was admitted to an outside hospital where they cleared somewhat over a few days, before being transferred to MGH. At that time, the patient's strength was 4+/5 in the left arm, normal elsewhere; the patient maintained their increased tone and upgoing toe on the left. Various alterations in the patient's medical management were begun and the patient continued to have their mental status head towards their baseline. The patient continued to have episodes admissions for decline in mental status, often accompanied by exacerbations of their weakness.

Eventually, the patient moved into an assisted living facility because of inappropriate behaviors, difficulties with memory, and attention. While there, the patient suffered a fall with a head strike; subsequent imaging showed new findings. The final available neurologic exam (age XX) showed mild left nasolabial fold flattening, increased tone and spasticity in the left arm and leg with the patient's left wrist held in flexion (the patient was unable to fully extend their wrist or fingers). On the left, the patient's strength was 4/5 at the deltoid, triceps, wrist extension, and greater weakness on finger extension and abduction. The patient had mild left hip flexion weakness, 4/5 knee flexion, and an inability to dorsiflex at the left ankle. The right arm and leg were full strength. Left sided reflexes were brisk with crossed adductors and pectoral spread. The patient was unable to stand. The patient died a year later, age XX, and an autopsy was performed.

**Example 2:**

Neuropathological definitive diagnosis: Brain with multiple embolic infarcts of varying ages from 24 hours to 4-5 days

DIAGNO/Neurologist Dx:

|  | Top 1 Dx | Top 2 Dx | Top 3 Dx |
| --- | --- | --- | --- |
| DIAGNO | Aortic Dissection-induced Spinal Cord Ischemia | Hypoxic-Ischemic Encephalopathy | Cocaine-Induced Vascular Damage |
| Neurologist | Acute ischemic stroke | Intraparenchymal hemorrhage | Anoxic brain injury |

Case summary: A XX-year-old individual was woken from sleep by severe chest pain that radiated to their back. Their partner immediately called EMS and the patient was transported to MGH ED. The patient also had bilateral leg numbness with parasthesias.

The patient's past medical history includes poorly controlled hypertension (secondary to medication non-compliance), active cocaine use (last used XX week prior to admission) and tobacco use, obstructive sleep apnea (non-compliant with CPAP) and heart failure with preserved EF. The patient did not take any medications and had no known allergies. The patient has a family history of cardiac disease. The patient's sister had an MI at age XX and their father died of unknown cardiac disease.

On arrival, the patient's vitals were T 98 HR 66 BP 216/102 RR 24 99% on 4L. Imaging demonstrated a large aortic dissection that started after the takeoff of the left subclavian artery, spanned the entire abdominal aorta and terminated in the right external iliac artery. The patient was started on medications for blood pressure and pain control. On exam, the patient was diaphoretic and in pain. Cardiac exam did not suggest any evidence of murmurs. The patient's neurologic exam was documented as evidence of bilateral lower extremity numbness with mild weakness. The patient was admitted to the SICU for further care.

During the first 3 days of the patient's admission, there were some difficulties with blood pressure control and fluid status, but the patient overall remained clinically stable. Though the patient's movements were limited by pain, there were no documented new or worsening neurologic symptoms. However, over the next 2 days, the patient had intermittent chest pain episodes which became more frequent. Reimaging, which had previously suggested a stable aortic lesion, now showed a 5 mm increase in the size of the patient's thoracic aorta, so a two-staged operation was performed. After the patient's second surgery, the patient was noted to be agitated but moving all extremities to pain. The patient was given an antipsychotic for agitation. Several hours later (in a poorly defined three-hour window), the patient was noted to be agitated and started posturing. An imaging study was performed and neurology and neurosurgery were consulted.

Neurologic examination (performed 15 minutes off propofol) found the patient to have their eyes closed, without response to voice or painful stimuli. There was no evidence of spontaneous tearing. The patient's pupils were 2.5 mm on the left and 2 mm on the right; both were reactive. There was “highly questionable” blink to threat on the right and none on the left. The patient's primary gaze was dysconjugate with right esophoria. With oculocephalic stimulation, there was right eye abduction and downward deviation but minimal adduction of the left eye; no vertical eye movements were elicited by the procedure. Corneal reflex was significantly stronger on the left than the right. Both gag and cough reflexes were intact. The patient was not over-breathing the ventilator. The patient's motor tone was decreased throughout. In response to stimulation, the patient had extensor posturing of the right arm and “incipient” extensor posturing of the left; both legs showed triple flexion although more on the left than the right. The patient's plantar reflex was mute on the left and upgoing on the right. Management was initiated.

The following day, the patient's pupils were both 5 mm and the right was non-reactive. Oculocephalic movements elicited minimal left eye abduction and no movements of the right eye. Corneal reflexes were absent as was response to nasal tickle. The patient's gag was weak and there was no cough with suctioning. The patient's motor tone was increased and had extensor posturing of both arms. In the setting of the patient's progressive neurologic decline over several days, a family meeting was held and the patient was transitioned to intensive comfort measures. The patient died the next day (2 weeks after admission) and an autopsy was performed.

**Example 3:**

Neuropathological definitive diagnosis: Diffuse Large B Cell Lymphoma, EBER positive, pons and medulla

DIAGNO/Neurologist Dx:

|  | Top 1 Dx | Top 2 Dx | Top 3 Dx |
| --- | --- | --- | --- |
| DIAGNO | Brainstem Stroke | Central Pontine Myelinolysis (CPM) | Progressive Supranuclear Palsy (PSP) |
| Neurologist | Lupus cerebritis | Acute demyelinating encephalomyelitis | CNS lymphoma |

Case summary: A XX-year-old patient began experiencing episodes of dizziness, weakness and difficulty eating and drinking. These complaints prompted the patient to seek care. The patient was admitted to an OSH where a series of studies were performed and a treatment was initiated. The patient was then discharged to a rehab facility.

The patient's past medical history was significant for a stroke at the age of XX, prompting a diagnosis of anti-phospholipid antibody syndrome at that time. The patient was started on anti-coagulation, which was continued for the next five years, but then stopped, continuing to take a daily baby aspirin. Approximately XX years later the patient was diagnosed with lupus, for which the patient was maintained on chronic immunosuppressive therapy.

While at the rehab facility, the patient developed dysphagia and increased difficulty swallowing. A few weeks after the initial evaluation, the patient was admitted to an OSH because of increasing somnolence. The patient required intubation for airway protection and was transferred to MGH.

On presentation to MGH the patient's exam was significant for dysconjugate gaze with ocular bobbing bilaterally. Pupils were anisocoric with right – 5 mm and left – 3 mm. Right pupil was not reactive to light. The patient demonstrated limited ability to follow commands. The patient had some intact extraocular movements, better in the vertical than horizontal plane. Motor exam was antigravity throughout without drift, more brisk to activate on right than left, but the patient was not fully cooperative with the exam. Sensation to noxious stimuli was present throughout.

Imaging was obtained and an LP was performed (RBC 22->1, WBC 32->40 [96-98% lymphocytes], glucose 77, protein 93). A range of additional studies were performed. The patient's exam did not improve and a possible therapeutic approach was discussed at a family meeting. It was decided to proceed with the attempt at treatment; a tracheostomy tube was placed in order to facilitate this. At this time the patient's exam showed the patient to be alert, regarding and tracking and able to follow basic midline and peripheral commands. The patient seemed to understand their circumstance. Vertical gaze was intact but the patient could not move their eyes horizontally. The patient also had difficulty closing their eyes. Facial sensation was intact to light touch. The patient's strength was at least 4/5 throughout with L > R weakness and the patient moved all extremities spontaneously. Light touch perception was intact.

Despite this therapy the patient continued to decline over the following weeks and the patient's imaging showed continued and increasing abnormalities. Approximately XX days after the treatment, the patient's exam showed the patient to be able to answer Yes/No questions and the patient continued to demonstrate understanding of their medical issues. The bobbing eye movements persisted but the patient now had no voluntary eye movements. The patient's face appeared to be diplegia.

About XX days later, the patient's exam included ability to answer Yes/No questions, facial diplegia, vertical bobbing but no other eye movements, only trace movements in the patient's hands and feet. The decision was made to remove ventilatory support and the patient died shortly thereafter, XX months after the patient's first presentation, and an autopsy was performed.

**Example 4:**

Neuropathological definitive diagnosis: Metastatic Melanoma

DIAGNO/Neurologist Dx:

|  | Top 1 Dx | Top 2 Dx | Top 3 Dx |
| --- | --- | --- | --- |
| DIAGNO | Cerebellar Ataxia | Paraneoplastic Syndrome | Hypertensive Encephalopathy |
| Neurologist | Metastatic cancer with brain metastasis | Epilepsy | Cardiogenic syncope |

Case summary: A XX-year-old individual was waiting outside their home for a taxi when they had sudden unwitnessed loss of consciousness. The patient recalled walking toward the curb and then just “went out.” They woke up on the ground; their sunglasses were broken nearby. They estimated that they were only down for a few minutes because the taxi was just pulling up to the curb. When they woke up, they felt well enough to get into the taxi and proceeded as planned to their bank. When they arrived, a bank teller told them that they had blood on their face. They went home and contacted their doctor who recommended that they present to clinic the next day.

The patient's past medical history included hypertension, hyperlipidemia, prostate cancer, a skin lesion s/p Mohs procedure. They were a retired postal worker. Two months prior to this episode, they had discovered a mobile, soft, subcutaneous, non-tender nodule on their right shoulder while showering. The patient felt well otherwise and denied any new neurological changes including: weakness, sensory changes, language difficulties, visual changes, and coordination and gait problems. A range of imaging studies were performed, followed by establishment of a diagnosis. One month later, they had repeat radiologic evaluation and then were started on a new treatment.

At clinic, the patient was referred to MGH ED and neurology was consulted. When examined, there was a large bruise evident over their right eye. They were fully oriented, able to name days of the week backwards and demonstrated no abnormalities of language. Their pupils were equal and reactive, while their extraocular movements were normal. Visual fields were full to finger testing, their face was symmetric and their tongue midline. Motor testing was normal, with no pronator drift, no orbiting, and there was full strength throughout. Their reflexes with 2+ throughout and both toes went down. Finger-to-nose testing was unremarkable. The patient was admitted to an inpatient service for further work-up and evaluation. They were eventually discharged with an increase to their beta-blocker, initiation of an anticonvulsant medication 500 mg BID and continuous cardiac monitoring.

The patient continued with their prior treatment regimen following the loss of consciousness episode. Additional assessment and diagnostic testing was performed approximately 2 months later, resulting in the decision to shift their therapeutic regimen.

Over the next months, the patient developed new symptoms of nausea/emesis, “vertigo”, and they described their gait similar to a “drunken sailor.” They began using a cane and occasionally a wheelchair. At their clinic visit, they were alert, oriented, language fluent, comprehension and naming intact. Their gait was unstable, and they had poor finger-to-nose performance; strength was intact. Imaging studies were performed which prompted an intervention. Following this, they were assessed by PT who documented a gait speed of 1.19 m/s (better with cane than without). Shortly after discharge, they were started on another therapy.

Two months later, the patient presented to MGH with bilateral lower extremity weakness, failure to thrive, dehydration, and a fall at home. Limited information is available in the chart about their neurologic function, with the exam reported to show intact cranial nerve function, some weakness in forearm flexion with just the ability to move their legs against gravity. Lactic acid trended up to 4.1 in the initial 24 hours and then normalized. They also developed an episode of SVT with HR in the 200s and SBP to the 80s. After medical stabilization, they were discharged to home. The patient died approximately 1 month later, and an autopsy was performed.

**Example 5:**

Neuropathological definitive diagnosis: Diffuse Large B Cell Lymphoma, non-germinal center type

DIAGNO/Neurologist Dx:

|  | Top 1 Dx | Top 2 Dx | Top 3 Dx |
| --- | --- | --- | --- |
| DIAGNO | Alzheimer's Disease | Vascular Dementia | Lewy Body Dementia |
| Neurologist | CNS malignancy (ex CNS lymphoma) | CJD (Creutzfeldt-Jakob disease) | Autoimmune encephalitis |

Case summary: A XX-year-old individual began to develop memory difficulties. The patient's daughter felt that the patient was also more irritable, and was increasingly fatigued and had a decreased appetite, having lost 12 lbs. over the last year. The patient had trouble remembering conversations and appointments, with an instance where the patient showed up to the daughter’s house and took the granddaughter to school without notifying the daughter. When asked questions, the patient would repeat the question back and the family would often have to play charades to guess what the patient was trying to say. The patient's past medical history was notable for osteoporosis and the patient was status post right hip and knee replacements.

After about five months of worsening symptoms, the patient was evaluated at an outside hospital, and imaging studies were performed. The patient was diagnosed with a TIA and discharged on a heart monitor. The patient was then taken to the PCP, who started a medication and referred the patient to a neurologist, who the patient saw 5 months later.

By the time the patient saw the neurologist, the patient had become increasingly weak, dragging the left leg behind when walking until the patient finally had to use a wheelchair. The patient would complain of odd smells like burning oil. The family reported bowel and bladder incontinence. Information from that assessment is not available.

Five days after seeing the neurologist, the patient had an episode of incontinence where the patient voided on the floor of the bathroom, walked out to the living room and fell down the stairs. The patient was found down, but conscious and interactive, by the family. The patient was taken to the ED at MGH where the patient was drowsy and oriented only to person. The patient's speech was fluent, but the patient had minimal spontaneous output. The patient could follow simple but not complex commands, with impaired attention and memory. Pupils were equal and reactive to light. There was no ptosis. Extraocular movements were intact without nystagmus. There was left nasolabial fold flattening. The palate elevated symmetrically and the tongue was midline with protrusion. Strength was 4/5 in both arms and at least 3/5 in the legs, although the patient would not lift the left leg antigravity until asked to while the sock was being put on the foot. DTRs were 3+ on bilateral biceps, brachioradialis, left quadriceps, 2+ on the right quadriceps. Diagnostic studies were performed and medications were begun.

The next day, the patient was less responsive with no verbal output. The patient would withdraw to painful stimuli and spontaneously move the right side. The patient underwent a diagnostic procedure and was started on a new medication. Over the next few days, the patient's mental status improved partially, but continued to wax and wane.

Five days later, the patient's mental status declined. The patient wouldn’t open the eyes to command and groaned to sternal rub. The patient localized to pain on the right and moved the right upper extremity and lower antigravity spontaneously. The patient was unable to follow other commands. The left eye was slightly down and out. The patient withdrew slightly with painful stimuli on the left lower leg. The left upper arm showed increased tone. Due to the patient's increased somnolence, a LP was performed. The opening pressure was 9 cm of water, glucose 78, protein 352, RBC 1, Nucleated cells 9 (96L/3M/1Neut). The patient continued to decline and three days later, was transitioned to comfort care.

Over the next few days, the patient's mental status improved and the patient would answer yes/no questions, follow commands, and could eat small amounts of pureed foods. Seven days after the patient was transitioned to comfort care the patient's mental status declined again, where the patient would not respond to voice or painful stimulation. Two days later, 21 days into the current hospital presentation and approximately 10 months since the initial symptoms, the patient died and an autopsy was performed.

**Example 6:**

Neuropathological definitive diagnosis: Anaplastic astrocytoma, IDH-wildtype, WHO grade III of IV

DIAGNO/Neurologist Dx:

|  | Top 1 Dx | Top 2 Dx | Top 3 Dx |
| --- | --- | --- | --- |
| DIAGNO | Progressive Multifocal Leukoencephalopathy (PML) | Amyotrophic Lateral Sclerosis (ALS) | Primary Central Nervous System Lymphoma (PCNSL) |
| Neurologist | High grade glioma | Brain metastasis | Brain abscess |

Case summary: A XX-year-old individual had an episode of blurry vision when looking to the right. The episode lasted for an hour and was followed by a mild left-sided headache. The following day, while at work, the patient noted tingling in their right hand that spread to involve the entire arm. This lasted about 4 minutes and resolved spontaneously. The patient had no change in mentation noticeable to their co-workers. EMS was called and the patient was brought to an outside hospital. Imaging was performed, a medication was started and the patient was sent home. The following day, the patient had two additional episodes (<4 minutes long), of right hand, arm and face paresthesias. These episodes prompted the patient to present to the MGH ED.

The patient's past medical history included gout, hypertension, diverticulitis and hypercholesterolemia. The patient took an antihypertensive medication, another antihypertensive medication, aspirin, a diuretic, and a cholesterol-lowering medication as their baseline medications. The patient was married with two children (both healthy), worked as an accountant, had never smoked and drank socially. There was a family history of malignancy with prostate cancer (father) and lung cancer (sister). The patient's mother had diabetes and stroke.

When the patient was seen at MGH, their symptoms had resolved and they complained only of a mild cough. Their temperature was 98.5 C, HR 68, blood pressure 162/76, respiratory rate 16 and oxygen saturation was 97% on room air. Neurologic exam in the ED was normal, including light touch, strength testing and gait. Basic labs were unremarkable aside from glucose of 124. An additional diagnostic image was obtained. The patient was advised to continue the medication they were recently started on at the outside hospital at an increased dose. The patient was discharged to outpatient follow-up.

Exam remained unremarkable and imaging remained stable until XX months after the initial presentation when imaging abnormalities prompted additional studies. The patient was started on additional therapy. Over the subsequent year, the patient developed difficulty with short-term memory and problems with word finding. The patient scored 28/30 on a MMSE, losing two points for short-term recall. The patient developed fatigue, difficulty concentrating, and dizziness that gradually worsened over a period of months.

Subsequently, the patient required the use of a cane and had difficulty with tandem walking. Additional therapy was started. The patient began to fall at home. Additional diagnostic imaging was performed and prompted a change in therapy. The patient subsequently developed extreme fatigue, episodic difficulty speaking, diffuse bilateral weakness in their extremities, lower worse than upper, and occasional urinary incontinence.

The patient presented to an outside hospital due to inability to get out of the shower. The patient was treated for groin cellulitis and their most recently instituted therapy was held. Weakness and fatigue were so severe now that the patient required a Hoyer lift to move from chair to bed and could not move their legs. Family noted occasional bilateral tremors and at least one episode of bilateral flexor posturing during which the patient was not responsive. The patient required assistance to eat. The last documented neurologic exam showed the patient to be oriented to location and person but not date, with delaying in following complex tasks. The patient's speech was slurred and hoarse; there was a paucity of output. The patient's cranial nerves were intact. The patient was unable to hold their arms up for more than 2-3 seconds. The patient's proximal arm strength was scored as 2-3/5 while they had no movements of the lower extremities. The patient's sensory exam was unreliable and inconsistent.

The patient presented to an OSH with confusion and hypoxia. A diagnostic test was performed and the patient was subsequently discharged to home with hospice. The patient died shortly thereafter (age XX, less than 3 years after presentation) and an autopsy was performed.

**Example 7:**

Neuropathological definitive diagnosis: Right subdural organizing hematoma, acute on chronic

DIAGNO/Neurologist Dx:

|  | Top 1 Dx | Top 2 Dx | Top 3 Dx |
| --- | --- | --- | --- |
| DIAGNO | Cerebral Hemorrhage | Uremic Encephalopathy | Sepsis-Associated Encephalopathy |
| Neurologist | Subdural hematoma | Intraparenchymal hemorrhage | Epidural hematoma |

Case summary: A XX-year-old individual became somnolent while at a clinic visit. The patient had flown from Mississippi to Boston the previous day, and was traveling to have a dermatology consultation regarding skin lesions over their bilateral thighs and buttocks, which had been present for the past year. The patient had been seen on the day of their arrival in Boston for another medical consultation at MGH where no mention was made of neurologic deficit. The patient had reported increasing pain from their skin lesions that required escalating doses of pain relief medicine. The patient also reported nausea attributed to a medical therapy for their skin lesions and muscle twitching that was attributed to a medical therapy for pain.

The patient's past medical history was notable for end-stage renal disease due to uncontrolled hypertension, and the patient was on peritoneal dialysis. Previously, the patient had rejected a living related-donor renal transplant, and was currently being evaluated for a second transplant. The patient had also had a remote mitral valve replacement and was being treated with an anticoagulation medicine. The patient's skin problems had started the previous year with painful, bilateral hip nodules that progressed to ulcers and later extensive black eschars. A biopsy of the lesions was performed, and the patient was started on a medical therapy as treatment. The patient had been referred to MGH for a multidisciplinary second opinion on these skin lesions.

The night before the patient's MGH dermatology appointment, the patient had fallen, one of several recent falls attributed to their use of pain medicine. The following day, the patient was noted to have difficulty walking into the dermatology exam room, and then rapidly became progressively more somnolent, and was quickly arousable only to sternal rub. The patient had brief “seizure-like” activity on the exam table, and EMS was activated to arrange transport to the ED. The patient's partner reported the patient was complaining of a headache earlier that day. En route to the ED, the patient developed respiratory failure requiring bag mask ventilation. A medical therapy was discontinued and a treatment was provided to improve the patient's respiratory status, but the patient continued to be intermittently apneic and ultimately required intubation.

Prior to intubation, the patient's neurologic exam was notable for anisocoria with a fixed and dilated right pupil, but the patient was moving all of their extremities in response to noxious stimulation. After a study was performed, several emergent therapeutic treatments were initiated and additional consultation was obtained. Repeat neurologic examination was notable for right pupil fixed and dilated (~7mm), left pupil dilated (~4mm) and minimally reactive, absent brainstem reflexes and no response to noxious stimuli. Due to the patient's poor exam and significant medical comorbidities, the overall prognosis was thought to be grave and no escalation of care was sought by the partner. The patient was transferred to the Neuro ICU, and was then transitioned to comfort measures. The patient died within 24 hours of their initial presentation and an autopsy was performed.

**Example 8:**

Neuropathological definitive diagnosis: Corticobasal Degeneration

DIAGNO/Neurologist Dx:

|  | Top 1 Dx | Top 2 Dx | Top 3 Dx |
| --- | --- | --- | --- |
| DIAGNO | Amyotrophic Lateral Sclerosis (ALS) | Multiple System Atrophy (MSA-P for parkinsonian variant) | Progressive Supranuclear Palsy (PSP) |
| Neurologist | Corticobasal syndrome | Progressive supranuclear palsy | Amyotrophic lateral sclerosis |

Case summary: A XX-year-old left-handed patient presented to neurology clinic for evaluation of unsteady gait and tremor. Symptoms had begun two years earlier, when the patient noticed progressive numbness in both legs, right more severe than left, which had spread up to the knees. This was associated with a sense of unsteadiness but had not caused any major falls. During this time the patient also noticed the onset of a tremor in their hands, left worse than right. Their handwriting had worsened and they had trouble holding a glass of water without it shaking. The tremor worsened during times of anxiety or stress.

The patient's past medical history included hypertension, lumbar disc disease, chronic diarrhea, two spontaneous pneumothoraces, and a history of heavy alcohol use with ongoing but reduced consumption at the time of presentation. Medications included a beta-blocker, an antihypertensive medication, aspirin, and an anti-anxiety medication. Family history was notable for a maternal uncle with ALS and no other known neurologic disease.

On exam, the patient had normal mental status and a normal cranial nerve exam. The patient exhibited an upright posture with a normal facial expression, normal blink rate, and no bradykinesia. The patient did not have a rest tremor, but did have a high-frequency bilateral upper-extremity postural tremor that amplified as they reached for a target. The patient had full strength to confrontational testing and normal tone. The patient had decreased sensation to temperature, pinprick, and vibration sense up to the knees, which was more severe on the right. Romberg testing was abnormal. The patient exhibited a wide-based gait and could not tandem walk. Reflexes were normal and symmetric throughout, except for absent ankle jerk in the left ankle. Toes were downgoing. The patient was treated conservatively and followed.

Over the next year, the patient started using a cane and having falls because their right leg developed a tremor any time they stood on it. The tremor would improve after several seconds of standing. The patient's right knee would occasionally “buckle” during walking. On re-examination the following year, the patient was found to have 4+/5 strength in the right hip flexors and ankle dorsiflexors, normal tone, and slow and irregular foot tapping on the right compared to the left. The patient had circumduction of the right leg while walking. Reflexes were brisk and the patient had spread in the pectoral muscles, cross adduction at the patella bilaterally, and 3 beats of clonus in the right ankle. Right toe was extensor. There was no jaw jerk. A range of imaging and other diagnostic studies were performed. The patient was started on a medication that provided minimal improvement in their gait.

Two and a half years after the first assessment, the patient's gait had significantly worsened and they started using a walker. The patient could not stand up without the use of their arms. Their posture was stiff but upright. The patient had difficulty initiating steps and their steps became smaller. The patient always led steps with their left leg and had difficulty picking up their right leg. Turns were cautious and done en-bloc. The patient's motor exam was unchanged however. Another medication was begun but without benefit.

Four years after presentation, the patient had developed increased bilateral upper arm and right leg tone, worsening right leg weakness (3+/5 hip flexion and foot dorsiflexion), and severe gait difficulty that led them to become wheelchair bound. The patient required two people for transfers and could only take small shuffling steps. The patient tended to fall backward. The patient also had some use-dependent tremor of their right arm that was more pronounced than on the left. Rapid movements of their right arm were slower than the left, and the patient had difficulty initiating movements of their right leg. Their cognition, speech and swallowing, and bowel/bladder function remained intact. There was no astereognosis, agraphesthesia, or extinction to double simultaneous stimulation. The patient had two more neuroimaging studies that were abnormal.

Over the next three years, the patient's symptoms continued to progress. The patient developed dysarthria; the weakness and spasticity of both the right arm and the right leg worsened; their left arm began to have symptoms as well. The patient also developed urinary retention. The patient had difficulty initiating purposeful movements with their right arm and leg; when they would move their right arm they would induce a severe, high-amplitude shaking. Occasionally their right leg would elevate from the bed involuntarily. The patient also had occasional large-amplitude jerking of their limbs that would awaken them at night.

Nine years after the onset of symptoms, the patient was communicating primarily by blinking their eyes. The patient had difficulty swallowing, had a poor appetite, and had minimal purposeful movement. After discussion with their family, the patient was transitioned to comfort care. The patient died at home and an autopsy was performed.

**Example 9:**

Neuropathological definitive diagnosis: Glioblastoma

DIAGNO/Neurologist Dx:

|  | Top 1 Dx | Top 2 Dx | Top 3 Dx |
| --- | --- | --- | --- |
| DIAGNO | Metastatic Brain Tumor | Progressive Multifocal Leukoencephalopathy (PML) | Paraneoplastic Syndrome |
| Neurologist | Glioblastoma multiforme | Brain metastases | Primary CNS lymphoma |

Case summary: A XX-year-old patient was recognized by their partner to have an altered speech pattern. The partner found that the patient was speaking very slowly and deliberately. That day, the patient developed difficulty typing at work, trouble with their balance, and headaches that they related to their sinuses. The next day, the patient had difficulty driving, taking turns too wide and almost went off the road. The patient's symptoms continued to worsen and the next morning they felt so clumsy they lost balance twice, fell into a dresser and were unable to begin to get dressed due to difficulty with coordination in their left arm and leg. At this juncture, the patient was taken to an OSH, from which they were rapidly transferred to MGH.

The patient's past medical history was significant for a spindle cell neoplasm of the breast diagnosed XX months previously and treated with localized excision. Other history included hypertension, COPD, and stage 2 chronic kidney disease. The patient did not smoke at the time of presentation, but had smoked a pack a day from the age of XX to the age of XX. The patient drank one to two drinks per night.

When the patient arrived at MGH, they had a simple partial seizure witnessed in the ED and a therapeutic intervention was given. The exam at the time of presentation at MGH was significant for left sided facial droop at rest, lack of shoulder shrug on the left, and 0-1 motor strength in the left upper and lower extremities compared to 5/5 strength on the right. Imaging had been performed at the OSH, with additional studies done after arrival at MGH. An intervention was then performed.

Following this intervention, the patient had worsened left sided motor function (initially complete plegia), although this improved over the subsequent months with rehab treatment. The patient was followed as an outpatient over the next XX months with relative stability of their symptoms and exam.

Approximately XX months after the patient's presentation to MGH, they began to develop headaches and were noted by their partner to be “slower” than previously. These symptoms continued to slowly progress over the next couple months with increased weakness to the point that the patient became dependent on a cane to walk. A range of new interventions were pursued, with enough improvement that the patient was able to take a trip with their family to the Grand Canyon without incident.

The patient's symptoms continued to fluctuate for the next couple of months with continued headaches and occasional episodes of losing their balance. The patient complained of significant fatigue, progressive increasing left sided weakness and deterioration of their short term memory. The exam at this time was significant for flat affect, left sided pronator drift, and 4/5 strength in the left upper and lower extremities. The patient's gait was wide-based and hesitant.

The patient continued to deteriorate and home hospice was initiated. The patient died XX months later and an autopsy was performed.

**Example 10:**

Neuropathological definitive diagnosis: Cerebral amyloid angiopathy (CAA)

DIAGNO/Neurologist Dx:

|  | Top 1 Dx | Top 2 Dx | Top 3 Dx |
| --- | --- | --- | --- |
| DIAGNO | Traumatic Brain Injury (TBI) | Alzheimer's Disease | Intracerebral Hemorrhage |
| Neurologist | Cerebral amyloid angiopathy-related lobar hemorrhage | Hypertensive intracerebral hemorrhage | Post-cardiac arrest encephalopathy |

Case summary: A XX-year-old individual was found down at home by their son. The patient was last seen well by their partner a day earlier. The patient had helped their partner pack up the car for a weekend trip to the beach, but did not join them on the trip. The partner spoke to the patient on the phone a few times later that day, with the last conversation that evening. The next morning, when the partner called home at 10:30AM, the patient did not answer the phone. A few more attempts were made, and concerned because they were unable to contact the patient, the partner asked their son to check on them. When the son entered the house, he found the patient in their pajamas, face down on a carpeted floor. There was blood on the closet door and the pillow. EMS was called and the patient was brought to an outside hospital. On arrival there, GCS was 3. The patient was intubated for airway protection, imaging was performed, therapy was begun and the patient was transferred to MGH ED for further care.

The patient's past medical history was notable for arthritis, GERD, and elevated PSA (numerous biopsies without evidence of malignancy). The family history was notable for both parents having been diagnosed with Alzheimer disease in their 70s. The patient had retired from the judicial bench less than a year before presentation. The patient drank wine 2-3 times per week and was a non-smoker. The only medication at the time of presentation was ibuprofen; the patient was previously on a medication for prostate issues but had discontinued it several months prior to presentation.

Additional history was obtained at MGH, which included about a year of subjective memory concerns. Given the family history, this had made the patient concerned that they were developing Alzheimer disease like their parents, so the patient and their partner had met with a lawyer to sign a living will about two months prior to presentation. There had also been an isolated episode of left sided facial pain 6 months prior to presentation. It was anterior to left ear, worse when chewing. It had resolved with a course of ibuprofen.

The patient's exam on presentation to the MGH ED was notable for mild bradycardia to 57 and mild hypotension to 102/55 (on sedation). The patient was afebrile and oxygen saturation on the ventilator was 100%. General exam was notable for an abrasion on the left forehead, periorbital ecchymoses on the left and multiple ecchymoses on the left side of the body. The patient was intubated with a C-collar in place. There was a loud abdominal bruit, with a pulsatile palpable aorta. Distal pulses (dorsopedal and popliteal) were intact.

The patient's neurologic exam ‎showed them to be unresponsive to voice or noxious stimuli, to not follow any commands but grimace to noxious stimuli. The pupils were weakly reactive, going from 2mm to 1.5 mm bilaterally. Oculocephalic testing was not performed as the patient was still in a C-collar. Corneal reflexes were weak bilaterally, gag reflex was absent but cough reflex was present. Tone was increased and the patient was rigid in their lower extremities bilaterally. The right upper extremity withdrew to stimulation, associated with high frequency shaking of the arm (this shaking was later noted to occur independent of stimulation). The left upper extremity flexed with noxious stimulation, the right lower extremity withdrew to noxious stimulation and there was no movement of the left lower extremity. The reflexes were notable for up-going toes bilaterally. The patient was also noted to have intermittent bilateral shaking of their extremities. Additional diagnostic studies were performed and additional therapy was started.

After therapy the pupils were bilaterally reactive 2mm->1mm and the patient developed extensor posturing in left upper and lower extremity to noxious stimulation (GCS 6T) and their left eye would open slightly with noxious stimulation. Repeat diagnostic imaging was performed. A family meeting was held, and in accordance with the patient's previously stated wishes, care was transitioned to intensive comfort measures.

The final neurologic exam before the patient was extubated found them lying in bed with their eyes closed. The pupils continued to be reactive (2mm to 1mm), with intact corneal reflexes. Right upper and lower extremities withdrew to painful stimuli, while the left upper extremity showed extensor posturing and the left lower extremity showed triple flexion. The patient died shortly after extubation (five days after presentation to the ED) and an autopsy was performed.

**Example 11:**

Neuropathological definitive diagnosis: Spinocerebellar ataxia (SCA)

DIAGNO/Neurologist Dx:

|  | Top 1 Dx | Top 2 Dx | Top 3 Dx |
| --- | --- | --- | --- |
| DIAGNO | Multiple System Atrophy (MSA-P for parkinsonian variant) | Progressive Supranuclear Palsy (PSP) | Hereditary Spastic Paraplegia (HSP) |
| Neurologist | Spinocerebellar atrophy (SCA) | Dentato-rubro-pallydo-luysian atrophy (DRPLA) | Hereditary spastic paraparesis (HSP) |

Case summary: A XX-year-old right-handed patient presented to neurology clinic for abnormal gait. The symptoms started two years prior to presentation, when the patient noticed some difficulty walking. The patient noticed that they “lurched to the right or left” and had a stiff gait. The symptoms waxed and waned and worsened in the dark. They had been roughly stable for a year when the patient presented in clinic, and the patient did not complain of any other neurologic symptoms, including diplopia or difficulty with fine finger movements.

The patient was a substitute teacher with no significant past medical history other than some insomnia and anxiety; they took no medications. Family history was notable for abnormal gait in the patient's mother, maternal grandmother, and brother. The patient was unmarried, lived independently and did not smoke, drink, or use any illicit drugs. The patient enjoyed going to the gym.

On initial exam, the patient demonstrated a normal mental status. The patient did not have difficulty with comprehension, production, or repetition of written or verbal language. However, the patient's speech sounded dysarthric. The patient's lingual, labial, and pharyngeal sounds were uttered normally, but the articulation of spontaneous speech sounded abnormal. Prosody was also abnormal. On cranial nerve exam, the patient had a lid lag and retraction. Pupils were normal. The patient's extraocular movements were full and only notable for non-sustained nystagmus on lateral end-gaze. The rest of the patient's cranial nerves were normal. The patient's muscle power and bulk were normal in all limbs, but tone was increased in both legs. There was no resting tremor or adventitious movement. Sensation was normal to pin and vibration throughout. The patient's reflexes were brisk throughout. There was no clonus and toes were downgoing. The patient had some subtly slow and inaccurate movements on finger-nose testing that became clearer with repeat testing. Heel-to-shin was normal, and rapid alternating movements were mildly slow. The patient was able to stand steadily and only swayed a little when their eyes were closed. The patient's gait was stiff and it appeared that their right leg did not lift off the ground as quickly as the left leg. The patient could stand on each foot for 15 seconds, but could not sustain a tandem gait for more than 3 steps.

One year later, the patient was re-examined and noted to have saccadic breakdown of left eye movements during tracking and vertical diplopia on upgaze. The patient developed clonus of the right ankle and Hoffman’s sign bilaterally. The patient was no longer able to stand on one foot or in tandem. The patient's walking had worsened. The patient noted that symptoms were much worse if they were trying to start walking after standing or sitting still for some time; symptoms would improve with continued walking. The patient was started on a medication with some improvement in symptoms.

The patient received their care elsewhere over the next several years, until six years after presentation (eight years after onset of symptoms) when the patient presented to an OSH for pain from muscle spasms. There the patient was noted to have a fever, diaphoresis, malaise, and episodes of high muscle tone with clenched extremities. The patient was initially awake, oriented, and conversant. The patient had a WBC of 21.4. CK was 1000. Medical management was begun but the patient's CK rose to 5000 and their mental status deteriorated. The patient was transferred to MGH. Shortly after arrival, the patient's mental status deteriorated further. The patient was intubated for airway protection and sent to the ICU. The patient had an extensive workup, was treated for pneumonia and received a series of other empiric treatments. While in the ICU, the patient had profound transient episodes of diaphoresis, tachycardia, and hypertension.

The patient's exam in the ICU was poor. The patient's eyes were partially open and they did not fix, track, or blink to threat. The patient did not follow commands or withdraw to noxious stimulation. Pupils were equal and reactive and corneal reflexes were intact, but there were no eye movements with passive head rotation. The patient's head and neck were held in extension, which became exaggerated with stimulation. The patient's tone was variable; with stimulation the patient often developed lead-pipe rigidity with a superimposed catch in the arms. Both feet were held in plantarflexion. Reflexes were brisk throughout and there was clonus in both wrists, L>R. After several failed treatments and two weeks in the ICU, the patient was trialed on multiple medications with minimal improvement. After one month in the hospital without improvement, the patient's care was transitioned to focus on comfort. The patient died and an autopsy was performed.

**Example 12:**

Neuropathological definitive diagnosis: Diffuse Large B Cell Lymphoma (DLBCL)

DIAGNO/Neurologist Dx:

|  | Top 1 Dx | Top 2 Dx | Top 3 Dx |
| --- | --- | --- | --- |
| DIAGNO | Multiple System Atrophy (MSA-C for cerebellar variant) | Progressive Supranuclear Palsy (PSP) | Paraneoplastic Syndrome |
| Neurologist | Primary CNS lymphoma | Autoimmune encephalitis | ADEM |

Case summary: A XX-year-old individual became confused, with slurred speech and difficulty using their right hand two days after knee surgery. These symptoms lasted less than a day, but the patient began to complain of dizziness and nausea. The patient described that, while lying down they felt as if they were on a boat and when standing up they felt unsteady. The patient was seen by a neurologist and was given a medication for dizziness, which did not help. About three months after the knee surgery, the patient had a severe headache which required aggressive medical management. Two months later, the patient experienced visual changes, described as “ring of smoke” involving both eyes. The patient saw an ophthalmologist and was diagnosed with uveitis and had an inferior defect on their left eye visual field. The patient underwent scheduled follow-up imaging; abnormalities prompted an admission to MGH. The patient's past medical history included migraine, essential tremor, HTN, and bilateral hearing loss. The patient's mother had dementia, possibly AD, and father was a smoker and died of bladder and lung cancer.

On admission, the patient's mental status, concentration, attention, and memory were intact. The patient's visual fields were intact to confrontation and cranial nerve exam was unremarkable. The patient's speech, strength, and reflexes were normal; intention tremor was prominent. There was mild upper extremity dysmetria on the right, but intention tremor in the left arm and slight dysmetria on the right. The heel-to-shin was normal bilaterally but there was impaired rapid tapping of the right leg on the ground. The patient's gait was mildly unstable and they could not tandem gait or stand with feet in the tandem position.

The patient's CXR showed patchy opacities in the lungs predominantly in the right lower lung and left lung base. The patient's CSF showed a total protein of 64, WBC 6; RBCs 0, IgG index was normal. Cytology was negative for malignancy. A repeated CSF showed protein of 43 and WBC 7 with unremarkable cytology and flow. HIV was negative. Other labs from serum revealed a normal ESR/CRP and LDH, negative ANA, Anti-Ro, Anti-La, and RF. The patient's ACE was 26. A range of non-CNS imaging studies were normal. A diagnostic procedure was performed and management begun. The patient was discharged home with partial improvement of their nausea, as they were treated with medications for nausea and anxiety.

Two months later, the patient was readmitted due to worsening of their balance and gait with frequent falls, more towards the right. The patient had started to use a walker to ambulate. The patient was alert and attentive with intact language, naming repetition, reading, and writing. The patient's affect was now tearful and concerned and they described their mood as ""depressed"". On neurologic exam, the patient had nystagmus when fixating on a finger without extinction but not on far gaze; there was also horizontal nystagmus with end gaze in all directions. The patient's tremor was most prominent in the left hand and was present when they stretched out their hands. The patient now had dysmetria on finger-to-nose and heel-to-shin bilaterally but worse on the left. The patient was able to stand unassisted; their gait was narrow-based with the walker and unsteady (near fall to the right) without it. New abnormalities were present on the patient's imaging studies and new treatment was started.

The patient had multiple admissions over the ensuing months. There were fluctuations in their mental status (loss of recall of their history and impaired orientation). The patient's nystagmus continued to be present, with some degree of complaints of double vision on lateral gaze as well. The patient developed a moderate right facial droop with intact sensation and mild right masseter weakness. The patient began to have a tendency to fall backwards when standing. On subsequent admissions, the patient was found to have difficulty with motor sequencing tasks (right worse than left) and to have some difficulty coordinating their steps when using the walker. The patient eventually had difficulty rising to a standing position, had a wide-based stance with the walker, and could only take a few wide-based shuffling steps with the walker. Unfortunately, during one admission, the patient developed jerky left arm and leg movements and long periods of unresponsiveness. The patient was hypotensive to 70s, tachycardic to 120's, prompting acute treatment and intubation. Imaging studies showed new findings. The patient's CSF showed protein of 134 with 2 cells and normal glucose. The patient eventually required a tracheostomy and PEG. At discharge, the patient would open their eyes to voice, tracked and regarded the examiner. The patient was able to follow axial and appendicular commands. The patient had bilateral weakness: only able to move their right thumb, localize noxious stimuli on their left arm, and wiggle their toes bilaterally.

About XX months after the first assessment, the patient returned to MGH after further decline. At that time, the patient would only rarely lift their head to kiss their partner and attempt to mouth some words but was mostly asleep. The patient had weak withdrawal to nail bed pressure in the upper extremities and brisk withdrawal in the lower. There was again worsening of the patient's imaging. After family discussions, the patient was transitioned to hospice care with focus on comfort only. The patient died a few days after that at age XX and an autopsy was performed.

**Example 13:**

Neuropathological definitive diagnosis: Cerebral amyloid angiopathy (CAA)

DIAGNO/Neurologist Dx:

|  | Top 1 Dx | Top 2 Dx | Top 3 Dx |
| --- | --- | --- | --- |
| DIAGNO | Intracerebral Hemorrhage (ICH) | Ischemic Stroke | Subarachnoid Hemorrhage (SAH) |
| Neurologist | Left middle cerebral artery territory infarction with hemorrhagic transformation | Cerebral amyloid angiopathy-related hemorrhage | Alzheimer's disease |

Case summary: A XX-year-old individual was playing golf when they became unable to respond to questions, had difficulty finding words and was dysarthric. The patient was rapidly brought to an outside ED where they were seen 40 minutes after onset of symptoms. Their past medical history was notable for mild dementia, hypertension, aortic valve replacement (porcine valve) and coronary artery disease. Their medications at home included an antiplatelet medication, a diuretic, a beta-blocker, a medication for benign prostatic hyperplasia, potassium, an antidepressant, and a sleep aid. The patient had no prior history of previous similar episodes and there was no family history of neurologic disease. They were not a smoker.

On the first exam at the outside institution, their blood pressure was 156/64. The patient was found to have a receptive and expressive aphasia, with difficulty understanding questions, although they were able to follow all commands. Laboratory findings include glucose 98, INR 1.2, platelets 181K and Cr 1.4. Imaging was performed and a therapeutic treatment was sought. After initial improvement of symptoms, the patient began to feel ill, with worsening headache and vomiting emerging within about 60 minutes of initiation of treatment. Another study was performed. The decision was then made to transfer the patient to MGH. Prior to transfer, they further declined and became agitated, requiring intubation in association with receiving 250 ug of fentanyl and 10 mg of versed.

On arrival at MGH (approximately 4 hours after the start of their abrupt decline) the patient was somnolent with pinpoint pupils, localizing painful stimuli with their right arm only, and they were transferred to the neuro ICU where they received multiple therapeutic interventions – some of which had been started during the transfer. Their exam off sedation the following morning showed little change with pupils equal and reactive 2-->1.5 mm, roving eye movements, but not tracking. The patient was localizing to pain in the left upper extremity and the right was withdrawing but with less movement. Bilateral lower extremities withdrew to painful stimuli. Following a family meeting the decision was made to transition care to comfort measure only. The patient died the next morning and an autopsy was performed.

**Example 14:**

Neuropathological definitive diagnosis: Cerebral amyloid angiopathy, severe

DIAGNO/Neurologist Dx:

|  | Top 1 Dx | Top 2 Dx | Top 3 Dx |
| --- | --- | --- | --- |
| DIAGNO | Acute Ischemic Stroke (likely Left MCA territory) | Vascular Dementia | Hypertensive Encephalopathy |
| Neurologist | ICH due to CAA (intracerebral hemorrhage due to cerebral amyloid angiopathy) | ICH due to underlying hemorrhagic tumor (ex. lung cancer metastasis) | "Left sided stroke (probably posterior circulation/brainstem) |

Case summary: A XX year-old patient was found unresponsive in their car by the police. The patient had been feeling unwell for several days and arranged to be seen by their PCP. The patient was last seen well by their family the night before their doctor's visit. No information is available about the exam in the PCP's office. While driving home from this visit, the patient began to feel dizzy and confused. The patient pulled over to the side of the road and called EMS. When the police arrived, the patient was in their car and unresponsive. There were no reported signs of trauma.

The patient lived independently and was fully functional with all activities of daily living. By report, the patient was taking no medications. The patient's daughter-in-law reported that there had been some mild cognitive issues with memory and word-finding difficulty that were slowly progressive over the last year.

The patient was brought to an outside hospital ED where their mental status continued to decline. The patient's hematocrit was 35, platelets 264, INR 0.9, and creatinine 0.8. The patient's systolic blood pressure was in the 180's and their heart rate declined from the 80's to the 50's. Imaging studies were abnormal. Therapy was initiated and the patient was transferred to MGH by MedFlight.

On arrival at MGH, the patient was afebrile, hypertensive (164/72) and bradycardic (56). Pupils were reactive bilaterally. Corneal, cough and gag reflexes were present. The patient's left upper extremity localized to noxious stimuli and their right upper extremity did not respond; the patient's left lower extremity withdrew and their right lower extremity triple flexed. The patient was admitted to the neuro ICU. Additional imaging studies were performed and were abnormal.

During the course of the patient's stay in the ICU they remained critically ill. The patient was euthermic, eunatremic and euglycemic. EKG showed a first degree AV block. On exam about 18 hours after admission, the patient opened their eyes to voice, tracked and regarded the examiner. The patient followed no commands. The patient's left arm and leg moved spontaneous and antigravity while the right leg showed non-stereotyped withdrawal and their right arm was flaccid.

The following day, the patient's eyes opened to voice, gazed toward the midline, and their pupils were reactive (3 mm to 2 mm). The patient did not follow axial or appendicular commands and did not mimic. The patient had a right facial paralysis. The patient's left upper extremity localized and the left lower extremity withdrew; the right lower extremity triple flexed and the right upper extremity was mute.

Following family discussions on goals of care and according to the patient's pre-stated wishes the patient was transitioned to comfort measures only. The patient died 2 days later (5 days after their initial presentation) and an autopsy was performed.

**Example 15:**

Neuropathological definitive diagnosis: Primary CNS Lymphoma (PCNSL), Diffuse Large B-cell Lymphoma subtype

DIAGNO/Neurologist Dx:

|  | Top 1 Dx | Top 2 Dx | Top 3 Dx |
| --- | --- | --- | --- |
| DIAGNO | Paraneoplastic Syndrome (Paraneoplastic Encephalitis/Encephalomyelitis) | Chronic Inflammatory Demyelinating Polyneuropathy (CIDP) | Alzheimer's Disease |
| Neurologist | Lymphoma (with CNS involvement and neurolymphomatosis) | Paraneoplastic syndrome (ex Anti-Hu) | Tumefactive MS |

Case summary: A XX-year-old individual was found unconscious in the shower by their home health aide. By report from their children, the patient had been having increasing memory difficulties and difficulties with activities of daily living (late to appointments, forgetting to pay bills) over the previous several months. The patient was taken to an outside ED, where EKG and imaging studies were unremarkable. The patient was admitted, received hydration and had their medications adjusted. Their neurologic examination was notable only for right foot drop and the patient was discharged to a rehab facility for continued care and physical therapy. While at the rehab facility, their mental status declined (no details available) and the patient was noted to be hyponatremic. Assessment by endocrinology suggested SIADH, which was managed medically. With correction of their Na to 135, their mental status did not clear. Their neurologic difficulties prompted repeat imaging, followed by transfer to MGH.

Their past medical history includes onset of right leg pain approximately XX years earlier (shooting from their hip down their leg) accompanied by bilateral numbness and pain in their feet. This responded to physical therapy and was well controlled for XX years until the right leg pain returned. About XX months later the patient developed right foot drop. When examined, the patient was alert and oriented X 3 with no apparent difficulties with memory, comprehension, or ability to follow commands. There was atrophy of their right hand involving extensor digitorum brevis, first dorsal interosseous, and abductor digiti minimi. No fasciculations were noted. Their power (R/L) was noted to be 4+/4+ shoulder abductors, 1/4+ abductor digiti minimi, 1/4+ inferior oblique, 4+/5- abductor pollicis brevis, 4+/4+ hip flexion, 0/5 ankle dorsiflexion, and 0/5 ankle eversion. On sensory exam the patient had reduced vibration sense in the legs to the knees bilaterally and reduced pin prick and soft touch on the dorsum of the right foot on the lateral aspect. Reflexes were absent at the right knee and 2+ at the left knee and were absent at the ankles bilaterally. Reflexes were present and symmetric in the upper extremities. Plantar responses were flexor. The patient was also recognized to have a possible right thigh mass. This was biopsied, and while not having a specific histologic diagnosis there was evidence of clonal IgH rearrangement. No specific therapy was initiated, although the patient was treated with neurontin and methadone for their pain.

On admission to MGH, their neurological examination was notable for being awake with eyes closed and oriented to MGH and year but not month or date. The patient wasn’t able to name months backwards past December. The patient was able to follow one-step commands but had trouble with two-step commands (when asked to touch their right thumb to their left ear, the patient touched their right cheek). The patient also had difficulty initiating tasks. Their speech was slow and at times the patient seemed to lose their train of thought. Their recall of named objects at 2 minutes was 0/3. The patient was unable to demonstrate teeth brushing with either hand and could not voluntarily track fingers with their eyes. Their cranial nerve exam was unremarkable. Motor testing showed strength at 4+/5 throughout except their right leg, which was 3/5 on hip flexion, 3+/5 knee with ext/flex and 0/5 DF and 4/5 PF.

On admission, their Na was 126; over the hospitalization, it continued to fluctuate between 125 and the mid-point of the normal range. An LP on admission showed glucose 54, protein 42, 2 RBC, 40 WBC (85% lymphs, 11% mono) although the cytology showed some atypical forms. Other studies were sent on the CSF. Additional imaging studies were performed; chest imaging suggesting pulmonary emboli, prompting the initiation of an anticoagulant.

Eleven days after admission to MGH, an additional diagnostic procedure was performed, and additional medical management was begun. By the following day, their cognition appeared to have improved such that the patient was described as bright and alert, remembering that the day was a holiday. The patient continued to improve and additional therapy was begun XX days later, followed by another medication XX days after that.

On the evening that the third medication was initiated, the patient was noted to be more somnolent than usual in the evening. As the night progressed, the patient continued to be sleepy and their nurse noted that their pupils had become more sluggish. When the on-call resident arrived at the bedside, the patient’s pupils were fixed and the patient was unarousable. In accordance with their prior stated wishes, the patient was not intubated and did not have active resuscitation measures. Shortly thereafter, when apneic and pulseless, the patient was declared dead (XX days after admission to MGH) and an autopsy was performed.

Supplementary Note 7. Fifteen Examples of Cases where Both DIAGNO and Neurologists were Incorrect.

**Example 1:**

Neuropathological definitive diagnosis: Alzheimer disease neuropathologic change

DIAGNO/Neurologist Dx:

|  | Top 1 Dx | Top 2 Dx | Top 3 Dx |
| --- | --- | --- | --- |
| DIAGNO | Intracerebral Hemorrhage (ICH) | Ischemic Stroke (likely embolic) | Cerebral Amyloid Angiopathy (CAA)-related Hemorrhage |
| Neurologist | Intraparenchymal hemorrhage, likely related to cerebral amyloid angiopathy | Reversible cerebrovascular vasoconstriction syndrome | Metastatic brain tumors |

Case summary: A XX-year-old right-handed individual awoke with a severe headache. The patient took a medication for the pain. The patient subsequently noticed they were misreading their watch and soon thereafter experienced vomiting. The patient was admitted to an outside hospital where it was noted they had left sided neglect and a left homonymous hemianopia. Imaging showed an abnormality. The full details of this admission were not available for review, but the patient was told to avoid a medication in the future. The patient was discharged to rehab and their neglect improved but they had a persistent left homonymous hemianopia.

The patient's past medical history included coronary artery disease, bradycardia, and BPH. The patient did not have either hypertension or diabetes. The patient retired three years before the initial presentation, having worked in IT. The patient lived alone, had no tobacco exposure and limited alcohol intake. The patient's father had died of a heart attack and ""ischemic stroke"" (per the patient) at age XX while their mother died at age XX with congestive heart failure. Both of the patient's parents had demonstrated mild memory impairment prior to death. The patient had one brother who was alive with chronic mental illness (not otherwise described) but no neurologic diagnosis.

At age XX, the patient had a ""very slight headache"" while on the way to a dental appointment. The patient sat through a 3 hour dental procedure then drove home. The patient started to experience bilateral arm ""heaviness"" and on arrival home had 2 episodes of vomiting. The patient was concerned that they could be having an MI after reading about their symptoms online so they activated EMS. The patient was taken to an outside hospital where imaging again showed an abnormality. The patient was transferred to MGH, where they had persistent but mild bifrontal headache and resolution of nausea. The patient's systolic blood pressure was less than 140 without intervention. The patient's examination demonstrated no sensory or motor deficits apart from the previously documented dense left homonymous hemianopia.

The patient was seen in clinic at MGH shortly after this hospitalization. The patient's neurologic exam was normal except for deficits in the left visual field (then documented as being unable to detect motion in the left lower quadrant, some motion detection in the left upper quadrant), minimal stumbling on tandem gait, and a left-sided palmomental response.

Subsequently, the patient experienced slurred speech, visual disturbance, reportedly right sided weakness, and headache that came on suddenly while they were volunteering at a soup kitchen. The patient went to an outside hospital and on arrival at the ED, new imaging abnormalities prompted admission.

Soon after arrival, the patient became more somnolent and required intubation for airway protection. Repeat imaging demonstrated an evolving abnormality. The patient was admitted to the ICU, with various services consulted. A series of interventions were attempted, without significant benefit.

The day after admission, the patient's neurologic examination demonstrated that they were able to follow commands. The patient was also able to track the examiner in the horizontal plane. The patient had equal and reactive pupils and intact corneal reflexes. The patient had slightly dysconjugate gaze and impaired upgaze. The patient was densely hemiplegic on the left and with additional weakness on the right (leg > arm). The patient demonstrated no reaction to noxious stimulation in any extremity. The patient's reflexes were 1+ in left arm and 3+ in arm. The patient's toes were upgoing bilaterally. The patient died the next day, age XX, and an autopsy was performed.

**Example 2:**

Neuropathological definitive diagnosis: Corticobasal Degeneration (CBD)

DIAGNO/Neurologist Dx:

|  | Top 1 Dx | Top 2 Dx | Top 3 Dx |
| --- | --- | --- | --- |
| DIAGNO | Frontotemporal Dementia (FTD) | Progressive Supranuclear Palsy (PSP) | Alzheimer Disease (AD) |
| Neurologist | Progressive supranuclear palsy | Behavioral variant frontotemporal dementia | Behavioral variant Alzheimer's disease |

Case summary: A XX-year-old individual began having trouble handling pressure at work. At that time, the patient was a successful special education teacher. This was followed by a subjective sense of deteriorating memory and overall increased distress in the setting of being the primary caregiver for their partner with Parkinson disease. The patient also found that they were having increased difficulties planning activities and in certain situations such as traveling, as they needed to check and re-check details. The patient also started to develop other obsessive-compulsive behaviors such as re-washing and re-folding loads of laundry. Eventually, stress and anxiety prompted the patient to retire from teaching by age XX.

The patient's past medical history included hypertension, elevated cholesterol, classic migraine, rosacea, and bilateral hearing loss corrected with hearing aids as well as ADHD-inattentive type, depression, and generalized anxiety. Medications included an antihypertensive medication, a cholesterol-lowering medication, an antidepressant, another antidepressant, a topical antibiotic, an antibiotic, and a medication for migraines (prn). The patient had a graduate degree in education and had worked as a special education teacher until their recent retirement. The patient's mother had died of a cerebral hemorrhage (age XX) while their father died of unknown causes (age XX). There was an uncle with a diagnosis of Alzheimer disease (age XX). The patient's partner had Parkinson disease, and their two children were alive and well.

At age XX, the patient was seen in the neurology outpatient clinic. The patient's family described them as distractible, processing things slowly, and seeming to have difficulty finding their words. Additionally, the patient would exhibit unusual behaviors – angry outbursts and swearing. The patient had diminished social interest, warmth, and emotional responsiveness. The patient had recently sideswiped another car but did not tell anyone about it until the next day, when they told their partner in a ‘matter of fact’ manner without a sense of urgency or emotion. There was at least one occasion when the patient got lost and disoriented when driving a familiar route home from the airport.

On examination, the patient appeared well kept, made good eye contact, had a slightly tangential thought process with no evidence of hallucinations or delusions. The patient did not have insight into the changes in their behavior or personality. The patient scored 26/30 on MMSE with some impairment of executive function and attention. The patient's fund of knowledge of current events was poor and they recalled none of three words at 5 minutes. Reciting the months of the year backward took the patient 39 seconds, counting down from 20 took 19 seconds (with 1 error) and Trails B took 221 seconds (with 3 sequencing errors). The patient was unable to complete serial 3’s from 20 and they were only able to name 6 “F” words with perseveration on one word. The patient's speech was slow with frequent word-finding pauses and no paraphasic errors. On the Boston naming test, the patient named 56 of 60 items and acquired the other 4 with phonemic cues. The patient's cube drawing was nearly correct. The patient's cranial nerve exam was normal. Meyerson’s reflex was not present. There was a mild fine postural tremor of the fingers; strength and tone were normal. Reflexes were 3+ and symmetric throughout. There was no evidence of ataxia and gait was narrow-based. An imaging study was performed which was abnormal and the patient was started empirically on a trial of medication.

The patient was seen in follow-up about XX year later. The patient's family described progression of prior complaints as well as new problems with balance resulting in recurrent falls, most frequently backward. The patient appeared disheveled, maintained eye contact intermittently, and did not respond to many questions including their name, their partner’s name, and questions about their mood. Occasionally the patient would nod their head when giving multiple-choice answers but was easily distracted, picking lint from their pants and diverting their attention to objects out the window. The patient scored a 7/30 on MMSE. The patient was noted to have bilateral grasp reflexes, mild paratonia in the arms, mild symmetrical bradykinesia. Meyerson’s reflex was now present. The patient was able to stand up from their wheelchair without assistance but with marked retropulsion and had a shuffling gait.

Shortly after this visit, the patient was placed in a long-term care facility. The patient continued to decline and required increasing levels of care with transfers and feeding. The patient had lost a significant amount of weight and spent much of their time in bed, as they were not able to support their own posture consistently when sitting. The patient became effectively mute, estimated to have said about 20 words over 6 months.

The patient died approximately XX years after the onset of their symptoms (age XX) and an autopsy was performed.

**Example 3:**

Neuropathological definitive diagnosis: Cerebral Small Vessel Disease (SVD)

DIAGNO/Neurologist Dx:

|  | Top 1 Dx | Top 2 Dx | Top 3 Dx |
| --- | --- | --- | --- |
| DIAGNO | Progressive Supranuclear Palsy (PSP) | Multiple System Atrophy (MSA-P for parkinsonian variant) | Amyotrophic Lateral Sclerosis (ALS) |
| Neurologist | Neuroacanthocytosis | Neurodegeneration with Brain Iron Accumulation (NBIA) type 1 due to Pantothenate kinase mutation (Hallevorden-Spatz syndrome) | Multiple system atrophy |

Case summary: A XX-year-old left-handed patient fell in the bathroom and hit their head. The patient reported intermittent slurred speech and gait instability for the past year, with frequent falls and some syncopal events. Occasionally, the patient had the sensation that their blood sugar was low, but this was not always the case. The patient's coworkers and friends had noticed their voice become raspier and difficult to understand. At a recent family reunion, family members approached the patient's sibling out of concern that the patient had been drinking. The patient was unaware of any voice changes initially, but then began noticing slurring of their words when fatigued. The patient coughed with eating and drinking, had started using a straw to drink out of concern for choking and bit their tongue frequently. The patient also reported some slowed cognition and experiencing mild bowel and bladder urgency.

The patient's past medical history was notable for obesity status post gastric bypass surgery, diabetes mellitus type 2, and hypercholesterolemia. The patient's medications included insulin, an antihyperglycemic medication, a statin, baby aspirin, a multivitamin, and Vitamin B12 and calcium supplementation. The patient's family history was notable for a maternal aunt and sibling with breast cancer and a mother with pancreatic cancer. The patient had never smoked and drank alcohol occasionally. The patient was a clerical worker for a large company.

The patient's neurological exam revealed them to be mild inattentive with dysarthric speech and a breathy, mildly dysphonic voice. The patient's lingual and buccal movements were slowed with decreased buccal and lingual coordination on repeated vocalizations. The patient's face was symmetrical, but there was some increased facial tone with continuous contraction of the frontalis muscle and decreased blink rate. Jaw reflex was increased. There was a positive glabellar sign. There were no tongue fasciculations or abnormal movements. Gag was diminished. Remaining cranial nerve examination was normal, including intact extraocular eye movements including upgaze and no nystagmus. Motor examination revealed no weakness, but increased tone on the right relative to the left with minor spasticity and decreased fine motor movements. The patient had decreased performance on testing of motor (and cognitive) sequencing tasks. There was mildly impaired coordination with slowed and slightly spastic movements, but no obvious dysmetria. The patient was hyperreflexic throughout. The patient had bilateral positive finger flexor reflexes, grasp reflexes in the toes, but the toes were downgoing. There was no sensory loss to multiple modalities. The patient's gait was wide-based with decreased arm swing on the left side. There may have been some truncal instability. The patient had problems with tandem walking; with difficulty on this task, the patient became emotional and could not control their crying. A range of additional investigations were performed.

Over the next few months, the patient's gait difficulties continued to progress to the point where the patient began using a cane and then a walker to steady themselves. The patient also began reporting incoordination in their upper extremities, dropping objects. The patient's voice became quieter. The patient developed increased emotional lability with inappropriate laughter and crying. Cognitive changes became more prominent as well, such as the patient forgetting how to use the computer at work (despite working there for XX years). Subsequently, worsening cognition and balance forced the patient to leave their job and to stop driving. Bladder and bowel urgency turned to occasional incontinence. The patient reported poor sleep

When seen a year after presentation the patient was noted to be orthostatic. The patient now had undershoot on voluntary saccades with a suggestion of saccadic intrusion on pursuit testing. There was now mildly increased tone throughout with cogwheeling noted at the wrist (R>L), bradykinetic execution of finger tapping and rapid opening-closing of hands. Also new were dysrhythmia of finger tap and rapid alternating movements, end-target dysmetria on finger-nose-finger, and slow turning and decreased right arm and leg swing.

The patient thereafter continued to decline and was provided symptomatic management of their symptoms. The patient died XX years after presentation and autopsy was performed.

**Example 4:**

Neuropathological definitive diagnosis: Multifocal areas of ischemia, infarction, and necrosis of different ages throughout the brainstem

DIAGNO/Neurologist Dx:

|  | Top 1 Dx | Top 2 Dx | Top 3 Dx |
| --- | --- | --- | --- |
| DIAGNO | Brainstem Glioma | Bickerstaff Brainstem Encephalitis | Multiple Sclerosis |
| Neurologist | Acute demyelinating encephalomyelitis | Autoimmune encephalitis | Brain abscess |

Case summary: A XX-year-old individual developed headaches, right-hand numbness, dizziness, and nausea one day after visiting a friend whose baby was vomiting. The next day, the patient visited an OSH and was told they had a viral illness, hydrated and sent home. Two days later, the patient developed severe vertigo and was unable to ambulate without leaning on walls. The patient's PCP prescribed antiemetics and ordered blood work, which was unremarkable. By day 11 of the illness, the patient's headache had resolved, but vertigo continued to worsen. Now the patient also had numbness in their right hand and the right side of their face; they had also had hiccups for several days. The patient was referred to an OSH ED, where a head MRI showed an abnormality, and the patient was transferred to MGH for further workup.

The patient's past medical history included a heart murmur diagnosed in childhood and an Achilles tendon rupture and repair in college. The patient was born in the US of African descent. The patient had limited travel, with their last trip being to London XX months earlier; they had an episode of “food poisoning” XX months before presentation. The patient had no history of tick bites, TB exposure, or pets at home. There was no history of genital ulcers. The patient had three lifetime sexual partners with most recent contact a few days prior to onset of symptoms. The patient's parents and siblings were alive and well. The patient worked at a genetics lab as a technician.

On initial exam at MGH, 12 days after symptom onset, the patient was afebrile with normal heart rate, respiratory rate and blood pressure. The patient was alert and oriented with normal speech. Pupils were equal and reactive, visual fields were full, and fundi were unremarkable. Extraocular movements were intact, but with saccadic intrusions and direction-changing end-gaze nystagmus on lateral gaze bilaterally. There was also a subtle ‘catch up’ saccade when the patient's head was turned from right to center. Sensation to light touch and temperature were decreased in the right V2-3 region. The patient's tongue deviated slightly to the right at rest. Other cranial nerves were normal. Motor strength was 5/5 throughout, and the patient had a right pronator drift. Sensation to light touch was decreased on the right arm, chest, and back as compared to the left. However, sensation to pinprick and temperature was decreased on the left as compared to the right. Reflexes were 2+ throughout. Cerebellar exam was normal on the left side, but on the right the patient exhibited dysmetria during finger-nose-finger and markedly impaired heel-knee-shin. Gait was severely impaired, with assistance needed for balance despite a wide, shuffling gait and laterally-extended arms.

Body imaging, including PET/CT, was unremarkable. A range of serologic studies were also unremarkable. Two days after admission, the numbness became more severe and extended to the patient's right leg. The patient's voice became hoarse, and their tongue deviated slightly to the right on protrusion. An LP was performed, which revealed a mild lymphocytic infiltrate (8 nucleated cells, 97% lymphocytes) and normal glucose, protein, and RBCs. The following day, the patient reported new right leg weakness and difficulty swallowing. Swallowing worsened over the next week and by day 12 of the patient's MGH course they underwent an elective tracheostomy and gastrotomy tube placement. A diagnostic procedure was attempted two days earlier.

The patient was begun on treatment for a presumptive diagnosis. The patient developed pneumonia and fever which responded to antibiotics. By MGH day 23, motor strength in all extremities was 4/5 except the patient's right leg, which they could only hold against gravity for 3 seconds. Three days later, the patient's eyes were deviated to the left and unable to cross the midline, and the patient was unable to move their tongue. Over the next several days, the patient's eye movement deficits evolved such that they had only subtle left deviation with slight difficulty in full leftward gaze. The patient had no movement of the right eyebrow with some movement of the left.

Weakness progressed over the next 2 weeks until all motor function was lost except for extraocular muscles. The patient now had no eye abduction, but had preserved adduction to command as well as with any effort at horizontal gaze; the patient was unable to move their eyebrows but had only a mild right facial palsy. Imaging showed progression, while the LP remained unremarkable.

By a month later, the patient was unable to respond to commands even with eye movements. The patient was discharged to a rehab facility where they developed hemodynamic instability prompting readmission. Now three months after onset of symptoms, the patient's pupils were fixed at 7 mm, and the patient had no dolls-eye movement or gag to deep suction, but reflexes were brisk and symmetric. The patient died two days later and an autopsy was performed.

**Example 5:**

Neuropathological definitive diagnosis: Bilateral hemorrhagic cerebral infarcts secondary to Cortical Venous Thromboses with secondary intracerebral hematomas

DIAGNO/Neurologist Dx:

|  | Top 1 Dx | Top 2 Dx | Top 3 Dx |
| --- | --- | --- | --- |
| DIAGNO | Amyotrophic Lateral Sclerosis (ALS) | Alzheimer's Disease | Frontotemporal Dementia (FTD) |
| Neurologist | Frontotemporal dementia with motor neuron disease | Alzheimer's disease with concomitant amyotrophic lateral sclerosis | Multisystem proteinopathy |

Case summary: A XX-year-old right handed patient had increasing difficulty remembering the details of conversations over the course of the previous two years. According to their brother, who lived next door, and another informant, there was no objective evidence of memory impairment although they did describe some increasing word finding difficulties. There were episodes in which the patient was not able to find their way in familiar settings as well as navigating at a hotel. The patient felt that they were slower at all tasks than they had been, and reported that mental calculations (calculating a tip) were more difficult as well. The patient had some difficulty remembering to take their medications (including a thyroid medication). The patient managed their own finances successfully and cared for their dogs.

The patient's past medical history included hypertension, hypothyroidism, osteoporosis, Lyme disease (treated with antibiotic course) and cataract removal. The patient's family history included a sister who died of complications of an intracranial aneurysm and their father died at age XX following a stroke. The patient was a former smoker (stopped decades earlier), widowed and lived alone. The patient had graduated from college, lived in various places around the world with their partner, and had worked as a dog trainer.

The patient was assessed at an outside facility, with an examination which demonstrated no focal neurologic signs. Testing demonstrated average to high average performance on abstract reasoning, set-switching/multitasking, attention span, rapid word generation and naming. The patient had initial difficulty with memory, recalling 3, 6, 7 and 8 words across four trials of a 9 word list; the patient recalled 8/9 at 30 seconds and 6/9 at 10 minutes. The patient's recall of a complex geometric figure was impaired, as the patient was unable to identify the figure on a page with 6 choices and did not improve their performance with 3 trials. Imaging studies were performed and the patient was followed by an outside physician.

About XX months after that assessment, the patient was seen by their primary care provider for routine follow up. The patient was late for the visit, having gotten lost trying to find MGH. The patient was unsure of their upcoming medical appointments (when, with whom and why). The patient's PCP suggested a driving evaluation although the patient resisted this idea.

The patient was seen at MGH XX months later after they complained of reduced dexterity in both hands (left worse) such that they had difficulty doing buttons and writing. The patient felt that this had been progressing slowly over months. The patient had started to need to use their arms to rise from a chair. The patient was able to walk, run short distances and go up and down stairs. The patient had not fallen, did not report any sensory changes, had no difficulty breathing, speaking or swallowing and had not had cramping. The patient's exam was notable for the presence of saccadic intrusion on smooth pursuit and a few beats of nystagmus bilaterally on end gaze but otherwise cranial nerves were normal. Muscle tone was normal, but the patient had mild weakness in arm abduction, external rotation, and extension of elbow, wrist and fingers bilaterally (but L>R), wrist flexion on the left, and thumb flexion bilaterally. In the patient's lower extremities, there was mild weakness in hip flexion, abduction, knee flexion, and foot dorsiflexion bilaterally, worse on the left. There were frequent fasciculations in the arms and forearms bilaterally. Alternating movements were slow but intact and finger nose finger was within limits of weakness.

Reflexes were brisk in the upper limbs except for the triceps which were diminished bilaterally, more on the right side (site of prior trauma). Quadriceps reflexes were normal and gastrocnemius reflexes were diminished bilaterally. There was no clonus. Plantar responses were equivocal to mute. The patient had a jaw jerk and bilateral palmomental reflexes. Vibratory sensation was absent in the distal lower extremities, and slightly reduced in their fingers.

The patient had difficulty rising from a chair without the use of their upper limbs. The patient swayed prominently on Romberg testing but did not fall. The patient could walk on their toes but had difficulty walking on their heels even with support for balance. The patient could not tandem walk. Laboratory findings included normal vitamin B12, copper, zinc, and folate and negative serology for syphilis. Additional studies were performed.

On follow up XX months later, the patient was repeating questions and laughing frequently during discussion of their test results and disease. It was not evident that the patient understood their evolving disease. The patient's exam was unchanged apart from evident atrophy of the intrinsic muscles of the hands and frequent fasciculations now also seen in their calves.

Four months later the patient was found at home unresponsive by the visiting nurse. The patient was taken to an outside ED where they had a witnessed seizure. Vital signs were notable for a blood pressure of 201/99, O2 sat 97%, heart rate 66. The patient was unresponsive to sternal rub with pinpoint pupils. After reviewing imaging findings, no intervention was performed and the patient received care directed at comfort. The patient died shortly thereafter (age XX; 5 years after the onset of their memory complaints and less than a year since the onset of their weakness) and an autopsy was performed.

**Example 6:**

Neuropathological definitive diagnosis: Acute hypoxic/ischemic changes

DIAGNO/Neurologist Dx:

|  | Top 1 Dx | Top 2 Dx | Top 3 Dx |
| --- | --- | --- | --- |
| DIAGNO | Paraneoplastic Encephalitis | Autoimmune Encephalitis | Neurodegenerative Disease (e.g., Lewy Body Dementia) |
| Neurologist | Paraneoplastic syndrome (Ex anti Ma2) | Viral basal ganglia encephalitis (ex West Nile Virus) | Fungal basal ganglia encephalitis (ex Cryptococcosis) |

Case summary: A XX-year-old patient presented to a community hospital with increased frequency of panic attacks. For the past several years, the patient had been under significant stress because they cared for their partner, who had advanced Alzheimer disease. Their partner was recently admitted to a nursing home for increased medical needs, with the transfer of care increasing the patient's stress levels. At the initial presentation, the patient reports increasing frequency of panic attacks, SOB, and increased tremor in the left foot with new right foot tremor. Pharmacologic intervention was undertaken. A few months earlier, the patient had injured their left foot which had increasing pain and burning sensation, for which they received treatments.

The patient's past medical history included hypothyroidism, coronary artery disease s/p stent to right coronary artery XX years earlier, anxiety, hypertension, osteoarthritis, possible Sjögren’s, and polymyalgia rheumatica for which they were maintained on daily prednisone.

A few weeks later, the patient returned to the hospital with more vigorous tremors, hallucinations, agitation, and fever. No exam from the presentation to the outside hospital is available and the historical details are limited. The patient was started on a sedative drip, intubated for airway protection, and transferred to the ICU. While in the ICU, the patient had two episodes concerning for seizure. LP showed colorless CSF with protein 49, glucose 64, WBC 1, and RBC 1083. A range of studies were sent to the lab and several days later, the patient was transferred to MGH for further diagnostic evaluation and continuation of care.

On arrival, the patient was sedated, intubated, and on a ventilator. Temperature was 38.7 °C, heart rate 105, and BP 177/76. Cardiac, abdominal, and respiratory physical exams were normal. Sedation was decreased for better physical exam assessment of the patient. There was no spontaneous eye opening and the patient had a left gaze preference that did not cross midline. Pupils were equal and reactive to light. There was a weak cough to suctioning and an absent gag reflex. The left ankle was contracted with a limited range of motion. The patient inconsistently followed commands, sometimes showing their thumb and squeezing their hand. Deep tendon reflexes were increased left > right. Hoffmann's reflex was absent. Marked increased tone was present in the left upper and lower extremities.

After sedation was decreased, the patient’s rigidity worsened and they were ultimately restarted on a sedative drip to manage both sedation and muscle rigidity. An LP performed at MGH showed an opening pressure of 7 with clear, colorless CSF, WBC 6>1 (94% lymph), RBC 93>11, glucose 70 and protein 33. Multiple additional studies were performed including laboratory and imaging investigations. PET imaging demonstrated a lung mass, with endobronchial biopsy showing a carcinoid tumor. Additional abnormal findings were obtained. A series of treatments were initiated.

Two weeks later, the patient remained hospitalized with no significant improvement in their condition. The patient now did not open eyes to stimulation and followed no commands and only grimaced to noxious stimuli. Their pupils symmetric and reactive. Tone was high in all extremities except the right upper extremity. Given the poor response to therapy and failure to improve, goals of care were shifted to ensuring the patient's comfort. A month after being transferred to MGH, approximately two months after the onset of their symptoms, the patient died and an autopsy was performed.

**Example 7:**

Neuropathological definitive diagnosis: Hydrocephalus

DIAGNO/Neurologist Dx:

|  | Top 1 Dx | Top 2 Dx | Top 3 Dx |
| --- | --- | --- | --- |
| DIAGNO | Progressive Supranuclear Palsy (PSP) | Alzheimer's Disease (AD) | Corticobasal Degeneration (CBD) |
| Neurologist | Progressive Supranuclear Palsy (PSP) | Corticobasal degeneration (CBD) | Multiple system atrophy (MSA) |

Case summary: A XX-year-old individual had progressive memory loss and confusion over 18 months. During the final 2 months the patient became unable to care for themselves and started to have difficulty walking. When the patient fell and hit their head, they were admitted to an outside institution. The exam showed decreased attention and inability to count backwards from 20. The patient had extremely poor naming, and was unable at times to name their own children. There was a square wave jerk nystagmus on primary gaze. The patient had increased tone bilaterally with some cogwheeling. The patient did not have any resting tremor but did have stimuli and startle myoclonus. The patient also had extremely brisk reflexes with Hoffmann in bilateral upper extremities; jaw jerk, snout and strong grasp reflex were evident. The patient's gait was slow, narrow based with small shuffling steps requiring much assistance. Imaging and EEG were abnormal. Lumbar puncture showed few cells (3 WBC, 4 RBC), glucose 79, protein 33, VDRL non-reactive, 14-3-3 equivocal. Other notable studies included anti-thyroglobulin antibodies (17) but negative studies for Hu, Ri, Yo, Ma and Ta, normal ACE levels. Various treatments were provided and the patient was discharged to a rehabilitation facility with some improvement.

Past medical history was notable for glaucoma and thyroid disease. The patient did not smoke and drank very little. The patient exercised regularly with walking and stretching. The patient was widowed and had worked as a legal secretary. The family history was notable for stroke, cancer and hypercholesterolemia.

The patient was seen at MGH several months after their discharge to rehab. The patient's daughter reported improved ability to take care of themselves and walk. The patient required assistance in dressing and bathing but was able to feed themselves and participate in group home activities. The patient was oriented only to person. The patient was able to name their children without errors, and named 3 out 5 objects. The patient's motor exam was essentially unchanged in tone however, the patient had less stimulus myoclonus. The patient was now able to ambulate without assistance across the room with improved speed and steadiness although still with small and narrow based steps. At a follow up visit a few months later, continued improvement was noted with more fluent language, naming 5 of 5 objects without hesitation. The patient was able to follow two-step commands accurately, also an improvement. Short term memory remained poor (0/3 objects including for hidden objects). The patient's nystagmus had resolved. The patient's reflexes were still brisk but the patient had lost the palmomental and rooting reflexes. The patient had a brisk and steady gait, narrowed based with regular steps. The patient was able to draw a clock face but failed at indicating the requested hand position.

About a year later, the patient's exam showed increased Gegenhalten and tone and some difficulty with sequential movements. The patient had only 4+/5 strength of their hip flexors, knee extensors. The patient's reflexes were brisk as before including grasp reflex, palmomental and glabellar tap. The patient's gait was fairly good but the patient had decreased right arm swing and turned en-bloc with some unsteadiness. The patient was able to generate 4 animals and 1 ""A"" word in a minute. There were no anti-thyroglobulin antibodies detected and the patient's CSF remained unremarkable. An additional intervention was undertaken, but without clear response. Three months later, the patient was seen after an episode in which they left their assisted living facility, inappropriately dressed and confused about their purpose. The patient's exam was unchanged. Over the next several years, the patient continued as a relatively stable set of deficits and findings on exam.

About five years after the patient's first assessment at MGH, impaired upgaze was noted for the first time. Intermittently, the patient showed mild retropulsion in their gait. The following year, some vertical gaze was further impaired with the addition of mildly restricted downgaze. At this time the patient generated 2 animals and 1 ""S"" word in a minute. The patient was oriented to season and year (better than in the past). The patient was able to count backwards from 20 accurately, but required 2 prompts to do the days of the week backwards, and made 1 error. The patient had partial recall of 1 object at 5 min (rhinoceros instead of hippopotamus) although had normal registration. The patient was able to ambulate without assistance.

A year later, the patient's exam was notable for very poor orientation but better word generation (6 animals and 4 ""A"" words in a minute). The patient's gait was slow and hesitant, but not wide-based or shuffling. The remainder of the patient's exam was stable. The patient was able to ambulate without assistance, but holding on to furniture. On exam 8 months later, the patient was only mildly inattentive, requiring 1 prompt to complete days of the week backwards (1 error). The patient produced 7 fruits & vegetables and 8 ""S"" words (including ""subordinate"" and ""salary"") in 1 min. The patient's recall was still impaired with 0 of 3 at 3 minutes.

Eight months later, there was evidence of some sleep-wake cycle confusion. The patient's blood pressure was 122/72 lying and 104/78 standing; the patient was symptomatic with the change. The patient performed days of the week without prompting or errors and produced 2 animals and 7 ""A"" words in a minute. The patient's gait was more hesitant than in the past, with some freezing at doorways.

Over the next few years, the patient developed increasing sleep-wake cycle problems and was noted to fall asleep during clinic visits. The patient required a ride-sided enucleation after a globe rupture (unclear etiology but not believed to be traumatic). The patient was much more inattentive, unable to perform days of the week testing, generated 1 animal in a minute and required assistance to walk. The patient died at age XX (XX years after their first visit to MGH) and an autopsy was performed.

**Example 8:**

Neuropathological definitive diagnosis: Atypical lymphoid infiltrate, most severe in the right frontal and temporal cortices

DIAGNO/Neurologist Dx:

|  | Top 1 Dx | Top 2 Dx | Top 3 Dx |
| --- | --- | --- | --- |
| DIAGNO | Progressive Multifocal Leukoencephalopathy (PML) | Creutzfeldt-Jakob Disease (CJD) | Autoimmune Encephalitis |
| Neurologist | Seizures | Encephalitis | Meningitis |

Case summary: A XX-year-old individual had sudden onset of expressive aphasia and confusion. The patient was brought to an ED where their NIHSS was 7 for confusion, inability to follow commands, left upper extremity drift and extinction to double simultaneous stimulation on the right. CT was unremarkable so the patient received IV tPA for presumed stroke. Subsequent imaging was abnormal. The patient's speech returned to normal the following day. A full workup was done and was unremarkable. Medical management was begun and the patient was discharged home.

The patient's past medical history was significant for hypertension, hyperlipidemia. The patient was a retired mechanic who lived at home with their partner and enjoyed gardening.

The patient did well at home for three months until they began experiencing multiple short episodes of left hand and foot numbness and weakness – these began in the hand and then spread up the arm and down the leg. They progressed over several days from occurring only rarely and lasting only 1 minute at a time to occurring several times per day and lasting up to 15 min followed by some confusion. The patient's family also felt that the patient had become generally more fatigued.

The patient was admitted to MGH. The patient's exam was notable for fluent speech, some difficulty following commands and trouble with calculations. The patient had a left sided pronator drift, decreased dexterity in the left hand, leaned to the left when seated and had an unsteady gait with intermittent falling toward the left. Studies were performed and the patient's medication was adjusted. The patient improved and was discharged home, although at home experienced several more episodes prompting a medication adjustment.

One month later, the patient was admitted to MGH with confusion, urinary incontinence, fevers and muscle rigidity. The patient was not speaking and was unable to follow commands. The patient was diffusely rigid with hyperreflexia and clonus in the lower extremities. A range of additional studies were done and were mildly abnormal. The patient was started on two different treatments, one of which the patient responded to rapidly and the patient was again discharged to rehab. The patient's fevers returned so the patient was started on a maintenance medication and again, the patient improved symptomatically.

Two months later, the patient was readmitted to MGH with recurrent fever and encephalopathy. The patient's management was again adjusted and the patient returned to the rehab facility with a short term plan of therapy. Three weeks later, the patient again developed a fever to 102 in the setting of their medication adjustment, prompting alterations and the patient again improved. Despite this, the patient was readmitted a week later with confusion, incontinence, and a transient episode of left sided facial droop. During this admission, a range of studies were again performed and were markedly abnormal and changed from prior. The patient was again managed medically, including with a new agent.

The patient was seen for short interval follow up in clinic and on exam was found to be bradyphrenic. The patient was able to register only 1 out of 3 words, was unable to recall any of the words even with prompting and could follow only one step commands. The patient required help with transfers and even with one assist the patient's gait was slow and unsteady. Two weeks later, the patient again returned to MGH with a high fever. The patient's treatment plan was continued.

The following day the patient's partner found the patient unarousable at home with a left facial droop (the patient had been increasingly lethargic at home with multiple episodes of diarrhea). The patient had not had fever or other infectious signs or symptoms. The patient was treated acutely, intubated and admitted to the BWH ICU. The patient's exam was notable for more sluggish movements on the left. Repeat studies were done and diagnosis was established. Following this, the patient had “disorganized gaze” and occasional left-sided gaze preference. Over the subsequent days the patient began to have eye opening to voice although the patient did not follow commands. The patient was extubated and eventually PEG was placed. At discharge a week later, the patient's exam was notable for orientation to place and self, hypophonic and dysarthric voice. The patient was intermittently following simple commands. The patient had no gaze preference or facial asymmetry. The patient moved all extremities purposefully and had brisk reflexes including crossed adductors but toes were downgoing bilaterally.

Approximately one week after discharge, the patient was transferred back to MGH for additional treatment. Shortly after this, the patient developed a viral infection and probable UTI. Despite treatment, the patient became increasingly somnolent and continued to worsen. Finally, after several family meetings, the patient was transitioned to comfort-focused care. The patient died shortly thereafter (seven months after presentation) and an autopsy was performed.

**Example 9:**

Neuropathological definitive diagnosis: Bilateral subdural hematomas, with mild right to left subfalcine herniation

DIAGNO/Neurologist Dx:

|  | Top 1 Dx | Top 2 Dx | Top 3 Dx |
| --- | --- | --- | --- |
| DIAGNO | Paraneoplastic Encephalitis | Chemotherapy-Induced Neurotoxicity | Progressive Multifocal Leukoencephalopathy (PML) |
| Neurologist | Toxic-metabolic encephalopathy | Brain abscess | Meningitis |

Case summary: A XX-year-old patient was seen by the Neurology Consult Service because of changes in their speech and orientation while an inpatient at MGH. The patient was 7 days into an admission for treatment of relapsed treatment-refractory lymphoma (DLBCL) when these symptoms emerged.

The patient's past medical history was notable for a diagnosis of marginal zone lymphoma made XX years previously on a lymph node biopsy. Staging at that time demonstrated no involvement of their bone marrow and the patient was treated with rituximab and bendamustine, with good response. XX years later, a biopsy of a retroperitoneal lymph node demonstrated non-germinal cell DLBCL. This prompted several lines of treatment, including standard combination chemoimmunotherapy, a BTK-inhibitor-based regimen, and a platinum-based salvage regimen with dose modification for toxicity. Finally, the patient was admitted to MGH for another approach to treatment. Otherwise, the patient's past medical history was notable for squamous cell carcinoma on their hand (XX years ago). The patient had a modest smoking history and a history of breast cancer in their sibling. There was no other family history of neurologic disease.

At this admission to MGH, the patient reported fatigue, leg swelling, difficulty urinating, and back pain and poor appetite but denied fevers, chills, night sweats and weight loss. The patient was alert, oriented with appropriate mood and affect. There were no focal neurologic findings. One day after initiation of treatment, the patient developed persistent low grade fevers without identification of an infectious source. On the fifth day, the changes in speech and orientation prompted a consultation request.

When the patient was examined, they were awake and alert but inattentive. The patient was not able to provide any description of their deficits or course. Their speech was fluent but perseverative and they were unable to count backwards from 20. The patient was oriented to location (MGH), but not date or time of day (morning vs. evening). The patient had difficulty naming their hometown and favorite sports team. The patient followed commands well. The remainder of the elemental neurologic exam revealed no abnormalities in the cranial nerves or sensation but there was mild leg weakness. The patient had diffuse myoclonus and asterixis. Their labs included Cr 2.16 (up from their baseline of 1.4), BUN 62, LDH 519 (had been as high as 1210 earlier in the hospitalization).

Medical intervention was begun which alleviated the patient's symptoms temporarily although their symptoms continued to fluctuate over the next two days. The patient's exam began to show more diffuse weakness and there was need for increased encouragement to get them to follow commands. The patient's speech now included paraphasic errors.

Seven days after therapy, the patient was difficult to rouse in the morning. When examined, the patient was lying in bed with their eyes closed and their head thrashing back and forth. The patient mumbled incomprehensively and could not follow commands. The patient blinked to threat bilaterally, had intact extraocular motions without facial asymmetry and was moving all extremities. Medical management did improve the patient's level of alertness but they were agitated and unable to follow commands. Imaging was obtained, complicated by an episode of a dystonic medication reaction with orobuccal dyskinesia and torticollis which resolved over hours.

A few days later, the patient seemed slightly less obtunded: they opened their eyes after a lot of noxious stimulus, but then kept them open, tracked the examiner, and followed commands to wiggle their toes; the myoclonus was reduced as well. Over subsequent days, the patient's neurologic status continued to fluctuate, and they remained moderately obtunded at best, although at times they opened their eyes easily in response to the examiner, smiled when following simple commands and could say “hi”. Imaging and other studies were performed; although possible interventions were considered, comorbidities prevented them from being implemented.

The patient's prognosis was felt to be very poor given likely worsening of their underlying illness (obstructive renal failure emerged over time, likely associated with retroperitoneal adenopathy) despite the aggressive therapy they had undergone. The patient developed worsening obtundation, orofacial dyskinesias and intermittent periods of apnea. When multiple treatments were attempted without benefit, discussions with the patient's family led to a shift in goals of care. The patient died a month after initiation of treatment and an autopsy was performed.

**Example 10:**

Neuropathological definitive diagnosis: White matter and axonal injury

DIAGNO/Neurologist Dx:

|  | Top 1 Dx | Top 2 Dx | Top 3 Dx |
| --- | --- | --- | --- |
| DIAGNO | Progressive Multifocal Leukoencephalopathy (PML) | CNS Lymphoma (especially Primary CNS Lymphoma) | Paraneoplastic Neurologic Syndrome |
| Neurologist | Neurologic side effects of checkpoint inhibitors | Immune effector cell-associated neurotoxicity syndrome (ICANS) of CAR T-cell therapy | CNS lymphoma with ocular involvement |

Case summary: A XX-year-old individual developed multiple neurologic deficits including visual impairment in the setting of treatment for an underlying malignancy. The patient had been in their usual state of health until approximately XX years earlier when they were discovered to have an elevated BUN and creatinine on routine screening labs. They were worked up for this and eventually found to have hydronephrosis associated with a retroperitoneal mass. A CT-guided biopsy was performed and demonstrated DLBCL. Staging was performed and there was no evidence of CNS involvement. The patient was treated with six cycles of chemotherapy, as well as IV medication for CNS prophylaxis. They subsequently received 36 Gy of radiation to the retroperitoneal disease. PET-CT revealed persistent FDG avidity and the patient underwent a biopsy of a ureteral site of uptake which was consistent with scar tissue.

Approximately a year after diagnosis, the patient developed leg swelling and was found to have progressive nodal disease on imaging. They underwent 3 cycles of chemotherapy and then underwent BEAM autologous stem cell transplant complicated by GI toxicity, febrile neutropenia, and slow blood count recovery. A PET-CT performed six weeks after transplant showed likely recurrent disease. A biopsy of the right internal iliac node was unsuccessful but subsequent scans showed stable adenopathy. Twelve months later, scans showed periaortic nodal progression (confirmed by biopsy as recurrent large cell lymphoma) and PET/CT demonstrated FDG avid lymphadenopathy above and below diaphragm.

The patient was enrolled in a clinical trial for refractory DLBCL and admitted. At the time of admission, they felt well and had no HA, fevers, chills, dizziness, lightheadedness, vision changes, nausea, vomiting or weakness. Their strength was reported as intact throughout as was sensation.

After a month in the trial, PET studies demonstrated complete remission. However, the patient had begun to develop cognitive problems in both memory and attention (reported by both the patient and their partner) as well as worsening vision. They had a series of increasingly frequent falls. The patient was seen several times in the ED and received fluid and blood component support. After one fall with head-strike, they had a head CT which was unremarkable.

The neurologic exam approximately 6 weeks after entering the clinical trial, showed the patient to be only oriented to themselves and place. Their speech was fluent without paraphasic errors, and they could perform serial 7s. Their recall was 2 out of 3 words at five minutes. Cranial nerve exam was notable for left visual field deficit, decreased activation on right side of face and a hoarse voice. There was pronator drift on the right but otherwise no evidence of weakness. The patient had difficulty standing on their own, requiring assistance.

Imaging studies were performed and were abnormal. That night, the patient developed a fever and was admitted. Their examination at that time was consistent with the above, but was also noted to have profound vision loss. Ophthalmology noted that the right eye was 20/800, and the left eye was 20/400 with severe visual field constriction bilaterally in all quadrants. The funduscopic exam was noted to be normal. The patient had hip flexor 3/5 hip flexor weakness bilaterally but there was noted limitation by pain. They had decreased reflexes at the patella and ankles. Given the worsening symptoms, the patient underwent further therapy.

Initially there was some improvement in vision with the treatment, but ultimately it progressed. Repeat examination was notable for only seeing fingers in central vision, as well as achromatopsia. The patient underwent further ophthalmological testing, as well as an LP (1 nucleated cell [52 lymph, 40 monos, 6 neutrophils, 2 other], glucose 103, protein 39). An additional series of studies were sent.

Eventually, the patient was only able to see shade and light and dark. Despite an apparent complete remission of the lymphoma, the patient had progressive and psychologically devastating neurologic deterioration. They indicated that they desired no additional significant life-sustaining therapies and were discharged to hospice. The patient died approximately 5 days later (XX years after presentation with DLBCL and about 2 months after developing the neurologic deficits) and an autopsy was performed.

**Example 11:**

Neuropathological definitive diagnosis: Metastatic small cell carcinoma of the lung DIAGNO/Neurologist Dx:

|  | Top 1 Dx | Top 2 Dx | Top 3 Dx |
| --- | --- | --- | --- |
| DIAGNO | Paraneoplastic Cerebellar Degeneration (PCD) | Creutzfeldt-Jakob Disease (CJD) | Wernicke's Encephalopathy |
| Neurologist | Opsoclonus-myoclonus-ataxia syndrome | LGI1 encephalitis | Paraneoplastic encephalitis |

Case summary: A XX-year-old patient initially developed nausea and vomiting. The patient was seen by their physician and a new diagnosis of hyperthyroidism was established. Over the next several months, the patient had multiple medical visits without any clear diagnostic abnormalities. About 2 months after the initial presentation, the patient was due to see their PCP but their gait was so unsteady that they were unable to go to the office. By this time, the patient had also had a 50-pound weight loss. Over the next two days, the patient developed severe dizziness worse than prior, difficulty walking, and abnormal eye movements. The patient was not able to stand, which exasperated their dizziness. The patient had not had fevers or neck stiffness, nor focal weakness or episodes of loss of consciousness or seizure-like activity. The patient was admitted to MGH via the ED.

The patient's past medical history included hyperthyroidism, hypertension, peptic ulcer disease, and hemochromatosis. The patient lived with their partner and dog. The patient had smoked for XX years, transitioning to vaping XX years before presentation. The patient drank an occasional beer and denied any other drug exposure. The patient had been self-employed as an insurance broker until these symptoms emerged. There was no family history of neurologic disease.

Neurologic exam demonstrated the presence of irregular jerking eye movements in all directions. Eyes were conjugate and range of ocular motions were normal. Pupils were reactive (3mm->2mm) bilaterally. There was no evidence of abnormal movements of the upper or lower extremities except for mild tremulousness. The patient had a wide-based, hesitant gait, but was able to take short steps with two-person assistance. An LP demonstrated normal IgG index and no oligoclonal bands, WBC were 5 and 8, with some RBC contamination. Protein and glucose were normal. A range of additional studies were performed which revealed abnormalities, including on chest imaging. This was subsequently assessed by fine needle biopsy and a treatment regimen was established. The patient was subsequently discharged to Spaulding.

The patient was seen in clinic about 2 months later. The patient scored 14 of 30 on MoCA, having trouble with the trails test, copying a cube, putting the correct time on a clock, paying attention for the letter A, and difficulty naming F words in a minute, coming up with 9, but with 5 of them repeated. The patient also had no recall of 5 words and had problems with orientation to date (off by six months) and place (thought they were at a rehabilitation hospital). There was perseveration and mild apraxia when asked to write with a pen. The patient still had constant irregular movements of their eyes. Visual fields were full to confrontation. Pupils equal and round, 2 mm. There was no ptosis and the EOM were intact. Light-touch sensation on face was normal bilaterally. There was no facial asymmetry, with a normal forehead wrinkle, blink, smile, and cheek puff. The tongue and palate were midline with protrusion and elevation, respectively. There was no dysarthria. The hearing was grossly normal to finger rub bilaterally. The patient had had periodic myoclonic jerks of their upper body, particularly while falling asleep during the visit. There was decreased vibration sensation in their lower extremities > fingers. DTRs were decreased (1+) but symmetric throughout.

The patient subsequently had a long (multi-month) hospitalization at MGH complicated by pneumonia and development of atrial fibrillation. The patient received a wide range of attempted therapeutic interventions during this hospitalization focused on their underlying illness and their neurologic symptoms. Brain imaging during this hospitalization showed improvement in some findings. After discharge, the patient was seen in clinic where they were using a walker. Their cognition remained poor (MoCA: 12). Their continual eye movements were still present.

The patient was hospitalized again shortly after this visit with orthostatic hypotension, worsening atrial fibrillation, and persistent pneumonia, eventually being intubated and cared for in the MICU. The patient's medical management continued throughout this time. The patient developed diaphragmatic myoclonus, which made it difficult to wean them from the ventilator and required placement of trach and PEG. The patient developed gastroparesis, which made it very difficult to feed them as they kept vomiting up feeds. The patient eventually made it to the RACU, where for a few days they would awake to voice and follow some commands. The patient then decompensated and started having “storming” with fluctuating vital signs and rigid arms with contraction towards their body. The patient's persistent eye movements became even more pronounced as did the diaphragmatic myoclonus. The patient slipped into coma; after about a week, their family decided to focus care on comfort. The patient died the following day and an autopsy was performed.

**Example 12:**

Neuropathological definitive diagnosis: Multiple Sclerosis, chronic

DIAGNO/Neurologist Dx:

|  | Top 1 Dx | Top 2 Dx | Top 3 Dx |
| --- | --- | --- | --- |
| DIAGNO | Alzheimer's Disease | Frontotemporal Dementia (Behavioral Variant) | Vascular Dementia |
| Neurologist | Frontotemporal dementia with motor neuron disease | Spinocerebellar ataxia | Multisystem atrophy |

Case summary: A XX-year-old patient was assessed because of XX years of progressive memory loss. Their partner reported that they “lived in a different world.” The partner noted that the patient was unable to recall events that happened within the last 10 minutes and often could not locate their car in a parking lot. The patient also had gait difficulties and several recent falls.

Their past medical history included a breast fibroadenoma, a large rectal villous adenoma, nephrolithiasis, and carpal tunnel syndrome. Family history was significant for heart disease and stomach cancer in their father, heart disease in two of their siblings, and Type 2 diabetes in their mother. The patient smoked tobacco daily and drank over 4 cups of decaffeinated coffee. They denied EtOH or other substance use.

On review of systems, the patient noted urinary frequency and leakage, decreased hearing, fatigue, changes in weight, temperature sensitivity, depressed mood, and sleep problems. Their initial exam revealed normal vitals with no autonomic instability. Their MMSE score was 28 (unable to recall 2 words at 5 minutes). On neurologic exam, the patient was noted to be anxious with intact cranial nerves, increased left-sided tone, with bilateral ankle clonus, left sided ataxia on finger to nose, and an ataxic gait. Diagnostic studies were performed and a working diagnosis was established.

The patient was seen again 2 months later in follow-up. On this visit, the patient was noted to have an unusual affect with a complete lack of insight into their diagnosis and an inappropriate lack of concern regarding their deficits. The patient noted that despite being a dental assistant for 14 years, they had recently started handing the incorrect tools to the dentist. The patient developed worsening anxiety, poor sleep, and fatigue. Cognitive testing was performed and demonstrated poor performance in global cognitive functioning, memory, attention, and processing speed. Their speech was noted to be fluent with normal prosody and syntax but a complete neurologic exam was not documented. At this point, the patient was started on a medication for their condition.

Over the next few months, the patient continued to decline cognitively and was unable to continue working. The patient began having difficulties managing money and taking their medication correctly. Their partner noted that the patient would often come home from the grocery store with six of something that they already had. The patient endorsed dizziness in addition to continued poor sleep. Their clinical exam had worsened, with the development of hyperreflexia and a positive Babinski on the left. Their gait was described as “lurching.” The patient was started on a new medication and repeat testing was performed. Several weeks later, their clinical exam had improved slightly, but their partner noted that the patient played computer games all day and had impairments in driving.

A year after their first neurology visit, the patient continued to have memory problems but had improvement in their affect, coordination, and sleep. Seven months later, the patient developed worsening balance and had a fall. The patient was unable to stand for longer than 45 minutes due to fatigue. The patient had trouble with meal planning and following through with plans. On exam at this time, the patient smiled inappropriately, had normal speech and cranial nerves, slightly increased tone and mild hyperreflexia on the left, a clumsy left hand, and normal gait.

Approximately 20 months after their first neurologic assessment, the patient developed chest pain and presented to an OSH. The patient was diagnosed with a STEMI and transferred to MGH for initiation of ECMO. The patient died on the 14th hospital day (age XX, approximately XX years after the onset of neurologic complaints) and an autopsy was performed.

**Example 13:**

Neuropathological definitive diagnosis: Granulomatous Ependymitis of unknown cause

DIAGNO/Neurologist Dx:

|  | Top 1 Dx | Top 2 Dx | Top 3 Dx |
| --- | --- | --- | --- |
| DIAGNO | Neurosarcoidosis | Paraneoplastic Syndrome | Central Nervous System Vasculitis |
| Neurologist | Neurosarcoidosis | Vasculitis (GPA > PCNSV) | CNS TB |

Case summary: A XX-year-old right-handed patient had an episode of severe headache, nausea, vomiting, vertigo, and confusion. Cranial CT was normal and their dose of a corticosteroid, which they had been on for polymyalgia rheumatica, was increased. Two weeks later, their symptoms worsened. Additional studies were performed, including MRI/MRA of the head and neck as well as an EEG. All were normal and their corticosteroid dose was tapered. They improved but continued to have episodic dizziness, trouble with memory, and imbalance.

In addition to polymyalgia rheumatica, their medical history included remote infectious meningitis, hypothyroidism, and pulmonary embolism s/p IVC filter placement. Family history was notable for a child with ovarian cancer and another with breast cancer. Prior to their illness, they lived by themselves. They visited Cuba for a week the previous year. They did not smoke, drink, or use drugs.

Approximately two months later, they had the acute onset of vertigo and nausea. They reported that they had turned their head to the left and immediately experienced vertigo, nausea, and vomited once. This lasted a few minutes, and resulted in admission to the neurology service of an outside institution. On exam, they were fully oriented with intact memory, attention, language, and praxis. There was no papilledema. There was rightward rotary nystagmus on primary gaze and lateral gaze bilaterally. Saccades were normal. Dix-Hallpike maneuver to the right elicited their symptoms. The rest of their cranial nerves were intact. Strength and sensation were normal. Reflexes were 1+ at the ankles, but otherwise normal. Plantar response was flexor. Coordination and gait were normal. MRI brain showed an abnormality. LP showed glucose 50, protein 113, WBC 25, and RBC 275. CSF cytology showed a T-cell predominant lymphoid profile, but no monoclonality. Negative work-up included CSF cultures, HSV PCR, CSF Tb PCR, CSF JCV PCR, CSF cryptococcal Ag, CSF ACE, ESR, CRP, HIV, TSH, anti-TPO Ab, and anti-NMDAR Ab. Serum paraneoplastic Ab panel showed elevated anti-striated muscle Ab (1:15360; NL < 1:60), anti-AChR binding Ab (0.79 nmol/L; NL ≤ 0.02) and modulating Ab (55%, normal 0-20). CT chest, abdomen, and pelvis showed a 5 mm right upper lobe lung nodule, numerous pancreatic cysts (enlarged from prior), stable hepatic hemangioma, and stable renal cysts. During their course, they developed ptosis and subtle facial weakness on the left. They also had new left-sided incoordination and an unsteady gait. They were then discharged.

A brain biopsy was performed and showed lymphohistiocytic inflammation with a necrobiotic nodule. They then developed bilateral hearing loss and more dizziness. They were readmitted to the outside institution. LP showed glucose 57, protein 187, RBC 419, WBC 99 (99% lymphocytes). They were begun on anti-mycobacterial therapy. They became intermittently somnolent. EEG showed bilateral theta slowing, more prominent on right, as well as brief runs of sharp waves on the right and generalized sharp waves seen frequently during patient stimulation. There was mild improvement after starting an anticonvulsant medication and they were discharged. Unfortunately, their symptoms worsened with intermittent disorientation, nonsensical speech and falls, prompting their family to bring them to the MGH ED for evaluation.

When seen at MGH (5 months after symptom onset), they were awake but inattentive. Their speech was fluent, but they could name only simple objects and followed 2-step commands intermittently. Their speech was perseverative. There were saccadic intrusions, but their face was symmetric and there was no nystagmus. Hearing was impaired bilaterally. Strength was normal. Reflexes were 1+ but absent at the ankles. Plantar response was flexor. They had mild appendicular incoordination, left greater than right. They were admitted for further evaluation. Repeat LP showed glucose 57, protein 126, RBC 98, WBC 18 (96% lymphocytes). An extensive work-up showed ANA 1:640 in speckled pattern and elevated ACE at 60. Negative studies included whole-body PET, CSF cytology and flow cytometry, and additional infectious (HHV6, EBV, CMV, toxoplasmosis, and fungi) and rheumatologic studies (IgG panel, C3, C4, Ro/La, Scl70, RF, CCP, dsDNA, ANCA). Their previous medication regimen was initially stopped, but restarted after a repeat biopsy showed necrotizing granulomatous inflammation. An additional medication was also started and they were discharged.

One month later, their family reported no significant improvement. They had persistent dizziness and seemed to be more anxious and depressed. They spoke of death and had hallucinations of their deceased parent. MRI brain showed mild interval improvement and a new medication was started. They developed swelling in the legs 2 months later and were diagnosed with bilateral DVTs. They were readmitted to MGH for initiation of anticoagulation and a MRI brain showed new changes. Their medication regimen was again escalated.

Given their age, course and status, the decision was made to provide palliative care, following discussion with their family. They remained in hospice for 4 months and died slightly more than a year after symptom onset and an autopsy was performed.

**Example 14:**

Neuropathological definitive diagnosis: Hereditary diffuse leukoencephalopathy

DIAGNO/Neurologist Dx:

|  | Top 1 Dx | Top 2 Dx | Top 3 Dx |
| --- | --- | --- | --- |
| DIAGNO | Progressive Supranuclear Palsy (PSP) | Corticobasal Degeneration (CBD) | Multiple System Atrophy (MSA-P for parkinsonian variant) |
| Neurologist | Corticobasal syndrome | Multisystem atrophy | Progressive supranuclear palsy |

Case summary: A XX-year-old individual developed problems with balance over 2-3 weeks. The patient found it hard to stay still when standing, feeling the urge to move around or shift balance. The patient would sometimes have to grab onto things to avoid losing balance. The patient denied numbness in their feet, shooting pains, back pain, urinary or bowel symptoms, nausea, vomiting fevers or chills.

The patient's past medical history included hypercholesterolemia and hypertension. The patient's family history was significant for a now-deceased uncle with a limp (either polio or MS according to family members).

Examination showed that the patient was alert and fully oriented. Cranial nerve exam was unremarkable apart from non-sustained horizontal nystagmus at extremes of gaze. The patient's strength was intact on direct testing and the patient could walk on their toes and heels, squat, and get up from a chair without difficulty. Reflexes were brisk in the legs but normal in the arms, while the patient's toes were considered equivocal but possibly upgoing. Heel-to-shin testing was “not perfect.” Finger-to-nose was “minimally off.” Sensory and vibration and position sense exams were normal. The patient's gait appeared steady but when the patient stood straight they felt slightly off balance; this did not worsen when closing their eyes. Imaging studies were performed and reported to be normal.

When assessed a year later, the patient reported that their legs continued to feel “a little wobbly and not very coordinated.” Examination only showed subtle cerebellar signs with a slightly positive Romberg sign. The patient received a trial of treatment without significant benefit. Imaging abnormalities were observed but not diagnostic.

Three years after the initial presentation, the patient was referred for evaluation. The patient now could not run, ride a bike, or talk and stand simultaneously. The patient had frequent episodes of tripping and felt more fatigued. The patient's speech had also deteriorated within the past year with poor articulation. The patient's partner felt that the patient was not as sharp as they used to be. The patient had noticed changes in their writing and buttoning their shirts. Examination showed the patient had trouble with Luria sequence. The patient could recall 4/4 after a few minutes’ time. CN II-XII showed mild saccadic intrusions, but was otherwise normal. Motor bulk, tone and strength was normal. Sensation was intact. The patient had positive grasps bilaterally with R worse than L (intermittently present). There was a mild snout reflex and a negative glabellar tap. RAMs were normal though slightly less coordinated with the left hand than right. There was no truncal ataxia. The patient was slow to initiate movements. Sequential finger taps were dysrhythmic (right worse than left). The patient had difficulty initiated and performing task with leg agility. The patient had mild difficulty with standing out of their chair with arms crossed. The patient's posture was slightly stooped and their gait was slightly wide based and cautious. The patient would sway toward the left when walking and sitting. The patient had postural instability and difficulty with toe walking. The patient could not tandem walk and had a hard time standing in tandem. There were times during the exam when the patient's left hand seemed to move involuntarily. Imaging demonstrated abnormalities which had progressed over time. The appearance prompted additional studies, resulting in a presumptive diagnosis being established.

A year later the patient's progressive cognitive decline (MoCA of 18) prompted initiation of medical management. Three months later, the patient was hospitalized after an episode of left-sided shaking; the patient was discharged after 4 days with a new medication. Over the next two years, the patient experienced worsening cognition, progression of their speech and mobility difficulties along with apraxia. The patient had several seizures as well.

At the time of the patient's last assessment, the patient was living at home with their partner but was at an adult day care twice a week. The patient needed full care for all personal hygiene needs and had to be fed, but could chew and swallow. The patient was wheelchair-bound. The patient responded to people and surrounding activity and had rare verbal outbursts. There were no spontaneous movement in the patient's arms or legs but the patient could turn their head. The patient was mute during the meeting with their physician. Examination showed that the patient could direct their eyes to the examiner and would follow with their eyes at times, demonstrating saccadic intrusions into pursuit eye movements and would blink to threat. There was paratonia throughout with almost no use of the right arm and leg. There was occasional “mini-myoclonus” of the fingers and hands and large amplitude fasciculations in the forearms. DTRs were 3 in the arms, 3 at the knees and 2 at the ankles. There was no clonus. Plantar responses were extensor on the right and flexor on the left. There was increased tone and extension of the fingers. Snout was not elicitable and there was a positive glabellar tap.

The patient died 4 months after the final assessment, approximately six years after presentation, and an autopsy was performed.

**Example 15:**

Neuropathological definitive diagnosis: Wolfram Syndrome

DIAGNO/Neurologist Dx:

|  | Top 1 Dx | Top 2 Dx | Top 3 Dx |
| --- | --- | --- | --- |
| DIAGNO | Multiple System Atrophy (MSA-C for cerebellar variant) | Spinocerebellar Ataxia (SCA) | Familial Alzheimer's Disease |
| Neurologist | Spinocerebellar ataxia (possibly type 7) | Leber's hereditary optic neuropathy | Multisystem atrophy |

Case summary: A XX-year-old individual had a year of progressive memory difficulties. The patient experienced difficulty recognizing people they had long known, seemed to remember them by how they looked years prior. The patient's partner reported ""they forget that they forget."" The patient tried using reminders for themselves, but forgot about them. The patient was no longer able to manage any finances (they were an accountant) and had been asked to leave several jobs as a comptroller.

In their early XXs the patient had developed bladder dysfunction of unclear etiology requiring intermittent strait catheterization. This was accompanied by subsequent erectile dysfunction. At age XX the patient developed polyuria and polydipsia and was diagnosed with DM (felt to be type 1). Also in their XXs the patient began dressing in strange color combinations and ultimately was diagnosed with color blindness in their XXs. At age XX the patient was found to have bilateral optic atrophy with preserved vision on routine ophthalmologic exam. Brain MRI at that time revealed severe atrophy of the cerebellar hemispheres and vermis, pons, and middle cerebellar peduncles as well as moderate cerebral atrophy. The patient was of Ashkenazi Jewish descent with no known consanguinity. The patient's mother died from melanoma. The patient's father died from multiple strokes and a myocardial infarction. One of the patient's adult children suffered from attention deficit hyperactivity disorder (ADHD) and Tourette syndrome. The patient's other child suffered chronic urinary tract infections. The patient's maternal grandmother had type 2 DM, and a maternal first cousin had type 1 DM. The patient did not drink alcohol and had no other toxic exposures.

At the patient's first visit, they demonstrated a blunted affect but were able to joke with their examiner. The patient was alert and oriented with fluent speech, intact naming and repetition. The patient named 13 “F” words in one minute. The patient had normal handwriting and clock drawing. The patient could follow complex three step commands. On memory testing the patient could not recall any of three objects at 5 minutes and got only 1/3 with multiple choice options. The patient's visual fields were full to confrontation. The patient had a slight left ptosis, bilateral end-gaze nystagmus, hypometric saccades, mild anisocoria (R>L) with both pupils slightly sluggishly reactive. There was disc pallor bilaterally on undilated funduscopic exam. Facial sensation and strength were intact as was hearing. The patient's palate moved symmetrically, their tongue was midline (but small as was their jaw). Motor examination was normal. The patient had decreased vibratory sense in their toes bilaterally. The patient performed well on finger-to-nose and heel-to-shin testing. The patient's gait was normal-based and steady, with good toe and heel walks. Tandem walk was poor, with tendency to veer to right. The patient's reflexes were 1+ throughout with downgoing toes.

Over the next several years the patient's memory continued to decline characterized by profound anterograde amnesia, with additional impairments in cognitive flexibility, executive function, naming, and high order visual processing skills. The patient developed symptoms of depression, which responded to treatment with an antidepressant medication. The patient developed progressive autonomic neuropathy with gastroparesis and severe postural hypotension, while their diabetes remained under excellent control. The patient's bladder dysfunction progressed requiring suprapubic catheterization at age XX. On examination at that time the patient appeared generally medically well. The patient's BMI was 25. Systolic blood pressure fell from 150 to 95 after one minute of standing, although without symptoms. The patient's affect was flat and they were passive throughout the interview, speaking only when spoken to. The patient was not oriented to time or place. The patient could repeat four words, but could not learn them despite multiple attempts. The patient was unable to provide information concerning major current political or national news. The patient could, however, recall sizable fragments of remote memory from their college years. Funduscopic examination revealed optic atrophy bilaterally with no sign of diabetic retinopathy. Visual acuity was 20/40 in each eye. Pupil responses to light and accommodation were normal. Eye movements were normal with the exception of saccadic intrusion into horizontal smooth pursuit. The patient had high tone hearing loss bilaterally and audiometry demonstrated moderate sensorineural hearing bilaterally. Muscle tone in the extremities was normal, bulk was intact, and strength was full. There was no evidence of dysmetria with finger-to-nose and heel-to-shin testing, and gait was slow but stable. Further studies were performed and treatment initiated rapidly but unfortunately with little clinical impact. The patient continued to progress and subsequent treatments were initiated to improve quality of life although a specific diagnosis was established.

The patient became wheelchair-bound and was dependent on their partner for all acts of daily living. The patient's language gradually worsened over several years. On examination at age XX the patient was slumped in the wheelchair, not interacting or talking, even though their eyes were open. The patient heard and responded well by showing fingers, raising a hand, closing eyes. The patient could open their mouth with coaxing. Tone in the limbs was normal, and power with formal testing in the arms was preserved when the patient could be coaxed to initiate and maximize, but power in the legs appeared to be reduced to antigravity at the hips and knees. The patient died at age XX (XX years after their first visit) and an autopsy was performed.
